# LDAK-KVIK performs fast and powerful mixed-model association analysis of quantitative and binary phenotypes

**DOI:** 10.1101/2024.07.25.24311005

**Authors:** Jasper P. Hof, Doug Speed

## Abstract

Mixed-model association analysis (MMAA) is the preferred tool for performing a genome-wide association study, because it enables robust control of type 1 error and increased statistical power to detect trait-associated loci. However, existing MMAA tools often suffer from long runtimes and high memory requirements. We present LDAK-KVIK, a novel MMAA tool for analyzing quantitative and binary phenotypes. Using simulated phenotypes, we show that LDAK-KVIK produces well-calibrated test statistics, both for homogeneous and heterogeneous datasets. LDAK-KVIK is computationally-efficient, requiring less than ten CPU hours and 5Gb memory to analyse genome-wide data for 350k individuals. These demands are similar to those of REGENIE, one of the most efficient existing MMAA tools, and approximately ten times less than those of BOLT-LMM, currently the most powerful MMAA tool. When applied to real phenotypes, LDAK-KVIK has the highest power of all tools considered. For example, across the 40 quantitative UK Biobank phenotypes (average sample size 349k), LDAK-KVIK finds 16% more independent, genome-wide significant loci than classical linear regression, whereas BOLT-LMM and REGENIE find 15% and 11% more, respectively. LDAK-KVIK can also perform gene-based tests; across the 40 quantitative UK Biobank phenotypes, LDAK-KVIK finds 18% more significant genes than the leading existing tool. Lastly, LDAK-KVIK produces state-of-the-art polygenic scores.

## INTRODUCTION

Genome-wide association studies (GWAS) have greatly advanced our understanding of the genetic basis underlying complex diseases, offering valuable insights into disease mechanisms and potential therapeutic targets.^1,2^ Since the first GWAS was carried out in 2005, sample sizes have regularly increased, such that studies involving over 100,000 individuals are now common.^3–7^ Initially, GWAS relied on classical linear or logistic regression to test for association between single nucleotide polymorphisms (SNPs) and phenotypes. However, in recent years, mixed-model association analysis (MMAA) has become the method of choice.^8,9^ MMAA can reduce false positives by accounting for cryptic relatedness, and increase true positives by factoring in the contributions of SNPs other than the one being tested.^10^

The most effective MMAA methods are two-step.^11–14^ In Step 1, they construct leave-one-chromosome-out (LOCO) polygenic scores (PGS) and estimate *λ*, a test statistic scaling factor. In Step 2, they regress the phenotype on the SNPs, including the LOCO PGS as an offset, then scale the resulting test statistics. The computational demands of a two-step MMAA tool depend mainly on its algorithmic design, whereas its power depends primarily on the accuracy of its LOCO PGS.^15,16^ For example, REGENIE^13^ tends to be faster than BOLT-LMM^12^ because its algorithm for estimating SNP effect sizes is block-based instead of genome-wide. However, BOLT-LMM tends to detect more significant associations than REGENIE because it usually constructs more accurate LOCO PGS.

In this paper, we introduce LDAK-KVIK, a novel two-step MMAA tool for analyzing quantitative and binary phenotypes. We first use simulated phenotypes to show that LDAK-KVIK controls type 1 error for both homogeneous and heterogeneous data sets, and also when analyzing highly imbalanced phenotypes (e.g., diseases with very few cases). We then apply LDAK-KVIK to real phenotypes from the UK Biobank.^17,18^ When used for single-SNP association analysis, LDAK-KVIK finds more significant associations than BOLT-LMM,^12^ REGENIE,^13^ fastGWA^19^ and GCTA-LOCO^10^ four of the leading existing MMAA tools. Meanwhile, when used for gene-based association analysis, LDAK-KVIK finds more significant associations than LDAK-GBAT,^20^ the leading existing tool for gene-based association testing.

## RESULTS

### Overview of LDAK-KVIK

A detailed description of LDAK-KVIK is provided in the **Online Methods** and **Supplementary Notes 1-5**. Here we highlight its key features.

LDAK-KVIK is a computationally efficient MMAA tool. Firstly, it never needs to store genotypes for more than 512 SNPs, and so has very low memory demands. Secondly, we have developed a chunk-based variational Bayes solver (illustrated in **Supplementary Figure 1**) that requires 5-20 times fewer updates than conventional variational Bayes solvers (**Supplementary Figure 2**).^12,21^ Our solver not only estimates SNP effect sizes (required when constructing the Step 1 PGS), but also efficiently computes terms of the form V^−1^A, where V is phenotypic variance matrix and A is a vector of genotypes or phenotypes (required when estimating heritability). Thirdly, we have developed a fast empirical algorithm for computing the saddlepoint approximation (SPA).^22–24^ Fourthly, LDAK-KVIK can analyze multiple phenotypes simultaneously.

LDAK-KVIK increases detection power, relative to existing MMAA tools, by using more realistic models for how SNP effect sizes vary across the genome.^25,26^ Firstly, LDAK-KVIK models how per-SNP heritability depends on minor allele frequency (MAF), whereas existing MMAA tools typically assume that per-SNP heritability is constant.^27,28^ Secondly, LDAK-KVIK uses a mixture prior distribution for SNP effect sizes that contains a Gaussian and a Laplace distribution (we refer to this as an elastic net prior, because of its similarities to elastic net regularization^29^), whereas existing MMAA tools generally restrict to mixtures of Gaussian distributions.^10–14^

By default, LDAK-KVIK sets *λ*=1, which is the correct value when analyzing homogeneous data. However, setting *λ*=1 will result in deflated test statistics when analyzing heterogeneous datasets. Therefore, we have developed a novel test for structure, based on the average pairwise correlation between 512 randomly-picked SNPs (the correlation is calculated after regressing out covariates). If this test finds high structure, LDAK-KVIK sets *λ* so that the mean test statistic of weakly-associated SNPs matches that obtained if we were to replace the elastic net prior distribution for SNP effect sizes with an infinitesimal prior and estimate *λ* using the Grammar-Gamma formula.^11^

In addition to testing SNPs individually for association with the phenotype, LDAK-KVIK can also perform gene-based tests. This is achieved by providing the results of the single-SNP analysis to our existing software LDAK-GBAT. ^20^ To avoid confusion, we refer to our new tool as LDAK-KVIK when testing SNPs for association, and as LDAK-KVIK-GBAT when testing genes for association.

### Data

We use genotype and phenotype data from the UK Biobank.^17,18^ In total, there are 487,152 individuals, of which approximately 95% are of European ancestry. From this we construct the following six datasets. The “white dataset” contains 367,981 white British individuals. The “homogeneous dataset” contains 63,000 unrelated white British individuals (no pair of individuals is closer than second-cousins). The “family dataset” contains 63,000 of the most closely related white British individuals (including approximately 13,300 full-siblings, 2,600 parent-offspring pairs and 300 second-degree relatives). The “twins dataset” contains 63,000 white British twins (generated by duplicating the genotypes of 31,500 individuals from the homogeneous dataset). The “British-Irish dataset” contains 63,000 unrelated white individuals, of whom approximately 51,300 are British and 11,700 Irish. Lastly, the “multi-ancestry dataset” contains 63,000 individuals selected to maximize ethnic diversity (approximately 63%, 24% and 13% are European, African and Asian, respectively). After restricting to autosomal, biallelic SNPs with MAF>0.001, all datasets contain 690,264 SNPs, except for the multi-ancestry dataset which contains 639,727 SNPs.

We first analyze simulated phenotypes. When generating these, we randomly select causal SNPs from the start of each chromosome then use the chromosome ends as null SNPs for evaluating type 1 error (illustrated in **Supplementary Figure 3**). We subsequently analyze 40 quantitative and 20 binary phenotypes from the UK Biobank. The quantitative phenotypes are height, body mass index, 20 biochemistry measurements (e.g., cholesterol, c-reactive protein, and urate), 16 blood measurements (e.g., haemoglobin concentration, lymphocyte percentage and platelet count) and two urine measurements (levels of creatinine and sodium). The binary phenotypes are defined based on ICD-10 codes^30^ (e.g., hypertension, obesity, and asthma) and have prevalences ranging from 0.02% to 29%. Additional details of the data are provided in **Supplementary Note 6**.

For the analyses below, we always include ten principal components as covariates; when analyzing real phenotypes, we additionally include age, sex, age^2^ and age*sex. LDAK-KVIK determines that the white, homogeneous, British-Irish and multi-ancestry datasets have low structure (so sets *λ*=1), but that the family and twins datasets have high structure (so LDAK-KVIK estimates *λ*). While it may appear surprising that the multi-ancestry dataset is judged to have low structure, this reflects that we include principal components as covariates (if these are excluded, our test finds extremely high structure).

### Existing tools

For single-SNP association testing, we compare LDAK-KVIK with classical linear and logistic regression, and with five existing MMAA tools: BOLT-LMM,^12^ REGENIE,^13^ fastGWA^19^, GCTA-LOCO^10^ and SAIGE^14^ (a summary of each tool is provided in **Supplementary Note 7**). Note that REGENIE and fastGWA are designed for both quantitative and binary phenotypes, whereas BOLT-LMM and GCTA-LOCO are designed for quantitative phenotypes, while SAIGE is designed for binary phenotypes. For gene-based association testing, we compare LDAK-KVIK-GBAT with our existing tool LDAK-GBAT,^20^ which we previously found to be consistently more powerful than five alternative tools (including MAGMA ^31^ and fastBAT^32^).

### Heritability models

We consider heritability models of the form 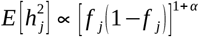, where 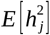 is the expected heritability contributed by SNP j, and fj is its MAF. When *α* =−1 all SNPs are expected to contribute equal heritability, whereas *α* >−1 (*α* <−1) implies that SNPs with higher MAF are expected to contribute more (less) heritability than SNPs with lower MAF. All five existing MMAA tools assume *α* =−1 throughout their operations (e.g., REGENIE assumes α=−1 when constructing the Step 1 PGS, while BOLT-LMM assumes α=−1 both when constructing the Step 1 PGS and when estimating λ). By contrast, LDAK-KVIK estimates α from the data.

### LDAK-KVIK controls type 1 error

We simulate quantitative and binary phenotypes for the homogeneous, family and twins datasets. For each dataset, we consider 12 different scenarios, obtained by varying the heritability (0.2 or 0.5), the number of causal SNPs (5k or 20k), and for binary phenotypes, also the prevalence (10% or 1%). When generating causal SNP effect sizes, we assume *α* =−1 (all causal SNPs are expected to contribute equal heritability). We perform single-SNP analysis using LDAK-KVIK, then measure type 1 error based on the average χ^2^(1) test statistic of null SNPs, and the proportions of null SNPs with p-values below 0.05, 0.001 and 5×10^−5^. **Figure 1** and **Supplementary Figure 4** show that LDAK-KVIK controls type 1 error for all 12 scenarios across all three datasets. This remains the case when we instead perform a gene-based analysis (**Supplementary Figure 5**).

**Figure 1.**
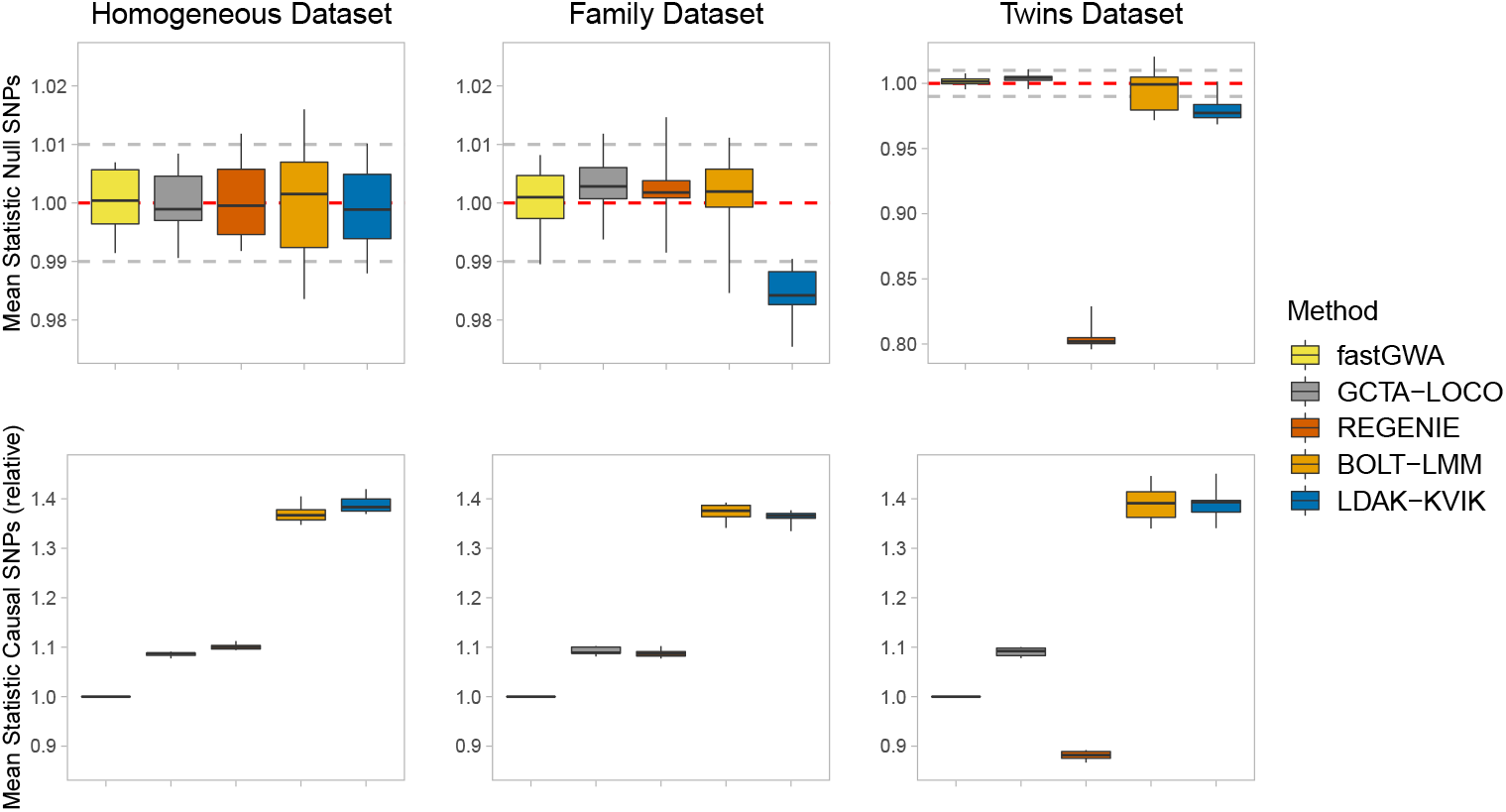
Type 1 error and power of MMAA tools for quantitative phenotypes. We generate ten quantitative phenotypes for each of the homogeneous, family and twins datasets (each dataset contains 63k individuals). Each phenotype has heritability 0.5 and 5k causal SNPs, with causal SNP effect sizes sampled assuming α =−1. We perform single-SNP association analysis using BOLT-LMM, REGENIE, fastGWA, GCTA-LOCO and LDAK-KVIK. Panels report the mean χ^2^(1) test statistic of null SNPs (top) and causal SNPs (bottom) for each replicate. The three horizontal lines in each box mark the median and inter-quartile range. Note that in the top panels, the horizontal solid and dashed lines mark, respectively, the expected value if a tool is well-calibrated and approximate 95% confidence intervals, while in the bottom panels, values are reported relative to the results from fastGWA.

For comparison, **Supplementary Figures 6-11** provide results from analyzing the simulated phenotypes using the existing MMAA tools BOLT-LMM, REGENIE, fastGWA, GCTA-LOCO and SAIGE. In general, each of the five tools controls type 1 error for all scenarios considered, except that REGENIE can produce inflated test statistics for common binary phenotypes (e.g., when analyzing the 40 binary phenotypes with prevalence 10% for the homogeneous dataset, the average *χ*^2^(1) test statistic is 1.03), while SAIGE can produce inflated test statistics for rare binary phenotypes when there is high structure (e.g., when analyzing the 20 binary phenotypes with prevalence 1% and heritability 0.5 for the twins dataset, the average *χ*^2^(1) test statistic is 1.10).

For **Supplementary Figure 12** we switch to the British-Irish dataset, showing that LDAK-KVIK continues to control type 1 error when applied to phenotypes with fine-scale population structure (specifically, phenotypes where citizenship explains 20% of the total variance). Meanwhile, **Supplementary Figures 13 & 14** report results for the multi-ancestry dataset. These show that LDAK-KVIK also controls type 1 error when applied to phenotypes with ancestry-specific genetic architectures (e.g., if we generate phenotypes where either the causal SNPs or assumed value of *α* vary across ancestries^33^).

### LDAK-KVIK is computationally efficient

Table 1 compares the runtime and memory requirements of LDAK-KVIK with existing MMAA tools when testing for association 690k directly-genotyped SNPs, using either the homogeneous or white datasets (63k and 368k individuals, respectively). For quantitative phenotypes, LDAK-KVIK and REGENIE have similar demands, typically requiring 5-10 CPU hours and less than 16 Gb memory to analyze phenotypes for the white dataset. fastGWA is the most computationally-efficient tool, but only if we ignore the one-off cost for constructing the genomic relationship matrix (which for the white dataset, took approximately 760 CPU hours). By contrast, BOLT-LMM and GCTA-LOCO are substantially more demanding. For example, BOLT-LMM takes over 100 CPU hours to analyze phenotypes for the white dataset, while GCTA-LOCO takes over 400 CPU hours just to analyse phenotypes for the homogeneous dataset (and thus it is not feasible for us to use GCTA-LOCO for the white dataset). The computational demands of LDAK-KVIK are similar when we switch from quantitative to binary phenotypes (which is also the case for REGENIE and fastGWA).

The runtimes in **Table 1** come from analyzing datasets with low structure. When applied to datasets with high structure, LDAK-KVIK is approximately 30% slower when the phenotype is quantitative, but approximately 70% faster when the phenotype is binary (e.g., if we force LDAK-KVIK to treat the white dataset as if it has high structure, the average runtime for quantitative phenotypes increases from 8 to 10 CPU hours, while the average runtime for binary phenotypes reduces from 10 to 3 CPU hours).

**Table 1.** Computational requirements of MMAA tools. We perform GWAS for five quantitative phenotypes (glucose, glycated haemoglobin, haemoglobin concentration, height and high-density lipoprotein) and five binary phenotypes (asthma, atrial fibrillation, ischaemic heart disease, dental caries and haemorrhoidal skin tags). Each GWAS analyzes 690k SNPs using either the homogeneous or white dataset, which contain 63k and 368k individuals, respectively (note that after excluding individuals with missing phenotypes, the average sample sizes are 59k and 343k, respectively). All analyses were performed using one core of an AMD EPYC Genoa 9654 CPU processor. Values report the mean runtime and memory usage across either the five quantitative or five binary phenotypes. We report two sets of values for fastGWA, depending on whether we exclude or include the computation of the genomic relatedness matrix (which only needs to be done once per dataset). It was not feasible for us to analyze phenotypes for the white dataset using GCTA-LOCO (we estimate each analysis would take over 10,000 hours and require over 500Gb memory).

| Phenotypes | MMAA Tool | Homogeneous Dataset (63k individuals) |  | White Dataset (368k individuals) |  |
| --- | --- | --- | --- | --- | --- |
|  |  | CPU Hours | Memory (Gb) | CPU Hours | Memory (Gb) |
| Quantitative | BOLT-LMM | 18 | 11 | 110 | 61 |
|  | REGENIE | 1.3 | 3 | 5.9 | 16 |
|  | fastGWA | 0.02 (24) | 1 (1) | 0.2 (760) | 3 (3) |
|  | GCTA-LOCO | 480 | 170 | Not Feasible |  |
|  | LDAK-KVIK | 0.9 | 2 | 7.7 | 5 |
| Binary | REGENIE | 1.8 | 3 | 10 | 16 |
|  | fastGWA | 0.1 (24) | 1 (1) | 1.2 (760) | 3 (3) |
|  | SAIGE | 32 | 11 | 230 | 61 |
|  | LDAK-KVIK | 0.9 | 2 | 10 | 5 |

**Supplementary Figure 15** and **Supplementary Table 1** provide further details of the LDAK-KVIK runtimes and results from additional analyses. For example, we see that LDAK-KVIK spends approximately 95% of its runtime in Step 1. This means that if we increase the number of association analysis SNPs from 690k to 9.0M, mimicking the situation where we perform a GWAS using imputed data, the impact on runtime is relatively modest (e.g., the average time to analyze quantitative phenotypes for the white dataset increases from 8 to 10 CPU hours). Meanwhile, we see that performing the gene-based association analysis (i.e., running LDAK-KVIK-GBAT as well as LDAK-KVIK) adds less than 5% to the runtime.

**Supplementary Tables 2 & 3** show that the runtime of LDAK-KVIK can be reduced by using multiple processors and/or analyzing multiple phenotypes simultaneously. For example, using two, four and eight CPUs reduces the runtime by approximately a quarter, a half and two-thirds, respectively, while analyzing five phenotypes simultaneously is about 60% faster than analyzing the same five phenotypes separately. **Supplementary Figure 16** shows that both the runtime and memory requirements of LDAK-KVIK are approximately linear in the number of individuals, and therefore, it is feasible to apply LDAK-KVIK to very large datasets (e.g., it took on average 25 CPU hours to analyze a dataset containing 1M individuals).

### LDAK-KVIK is powerful

Figure 1 and **Supplementary Figures 17-19** compare the power of MMAA tools when performing single-SNP analysis of the simulated phenotypes. For the quantitative phenotypes, LDAK-KVIK and BOLT-LMM are consistently the two most powerful MMAA tools, substantially ahead of REGENIE and GCTA-LOCO, while fastGWA generally has lowest power. For the binary phenotypes, the four MMAA tools have very similar power, reflecting that it is challenging to construct accurate PGS for phenotypes with low (observed-scale) heritability. **Supplementary Figure 20** shows that for gene-based analysis, LDAK-KVIK-GBAT tends to be substantially more powerful than LDAK-GBAT for quantitative phenotypes, while the two tools have similar power for binary phenotypes.

### The benefit of modeling the relationship between per-SNP heritability and MAF

We now generate phenotypes using values of *α* between −1 and 0, and compare the performance of the default version of LDAK-KVIK (which infers *α* from the data), to an alternative version that forces *α* =−1 (mirroring the assumptions of the existing two-step MMAA tools). **Figure 2** shows that the advantage of the default version over the alternative version increases with *α*, which reflects that the former is able to estimate *α* with reasonable accuracy, and use this information to construct more accurate LOCO PGS. These results are relevant because studies of real human traits typically find *α* ≫−1.^25,27,28,34,35^ For example, **Supplementary Figure 21** shows that across the 40 quantitative UK Biobank phenotypes, the mean estimate of *α* is −0.23 (s.d. 0.005), with *α* significantly greater than −1 for all phenotypes (P<0.01).

**Figure 2.**
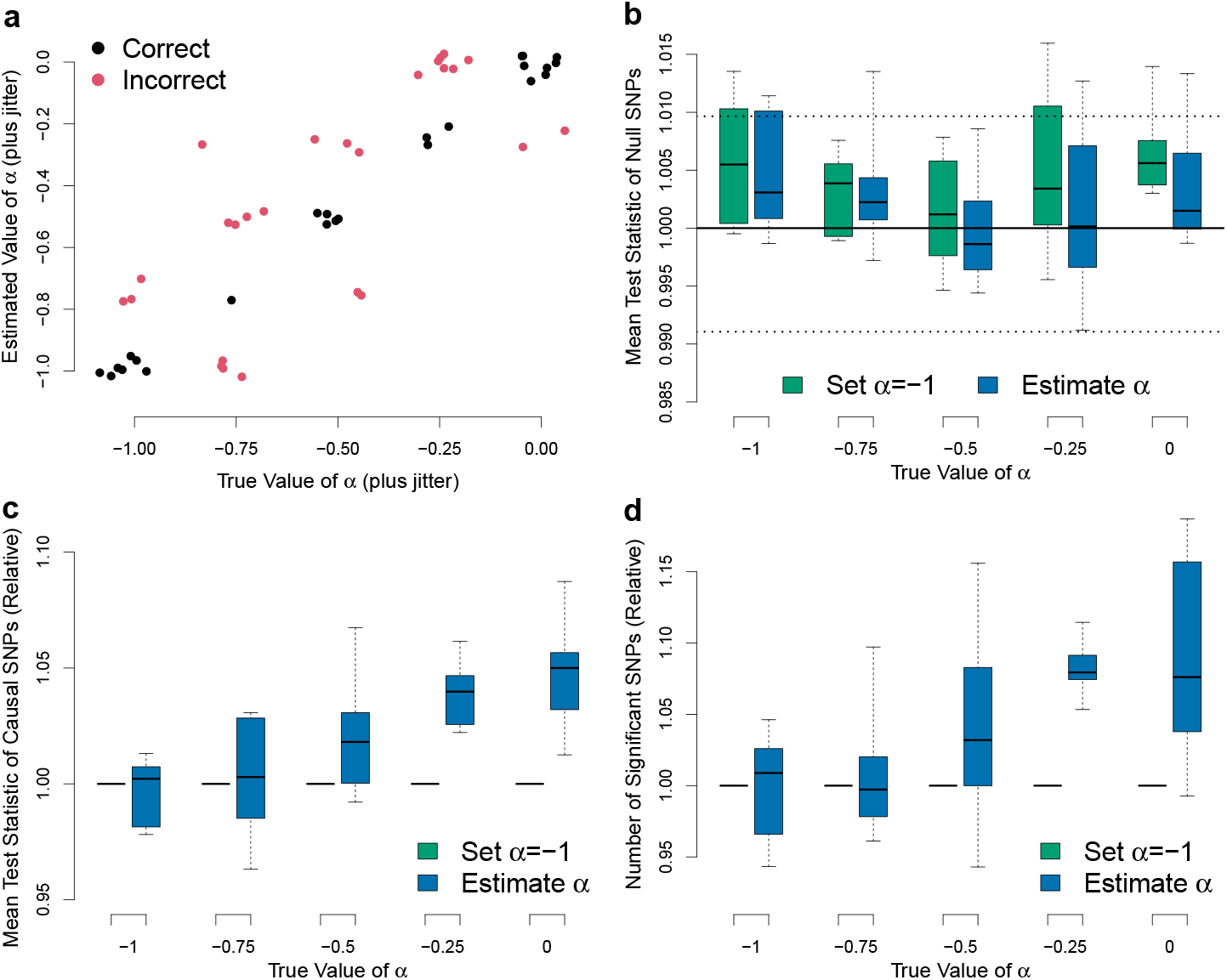
Exploiting the dependency of per-SNP heritability on MAF. We generate sets of ten quantitative phenotypes for the homogeneous dataset (63k individuals). Each phenotype has heritability 0.5 and 20k causal SNPs, with causal SNP effect sizes sampled assuming values of *α* ranging from −1 to 0. We analyze the phenotypes using the default version of LDAK-KVIK (which estimates *α* from the data), and an alternative version that sets *α* =−1. **a**, Points compares estimates of *α* from the default version of LDAK-KVIK to the true values, with colors indicating whether the estimate is correct (black) or incorrect (red). **b**, Boxes report the mean χ^2^(1) test statistic of null SNPs; the horizontal solid and dashed lines mark, respectively, the expected value if a tool is well-calibrated and approximate 95% confidence intervals. **c**, Boxes report the mean χ^2^(1) test statistic of causal SNPs. **d**, Boxes report the number of significant SNPs (P<5×10^−8^). Note that the three horizontal lines in each box mark the median and inter-quartile range, while values in the bottom panels are reported relative to the results from the alternative version of LDAK-KVIK.

### Single-SNP analysis of UK Biobank phenotypes

Figure 3 compares the number of associations found by BOLT-LMM, REGENIE, fastGWA, BOLT-LMM-inf and LDAK-KVIK when analyzing the 40 quantitative phenotypes (note that BOLT-LMM-inf is a version of BOLT-LMM that assumes the same model as GCTA-LOCO, but that can feasibly be applied to the white dataset). LDAK-KVIK identifies 17.9% more independent, genome-wide significant loci than fastGWA (SNPs with P<5×10^−8^, filtered so that no pair within 1Mb has squared correlation above 0.1), which is slightly higher than BOLT-LMM (17.6%), and substantially higher than REGENIE (13.5%) and BOLT-LMM-inf (12.6%). **Supplementary Figure 22** shows that ranking of MMAA tools is the same if we instead measure power based on the mean *χ*^2^(1) test statistic of SNPs that are genome-wide significant from fastGWA.

**Figure 3.**
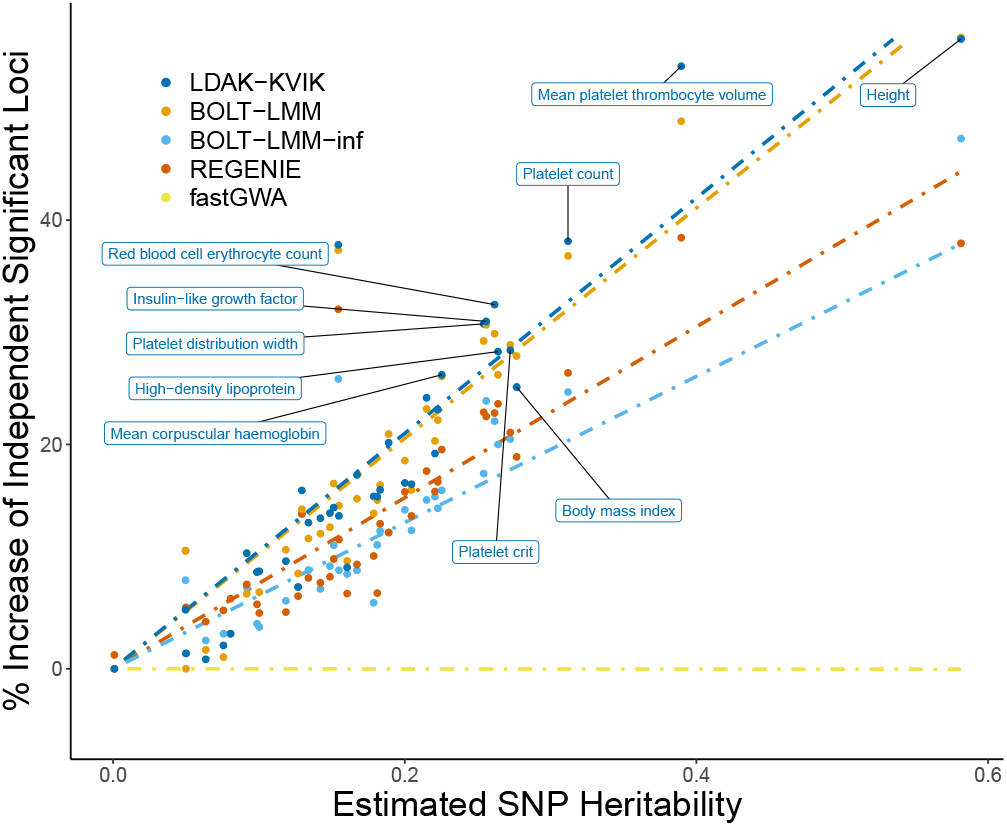
Performance of MMAA tools when analyzing 40 quantitative UK Biobank phenotypes. We analyze each phenotype using BOLT-LMM, REGENIE, fastGWA, BOLT-LMM-inf and LDAK-KVIK, then count the number of independent, genome-wide significant loci (SNPs with P<5×10^−8^, filtered so that no pair within 1Mb has squared correlation above 0.1). Points compare the number of loci found by each tool, relative to the results from fastGWA, with the estimated SNP heritability (obtained using our software SumHer^28^). The ten phenotypes with highest SNP heritability are named, while the dashed lines are obtained by regressing the relative numbers of loci found by each tool on the estimates of SNP heritability.

**Supplementary Figure 23** shows that when analyzing the 20 binary phenotypes, all four MMAA tools perform similarly. For example, REGENIE, fastGWA, SAIGE and LDAK-KVIK in total find 676, 649, 612 and 686 independent, genome-wide significant loci, respectively (for comparison, classical logistic regression finds 667). These results are consistent with those when analyzing the simulated phenotypes, and again reflect the difficulty of constructing accurate PGS for binary phenotypes.

### Accuracy of Step 1 PGS

**Figure 4a** compares the accuracy of the Step 1 PGS constructed by LDAK-KVIK and BOLT-LMM, for each of the 40 quantitative UK Biobank phenotypes. For comparison, we also consider ridge regression PGS, which are constructed using similar assumptions to the PGS constructed by REGENIE (the latter does not report SNP effect sizes, so we can not measure the accuracy of its PGS directly). As expected, the PGS accuracies mirror the detection powers of the three MMAA tools (**Figure 3**). For example, the PGS from LDAK-KVIK tend to be slightly more accurate than the PGS from BOLT-LMM and substantially more accurate than the PGS from ridge regression, explaining why LDAK-KVIK found slightly more significant loci than BOLT-LMM and substantially more significant loci than REGENIE. **Supplementary Figure 24** shows that for the 20 binary phenotypes, the Step 1 PGS tend to have very low accuracy, explaining why the MMAA tools performed similarly to classical logistic regression.

**Figure 4.**
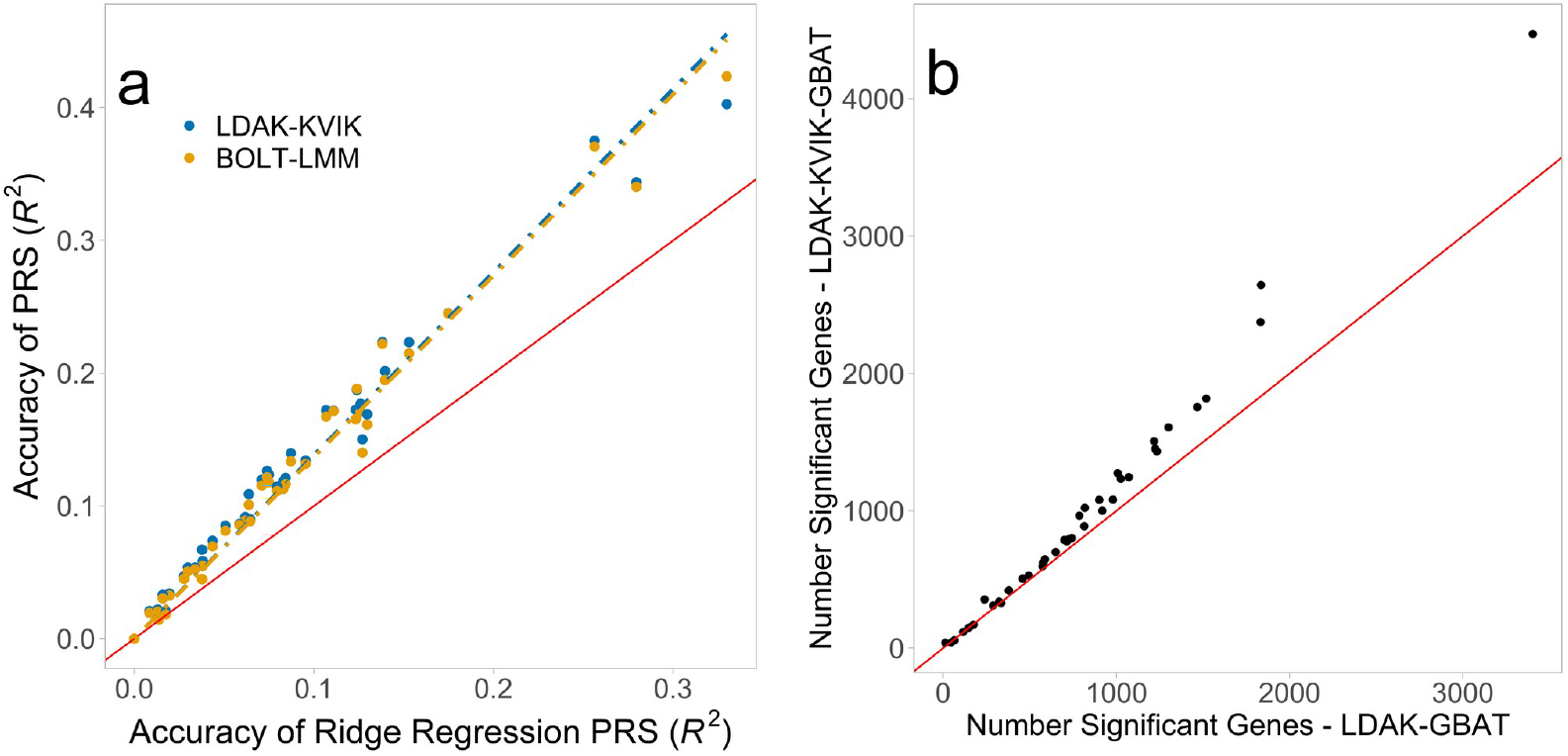
Accuracy of Step 1 PGS and power of LDAK-KVIK-GBAT. **a**, We applied BOLT-LMM and LDAK-KVIK to the 40 quantitative UK Biobank phenotypes. The y-axis reports the accuracy of the Step 1 PGS from each tool, measured by the squared correlation between predicted and observed phenotypes across 41k samples (distinct from those used for the association analysis). For comparison, the x-axis reports the accuracy of ridge regression PGS, which are constructed using similar assumptions to the REGENIE PGS. **b**, We applied LDAK-GBAT and LDAK-KVIK-GBAT to the 40 quantitative UK Biobank phenotypes; points compare the numbers of significant genes (P<0.05/17,322=2.9×10^−6^) found by each tool. In each panel, the diagonal line marks y=x.

### Gene-based analysis of UK Biobank phenotypes

We test 17,332 genes for association, defined based on RefSeq annotations.^36^ **Figure 4b** and **Supplementary Figure 25** report the number of significant genes (P<0.05/17,322=2.9×10^−6^) from LDAK-GBAT and LDAK-KVIK-GBAT. Across the 40 quantitative phenotypes, LDAK-KVIK-GBAT finds on average 18.4% more significant genes than LDAK-GBAT. By contrast, across the 20 binary phenotypes, there is no advantage using LDAK-KVIK-GBAT instead of LDAK-GBAT (in total, the tools find 1508 and 1515 significant genes, respectively).

### Enlarged analysis of 62 quantitative phenotypes

For **Supplementary Figure 26**, we increase the number of quantitative phenotypes from 40 to 62 (by adding 22 traits analyzed by the authors of fastGWA^19^), increase the total sample size from 368k to 459k (the latter represents all European UKBB individuals), and increase the number of SNPs from 690k to 9.0M (by including imputed variants). Across all 62 phenotypes, LDAK-KVIK and REGENIE find 13.3% and 9.0% more independent, genome-wide significant loci than fastGWA (if we restrict to the 22 new phenotypes, the values are 7.6% and 4.0%, respectively). Summary statistics and PGS SNP weightings for all 62 phenotypes are available at www.ldak-kvik.com/summaries.

### Using LDAK-KVIK on the UK Biobank Research Analysis Platform

The LDAK-KVIK website (www.ldak-kvik.com) provides guidelines for analyzing UK Biobank data on the Research Analysis Platform (RAP). For example, it explains how to extract phenotypes and covariates, and how to prepare the genotypes for Steps 1 & 2 (for Step 1, we recommend restricting to directly-genotyped SNPs with MAF>0.01 and call-rate above 90%, while for Step 2, we recommend converting the BGEN files into PLINK BED format). Note that while LDAK-KVIK can perform quality control on-the-fly (e.g., exclude SNPs based on MAF, call-rate, variance and/or information score), we generally recommend doing this when preparing the genotypes (because it usually only needs to be performed once, regardless of the number of phenotypes being tested). Similarly, while LDAK-KVIK is able to analyze multiple phenotypes simultaneously, we generally recommend analyzing phenotypes individually. This is because we have found that when using the RAP (or other high-performance clusters), it is usually more efficient, in terms of cost, time to complete all analyses and ease of debugging, to run many low-resource jobs instead of fewer high-resource jobs (e.g., we try to restrict to jobs that only use 1-4 cores, require less than 8Gb memory, and take less than ten hours to complete).

## DISCUSSION

We have presented LDAK-KVIK, a novel tool for performing single-SNP and gene-based mixed-model association analysis. We have shown that LDAK-KVIK can be validly applied to homogeneous and heterogeneous datasets, and to both quantitative and binary phenotypes. LDAK-KVIK is computationally efficient, able to analyse data for 100,000s of individuals within a few hours, and has low memory requirements. Furthermore, LDAK-KVIK is powerful; for example, when used for single-SNP analysis of quantitative phenotypes, LDAK-KVIK consistently finds more significant associations than the existing MMAA tools BOLT-LMM, REGENIE, fastGWA and GCTA-LOCO.

Compared to BOLT-LMM, the main advantage of LDAK-KVIK is its computational efficiency (e.g., when analyzing the UK Biobank phenotypes, LDAK-KVIK was at least ten times faster and required five times less memory). This is mainly due to the development of a chunk-based variational Bayes solver. **Supplementary Figure 2** shows that our variational Bayes solver can construct PGS 5-10 times faster than the standard (genome-wide) variational Bayes solver, and can compute terms of the form V^−1^A ten times faster than conjugate gradient descent. Furthermore, our variational Bayes solver has a small memory footprint (typically less than 1Gb), because it never needs to store data for more than 256 SNPs at a time.

Compared to REGENIE, the main advantage of LDAK-KVIK is its power (e.g., across the 40 quantitative UK Biobank phenotypes, LDAK-KVIK found 3.7% (s.d. 0.6%) more independent, genome-wide significant loci, and when restricted to SNPs significant from fastGWA, the test statistics from LDAK-KVIK were on average 2.7% (s.d. 0.3%) higher than those from REGENIE). This power increase mainly reflects that LDAK-KVIK uses more realistic models for the genetic architecture of complex traits. Specifically, LDAK-KVIK models the relationship between per-SNP heritabilities and MAF, and uses an elastic net prior distribution for SNP effect sizes. As shown in **Figure 4a**, these features result in LDAK-KVIK constructing more accurate PGS in Step 1, which then increases the chance of discovering associations in Step 2.

We recognize that LDAK-KVIK can be considered much slower than fastGWA. However, there are four points to note. Firstly, LDAK-KVIK is only slower if we ignore the time fastGWA takes to compute the genomic relatedness matrix. Although a one-off cost, this can be non-trivial (e.g., when analyzing 368k individuals, this step took approximately 760 CPU hours, which is longer than the total time LDAK-KVIK required to analyse all 40 quantitative phenoptypes). Secondly, LDAK-KVIK is usually substantially more powerful than fastGWA, reflecting that fastGWA focuses on controlling type 1 error, instead of improving power relative to classical linear regression. Thirdly, fastGWA is only designed for homogeneous data, or data containing related individuals, whereas LDAK-KVIK can also be applied to datasets including individuals from multiple ancestries. Fourthly, LDAK-KVIK includes an option to perform an approximate version of fastGWA, that uses a much faster algorithm for identifying related pairs of individuals. **Supplementary Figure 27** shows that when applied to the 60 quantitative and binary UK Biobank phenotypes, our approximate version of fastGWA gives results very similar to those from the original version of fastGWA, yet on average takes under two CPU hours to analyze each trait.

We realise that LDAK-KVIK has some limitations. Firstly, our method for calculating *λ* is approximate, and can result in deflated test statistics (evident from the results for the family and twins datasets in Figure 1, with further details in **Supplementary Figure 28**). Therefore, LDAK-KVIK would become more powerful if an exact solution for estimating *λ* was found (in **Supplementary Note 8** we summarize eight different approaches we tried for estimating *λ*). We note that when analyzing datasets with high structure, our strategy for estimating *λ* has similarities to that used by BOLT-LMM (both tools compare test statistics to those obtained using LOCO ridge regression PGS). However, the key difference is that LDAK-KVIK estimates *λ* based on the average test statistics of weakly-associated SNPs, whereas BOLT-LMM uses the estimated intersect from LD Score Regression (LDSC). **Figure 1** indicates that the BOLT-LMM approach can be advantageous (e.g., it avoids the deflation observed for the twins dataset). However, we prefer our strategy for two main reasons. Firstly, the approach used by BOLT-LMM assumes *α* =−1, and can result in substantial inflation when this assumption is inappropriate (e.g., for **Supplementary Figure 29**, we generate quantitative phenotypes for 368k individuals assuming *α* =−0.25, and show that the test statistics from BOLT-LMM are inflated by on average 9%). Secondly, when running LDSC, it is necessary to make assumptions regarding the location of causal variants (as this determines how to compute LD scores), but it is hard to assess the validity of these assumptions. For example, **Supplementary Figure 30** shows that when analyzing the phenotype height, switching from external LD scores to in-sample LD scores resulted in the estimate of *λ* increasing from 1.01 to 1.10.

A second limitation of LDAK-KVIK is that it estimates *α* using a grid search, with five pre-defined values (−1, −0.75, −0.5, −0.25 and 0). While the use of pre-defined values enables high computational efficiency (because LDAK-KVIK can evaluate model fit for all five values simultaneously), we appreciate that it limits the accuracy of the algorithm. For example, when analyzing the 50 phenotypes underlying **Figure 2**, for which the true values of *α* were always among the five pre-defined values, LDAK-KVIK only estimated *α* correctly 24 times. However, while imperfect, we believe that our algorithm is a marked improvement on the status quo, which is to simply assume *α*=−1.

Thirdly, we found that when analyzing binary phenotypes, LDAK-KVIK was often only slightly more powerful than classical logistic regression. We note that this was the same for other MMAA tools. Furthermore, it partially reflects that we have focused on non-ascertained phenotypes (e.g., the UK Biobank is a population-based sample, so the proportion of cases for each ICD10 disease will be close to the disease’s prevalence). For **Supplementary Figure 31**, we simulate ascertained binary phenotypes. We find that LDAK-KVIK continues to have good control of type 1 error, however its power advantage over classical logistic regression depends on the scenario considered (e.g., its advantage generally increases when analyzing more common phenotypes, but reduces when analyzing rarer phenotypes).

A fourth limitation concerns the analysis of phenotypes for which a substantial proportion of variation is explained by common environment. Previous work by the authors of fastGWA^19^ has shown that MMAA tools that utilize SNP-based PGS (e.g., BOLT-LMM, REGENIE and LDAK-KVIK), can produce inflated test statistics when applied to phenotypes with a strong environmental effect. As shown in **Supplementary Figure 32**, this inflation can potentially be avoided by using pedigree-based PGS (i.e., by using either fastGWA, or the approximate version of fastGWA contained within LDAK-KVIK).

Fifthly, we note that although LDAK-KVIK is able to analyze multiple phenotypes simultaneously, and doing so can substantially reduce total runtime, the speedups are less than those achieved by REGENIE (**Supplementary Table 3**). Therefore, while LDAK-KVIK and REGENIE have similar runtimes when analyzing a small number of phenotype (e.g., less than five), REGENIE can be much faster than LDAK-KVIK when analyzing many phenotypes (e.g., more than ten). Saying this, we hope that the increased detection power of LDAK-KVIK, relative to REGENIE, will compensate for the increased running time when analyzing many phenotypes,

Our simulations indicate that LDAK-KVIK can be validly applied to multi-ancestry datasets. Specifically, we found that LDAK-KVIK controls type 1 error both for phenotypes whose genetic architectures are the same across ancestries, and for phenotypes whose architectures are ancestry-specific^33^ (**Supplementary Figures 13 & 14**). For **Supplementary Figure 33**, we compare results from analyzing the 40 quantitative UK Biobank phenotypes using either the full dataset (487k individuals) or only the European individuals (462k individuals). We find there is limited benefit including the non-European individuals (e.g., the number of independent, genome-wide significant loci increases for 23 phenotypes, and decreases for 17 phenotypes). However, this likely reflects that almost all (95%) of the UK Biobank individuals are European, and so it would be interesting to investigate the impact on power for more diverse datasets.

We note that it is possible to speed up LDAK-KVIK, and the other two-step MMAA tools, by reducing the number of SNPs used in Step 1 (e.g., when analyzing UK Biobank data, the authors of SAIGE used only 94k SNPs when constructing the PGS). **Supplementary Figure 34** shows that when analyzing simulated phenotypes for the homogeneous, family and twins datasets, reducing the number of Step 1 SNPs from 690k to either 477k or 267k leads to a noticeable reduction in detection power, and thus we advise against this approach.

We finish by pointing out that although designed for association testing, LDAK-KVIK also produces state-of-the-art PGS (in addition to constructing the LOCO Step 1 PGS, it also constructs a genome-wide PGS, and reports the corresponding SNP effect sizes). Previous works have shown that BOLT-LMM is one of the leading tools for constructing PGS.^15,26^ Here we have found that LDAK-KVIK tends to produce more accurate PGS than BOLT-LMM, and has substantially lower computational demands.

## DATA AVAILABILITY

Our study used data from UK Biobank, which we applied for and downloaded from www.ukbiobank.ac.uk. The UK Biobank has ethics approval from the North West Multi-centre Research Ethics Committee (MREC).

## CODE AVAILABILITY

LDAK-KVIK is part of the software package LDAK, which can be downloaded from www.dougspeed.com. Full instructions for running LDAK-KVIK are provided at www.ldak-kvik.com.

## ACKNOWLEDGMENTS

This research has been conducted using the UK Biobank Resource under Application Number 21432. The computing for this project was performed on the GenomeDK cluster (Aarhus University). We thank Anders Halager and Dan Søndergaard (Aarhus University) for programming suggestions, David Balding (University of Melbourne) for helpful comments on the manuscript, Jakub Bajzik, Al Depope and Matthew Robinson (Institute of Science and Technology Austria) for assistance with the UK Biobank Research Analysis Platform, Hamed Heydari (University of Toronto) for advice on heritability estimation, and Soumeen Jin (Karolinska Institute) for testing LDAK-KVIK. DS is supported by the Aarhus University Research Foundation (AUFF), by the Independent Research Fund Denmark (project no. 7025-00094B) and by a European Research Council Consolidator Grant (ID 101088901, acronym ClassifyDiseases).

## AUTHOR CONTRIBUTIONS

JH and DS jointly developed the software, performed the analysis and wrote the manuscript.

## ONLINE METHODS

Here we provide a concise description of LDAK-KVIK; please see **Supplementary Notes 1-7** for an exhaustive version, as well as details of existing MMAA tools and the UK Biobank data. Note that when describing LDAK-KVIK, we first assume the phenotype is quantitative, then later explain the changes required when the phenotype is binary.

### Notation

Suppose there are n individuals, each genotyped for m SNPs, recorded for q covariates and measured for a phenotype. Let the (n x m) matrix X’ contain the genotypes, let the length-n vector Y’ contain the phenotypes, and let the (n x q) matrix Z contain covariates. We use C to denote the total number of chromosomes, use X and Y to denote, respectively, the genotypes and phenotypes after regressing out the covariates, and use λ to denote the test statistic scaling factor. Without loss of generality, we assume X_j_ and Y are standardized to have mean zero and variance one.

## LDAK-KVIK Step 1

The main outcome of this step are C LOCO elastic net PGS, each taking the form 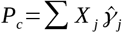, where 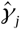 is the estimated effect size for SNP j, and the sum is across all SNPs not on Chromosome c. This step also estimates *λ* (described separately below).

LDAK-KVIK first computes models of the form 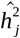, estimates of the heritability contributed by each SNP. For this, it considers heritability models of the form

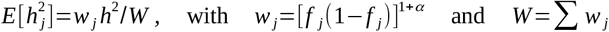

where fj is the MAF of SNP j, α determines how per-SNP heritability depends on MAF and h^2^ is the proportion of phenotypic variance explained by all SNPs. LDAK-KVIK estimates α and h^2^ using Randomized Haseman-Elston Regression^37^ and Monte Carlo restricted maximum likelihood ^12^ (REML), then sets 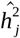 to its expected value given these estimates. Note that Monte Carlo REML must calculate terms of the form V^−1^Y, KV^−1^Y, V^−1^R and KV^−1^R, where V is an (n x n) variance matrix, K is an (n x n) genomic relatedness matrix (GRM), and R is a length-n vector whose elements are drawn from a standard Gaussian distribution; we calculate these terms using our novel Variational Bayes solver (described below).

LDAK-KVIK then constructs C LOCO elastic net PGS, for which it assumes^29^

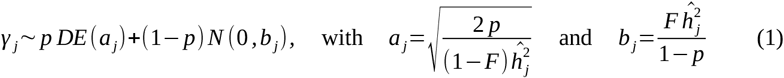

where DE(a_j_) denotes a double exponential distribution with rate a_j_, while N(0,b_j_) denotes a normal distribution with mean 0 and variance b_j_. The parameters p and F determine the relative contributions of the double exponential and normal distributions. LDAK-KVIK finds suitable values for these using cross-validation (by default, LDAK-KVIK uses 90% of samples to construct genome-wide PGS corresponding to ten different pairs of values for p and F, then picks the pair that has lowest mean-squared error measured using the remaining 10% of samples). Note that all PGS are constructed using our novel variational Bayes solver (described below).

## LDAK-KVIK Step 2

LDAK-KVIK tests SNP j for association using least-squares regression with either the model *E* [*Y* −*Pc*]= *X j β j* or *E* [*Y* −*Pc*]= *X j ‘β j* (note that the former uses residual genotypes, so is more accurate, whereas the latter uses raw genotypes, so is faster). By default, LDAK-KVIK copies the approach used by fastGWA^19^ and first tests all SNPs using the raw genotype model, then retests SNPs showing evidence of association (P<0.05) using the residual genotype model (if preferred, the user can instruct LDAK-KVIK to always use residual genotypes, or to always use raw genotypes). LDAK-KVIK then scales the resulting χ^2^(1) test statistics by λ.

### Novel test for structure

LDAK-KVIK picks 512 SNPs randomly from across the genome, then computes 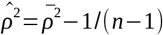, where 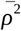 is the average squared correlation between the r pairs of SNPs on different chromosomes. To test 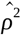 for significance, we use the fact that under the null hypothesis (i.e., if the data are homogeneous and so SNPs on different chromosomes are independent), each squared correlation is beta distributed with parameters 1/2 and (n-2)/2, and so 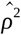 will have expectation 0 and variance 2(n-2)/[r(n-1)^2^(n+1)]. LDAK-KVIK determines there is high structure when 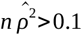 and the corresponding p-value is below 0.001 (otherwise, it determines the structure is low). The required calculations are very fast (e.g., when analyzing 368k individuals, they takes less than one minute) and have low memory demands (because it is only necessary to store genotypes for 512 SNPs). **Supplementary Figure 35** shows that our test for structure is robust to the choice of SNPs (e.g., its results change little if we reduce to 256 SNPs or restrict to SNPs with either high or low MAF).

Our test for structure is motivated by the fact that 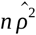 is an estimate of the maximum average inflation of test statistics when using classical linear regression. This is based on the following logic. Suppose that we could partition the genetic contribution to a phenotype as *G*=*L*_*j*_+ *D* _*j*_ where L_j_ and D_j_ are the components local and distal to SNP j, respectively, with heritability contributions h^2^_Dj_ and h^2^_Dj_. Then when testing SNP j for association with the phenotype, we could write the expected *χ*^2^(1) test statistic from classical linear regression as 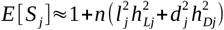, where 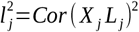 and 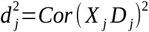 are the proportions of local and distal genetic variation tagged by SNP j, respectively. Under this partitioning, 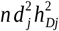 can be viewed as the expected inflation of S_j_ due to structure. Finally, if we assume that 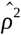 is a reasonable estimator of 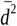, the average value of 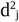 across all SNPs, and recognize that 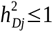, then it follows that 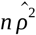 is an upper bound for the average inflation of test statistics due to structure. Please note that while 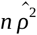 measures the potential inflation of test statistics when using the raw phenotype, it becomes a measure of deflation when using offset phenotypes (e.g., if 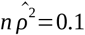, this indicates that test statistics from regressing Y on X_j_ will be on average up to 0.1 above their true values, but that test statistics from regressing Y-Pc on X_j_ will be on average up to 0.1 below their true values).

### Calculating the test statistic scaling factor

If the above test finds low structure, LDAK-KVIK sets *λ*=1. If it instead finds high structure, LDAK-KVIK estimates *λ* as follows. First it constructs C LOCO ridge regression PGS, for which it assumes 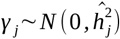. It then uses least-squares linear regression with the model *E* [*Y* −*Pc*]= *X j β j* to compute two sets of (unscaled) *χ*^2^(1) test statistics for 65,000 randomly-picked SNPs: for the first set (denoted by Uj), Pc is the LOCO elastic net PGS corresponding to SNP j, while for the second set (denoted by Tj), Pc is the corresponding LOCO ridge regression PGS. Next, LDAK-KVIK uses the Grammar-Gamma formula^11^ to compute *λ ‘*, the scaling factor corresponding to the second set of test statistics (by default it uses 20 randomly-picked SNPs). Finally, it sets 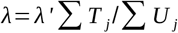, where the summations are across weakly-associated SNPs, which we define as those with 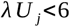 and 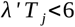 (because the definition of weakly-associated depends on *λ*, we use an iterative approach that typically requires 2-5 iterations).

Note that are three advantages to setting *λ*=1 for datasets with low structure (instead of estimating λ like we do for datasets with high structure). Firstly, it is more computationally efficient, because estimation of λ requires the construction of an extra set of LOCO PGS. Secondly, when the true value of λ is very close to one, the advantage of estimating λ is outweighed by the uncertainty of the estimate (e.g., estimates of λ for homogeneous datasets typically have standard error 0.002). Thirdly, we observe that our estimates of λ can be slightly conservative for homogeneous datasets (**Supplementary Figure 28**).

### Binary phenotypes

Many of the operations in LDAK-KVIK are based on a linear model of the form *Y i*=∑ *Xi, j* γ *j* +*ei*. In particular, this model is assumed (either explicitly or implicitly) when estimating h^2^, when constructing PGS and when testing SNPs for association. When the phenotype is quantitative, X_j_ and Y contain standardized residuals from linearly regressing X_j_’ and Y’, respectively, on Z, and LDAK-KVIK assumes 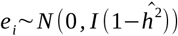, where I is an (n x n) identity matrix and 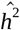 is the estimate of h^2^. When the phenotype is binary, LDAK-KVIK first computes the length-n vector μ *‘*, which contains estimates of the probabilities that each individual is a case given the covariates, and constructs the (n x n) diagonal matrix D, with *D*_*i,i*_∝μ′_*i*_(1−μ′_*i*_) and trace(D^−1^)=n. LDAK-KVIK then sets X_j_ to the residual from regressing X_j_’ on Z using weighted linear regression with weight matrix D, sets *Y* =*D*^−1^(*Y ‘* −μ *‘*), and assumes 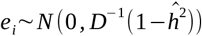. These changes are motivated by the observation that the estimated SNP effect size from regressing Y’ on both X_j_’ and Z using logistic regression is approximately equal to that from regressing Y on X_j_ using weighted linear regression with weight matrix D (see **Supplementary Note 5** for a proof). Note that ensuring trace(D^−1^)=n allows us to continue to treat h^2^ as a heritability (i.e., h^2^ continues to represent the proportion of variance of Y explained by all SNPs).

In addition to changing from standard to weighted linear regression, LDAK-KVIK makes two further modifications when analyzing binary phenotypes. Firstly, when it finds high structure, it switches to an approximate version of fastGWA (described below). This is motivated by the observation that MMAA tools tend to have very similar power when analyzing binary phenotypes, and particularly for datasets with high structure, so there is limited harm using a pedigree-based PGS instead of LOCO SNP-based PGS (but the former is much faster). The second modification affects Step 2. When analyzing quantitative phenotypes, LDAK-KVIK obtains the (unscaled) test statistics via a Wald Test (specifically, 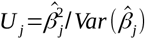, the square of the estimated effect size for SNP j divided by its estimated variance). When analyzing binary phenotypes, LDAK-KVIK first obtains Uj via a Wald Test, but if the corresponding p-value (after scaling by λ) is below 0.05, computes a revised test statistic using our empirical SPA solver (described below).

### Approximate version of fastGWA

The LDAK-KVIK version of fastGWA differs from the original version in four main ways. Firstly, whereas fastGWA constructs a GRM using all SNPs, our version uses only 1024 SNPs, picked semi-randomly from across the genome (specifically, LDAK-KVIK first randomly picks 5,120 SNPs, then retains the 1024 SNPs with highest variance). Secondly, when constructing the sparse GRM, fastGWA recommends truncating values below 0.05, whereas we truncate non-significant values (specifically, those with P>0.1/n, which typically corresponds to pairs of individuals with estimated relatedness below 0.15). Thirdly, we adjust estimates of relatedness for winners’ curse (**Supplementary Figure 36**).

Fourthly, when analyzing a binary phenotype, we assume the null model *Y* ~*N* (0, *σ*^2^ *K*_S_ + *E*^−1^), where K_S_ is the sparse GRM and E is a diagonal matrix with *E*_*i,i*_=μ′_*i*_(1−μ′ _*i*_). Note that this represents a simplified version of the quasi-likelihood used by fastGWA, because we set μ′, and therefore also E, based only on the covariates, whereas fastGWA updates μ′ to allow for the contribution of the sparse GRM. **Supplementary Figure 27** shows that despite these simplifications, our version of fastGWA performs similarly to the original version, both for quantitative and binary phenotypes.

### LDAK-KVIK-GBAT

Previously, we developed LDAK-GBAT, a tool for gene-based association analysis.^20^ This tests each gene using the mixed model 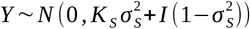, where K is a local GRM computed using only SNPs within the gene, and 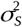 is the corresponding variance component. LDAK-GBAT finds the maximum likelihood estimate of 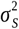, then obtains a p-value by testing if the estimate is greater than zero. Importantly, LDAK-GBAT requires only GWAS summary statistics and a reference panel. Therefore, LDAK-KVIK-GBAT applies LDAK-GBAT using the association results from Step 2, and using 5000 randomly-picked individuals from the data as an in-sample reference panel.

### Overview of Variational Bayes

This section explains the general idea of using variational Bayes to construct PGS, whereas the next section provides specific details of our novel variational Bayes solver. Please note that a detailed description of variational Bayes is provided in the supplementary material of the BOLT-LMM paper.^12^

When constructing PGS, LDAK-KVIK must compute *P*(γ∣*Y*), the posterior distribution of SNP effect sizes given the data. To do this, we first construct a model likelihood *L*(*Y*∣γ) by assuming that Y has the multivariate normal distribution 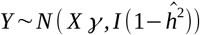, where I is an (n x n) identity matrix. We then specify π (γ), a prior distribution for SNP effect sizes, and use variational Bayes to approximate *P*(γ∣*Y*) as the product of independent, single-parameter posterior distributions (one for each SNP):

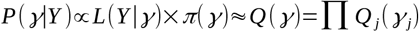

LDAK-KVIK updates *Q* (γ) one SNP at a time, by replacing the current *Q*_*j*_(γ _*j*_) with the distribution that minimizes the difference between the estimated and true posterior distributions (measured by the Kullback-Leibler divergence). Once convergence has been achieved, the corresponding PGS is constructed by setting γ *j*=η*j*, where η*j* is the expectation of *Q*_*j*_(γ _*j*_).

As explained above, Step 1 of LDAK-KVIK uses variational Bayes to construct elastic net PGS, assuming the prior distribution defined in Equation 1. In this application, each *Q*_*j*_(γ _*j*_) is a mixture of a left truncated normal distribution, a right truncated normal distribution, and a (non-truncated) normal distribution:

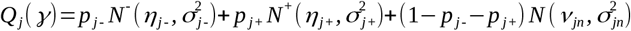

If 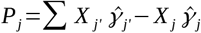 denotes a partial PGS where the effect sizes are based on the current estimate of *Q* (γ), then *Q*_*j*_(γ _*j*_) is updated by setting

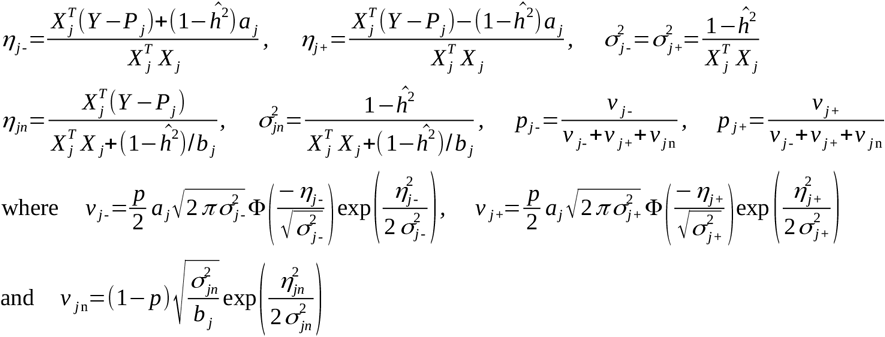

### Novel variational Bayes solver

The variational Bayes solver implemented in BOLT-LMM uses sequential genome-wide scans.^12^ This means that on each scan, it updates *Q*_*j*_(γ _*j*_) once for each SNP in the genome (i.e., it first updates *Q*_1_(γ_1_), then *Q*_2_(γ_2_), and so on until *Q*_*m*_(γ_*m*_)). BOLT-LMM performs multiple scans, continuing until LL, the approximate log likelihood, changes by less than a specified tolerance (by default, 0.0005). Typically, BOLT-LMM requires 50-150 scans (convergence is slowest when analyzing highly-heritable traits with very large sample sizes).

By contrast, our variational Bayes solver partitions the genome into chunks (by default, each chunk contains 256 SNPs), then uses chunk-based scans (see **Supplementary Figure 1** for an illustration). On Scan 1, our solver first updates

*Q*_*j*_(γ _*j*_) for SNPs in Chunk 1, continuing until LL has converged (the default tolerance is n/10^6^). Our solver then updates *Qj* (*γ j*) for SNPs in Chunk 2, then for SNPs in Chunk 3, continuing until it reaches the final chunk in the genome. On subsequent scans, our solver repeats the same process, except that it only considers chunks that had a sizeable impact on LL (specifically, it only revisits a chunk if on the previous scan, the updates for that chunk caused LL to change by more than n/10^6^). Our solver typically requires no more than ten scans to converge (because at this point, no chunks remain that have a sizeable impact on LL). We note that our solver has similarities to block coordinate descent methods, indicating that theoretical results regarding the convergence properties of the latter may also be applicable to our solver. ^38^

Our chunk-based solver has three main advantages over the genome-wide solver. Firstly, it prioritizes SNPs in regions that have a larger influence on LL (which is more efficient than simply treating all SNPs the same). Secondly, it enables on-the-fly reading of the genotypes (while this is, in theory, possible with the genome-wide solver, it would be necessary to read the data 50-150 times). Thirdly, it is more cache-friendly. **Supplementary Figure 2** compares our chunk-based solver with a genome-wide version when analyzing either 50k or 100k individuals. We see that both solvers have very similar accuracy (in that both produce models with very similar LL). However, we find that the chunk-based solver is substantially faster, reflecting that it performs fewer scans, and in turn fewer updates of *Q*_*j*_(γ _*j*_). For example, when analyzing 100k individuals, the genome-wide solver on average requires 56 scans (so updates each *Q*_*j*_(γ _*j*_) 56 times). By contrast, our chunk-based solver requires on average 4 scans, and on average updates each *Q*_*j*_(γ _*j*_) 11 times. **Supplementary Figure 37** shows that the default convergence criterion suffices, in the sense that results are almost unchanged if the tolerance is made five times smaller.

### Computing terms of the form V^**-1**^**A and KV**^**-1**^**A**

Our variational Bayes solver can be used to construct ridge regression PGS by assuming the prior distribution 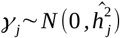. In this application, each *Q*_*j*_ (γ_*j*_) has the form 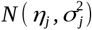, and is updated by setting

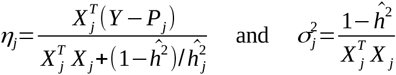

However, for this choice of prior, the posterior mean has an explicit form (derived in **Supplementary Note 4**). In particular, it can be shown that if P is a (genome-wide or LOCO) ridge regression PGS, then 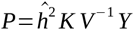 and 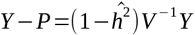 where V is a (genome-wide or LOCO) variance matrix. It follows that we can estimate terms of the form V^−1^Y and KV^−1^Y by using our variational Bayes solver to construct ridge regression PGS for Y, then dividing the estimated PGS by 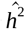 or dividing the corresponding residuals by 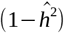. More generally, we can estimate terms of the form V^−1^A and KV^−1^A, where A is an arbitrary length-n vector, by performing the same steps but replacing Y with A (i.e., instead of constructing a PGS for Y, we construct a PGS for A). Therefore, our novel variational Bayes solver can not only be used to construct PGS, but also to compute the terms required when performing either Monte Carlo REML or using the Grammar-Gamma formula.^11^

### Overview of SPA

Here we summarize the SPA; for a fuller description, we recommend reading the supplementary material of the REGENIE paper.^13^ If A is a random variable, then its cumulant-generating function (CGF) is *K* _*A*_ (*t*)=log(*E*[exp(*tA*)]). The first, second and third derivatives of KA(t) are

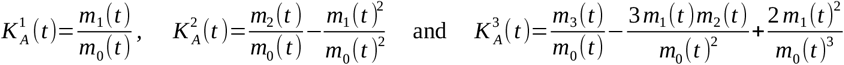

where *m*_k_ (*t*)=*E*[*A*^*k*^ exp(*tA*)]. The CGF is additive, such that if A and B are independent random variables, then *K*_*aA*+*bB*_ (*t*)=*K* _*A*_(*at*)+ *K*_*B*_ (*bt*). Similar relationships hold for the first, second and third derivatives:

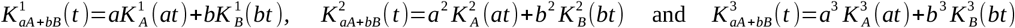

Now suppose A arises as a test statistic from regressing a phenotype on X_j_. The most common way to calculate a p-value is to compute *U*_j_ =(*A*−*E* [*A*])^2^/*Var* (*A*), where E[A] is the expectation of A under the null hypothesis, while Var(A) is its estimated variance, then to compare U_j_ to a *χ*^2^(1) distribution (or equivalently, compare its square root to a standard normal distribution). However, this approach assumes the null distribution of A is approximately normal, which can be inappropriate (e.g., if the phenotype is binary and very imbalanced, or when testing rare variants). The SPA method instead computes

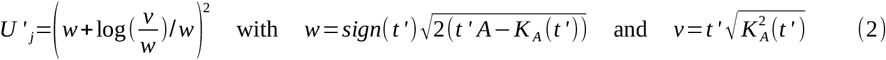

where t’ is the solution to K^1^(t’)=A, then obtains a p-value by comparing U’j to a χ^2^(1) distribution.

### Our Empirical SPA solver

Suppose the test statistic from regressing a phenotype B on X_j_ takes the form A=a1B1 + … + anBn. Our solver starts by calculating *K B* (*at*)=log(∑ exp(*atBi*)/*n*) for 41 predetermined values of a (evenly spaced between −2 and 2), and for 256 predetermined values of t (ranging from tmin to tmax, as described below). Note that these calculations correspond to assuming that the Bi are independent and identically distributed, and that their true distribution matches the observed distribution (see below).^23^ Our solver similarly computes 41 × 256 realizations for each of K^1^_B_ (at), K^2^_B_ (at) and K^3^_B_ (at). We refer to the predetermined values of a and t as “bin centres” and “knots”, respectively.

When LDAK-KVIK analyzes a quantitative phenotype, the (unscaled) Step 2 test statistic for SNP j is 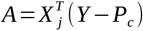, so we set ai=Xij and Bi=Yi-Pci in the above equations; when LDAK-KVIK analyzes a binary phenotype, the test statistic is 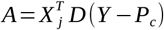, so we set ai=Xij and Bi=Dii(Yi-Pci). Note that when calculating the CDF of A and its derivatives, it is necessary to calculate terms of the form KB(ait), K^1^_B_(a_i_t) and K^2^_B_(a_i_t), and in general, ai will not match one of the 41 bin centres. Therefore, we approximate these terms using first, second and third order Taylor Series. Specifically, if a’ denotes the bin centre closest to ai, then we use the three approximations

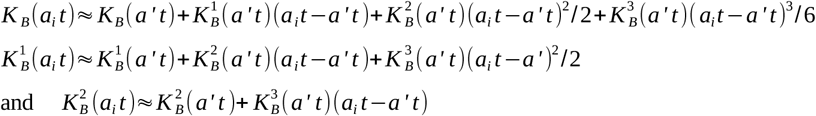

Our solver uses the second approximation to estimate K^1^_A_(t) = a_1_K^1^_B_(a_1_t) + … + a_n_K^1^_B_(a_n_t) for each of the 256 knots. It then identifies tL and tR, the knots immediately left and right of the solution to K^1^(t)=A, and uses the first and third approximations to calculate KA(tL), KA(tR), K^2^_A_(t_L_) and K^2^_A_(t_R_). Lastly, our solver uses linear interpolation to estimate t’, K_A_(t’) and K^2^_A_ (t’). Specifically, it computes 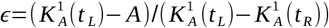 which estimates the location of the solution, relative to the closest two knots (e.g., ε=0.5 indicates that t’ is approximately halfway between t L and tR), then sets

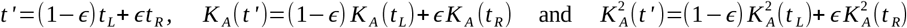

We now have all terms required to compute U’_j_, the χ^2^(1) test statistic defined in Equation 2. If our solver performed the above calculations naively, it would likely be no faster than existing SPA solvers, because computing each of the CGF and its derivatives still requires n operations (e.g., to compute KA(t) for a particular knot, it is necessary compute KB(ait) for each individual). However, the number of operations can be reduced dramatically by first summarizing X_j_ with respect to the bin centres. Specifically, if bk denotes the kth bin centre, and the function I(i,k) indicates whether ai is closest to bk, then our solver computes C1, …, C9, nine length-41 count vectors whose elements are

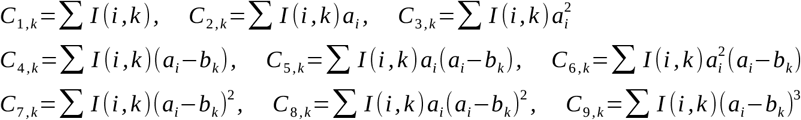

Given these nine vectors, we can rewrite the CDF of A and its derivatives as

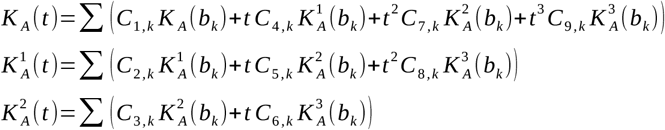

which can be computed using 164, 123 and 82 operations, respectively (numbers that, for large datasets, are substantially smaller than n).

When deciding the predetermined knot values, we initially use 256 quantiles from a Cauchy distribution,^23^ scaled such that 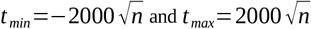. However, if our solver encounters a SNP for which this range is not sufficient (i.e., the corresponding test statistic is not bounded by K^1^_A_(t_min_) and K^1^_A_(t_max_)), then our solver resets the knot values so that t min and tmax are five times larger than their original values (this was not necessary for any of the analyses in this paper).

As explained above, our SPA solver assumes the true distribution of B, the adjusted phenotype, matches its empirical distribution. This has the advantage that our solver can be (validly) applied to any phenotype, whereas existing SPA solvers generally assume the phenotype has a Bernouilli distribution, so can only be applied to binary phenotypes. Furthermore, our solver makes no requirements on the form of X_j_ (although the predetermined bin centres assume that all elements of X_j_ are between −2 and 2, when this is not the case, our solver simply rescales X_j_). This means that our solver can automatically be applied to dosage data produced by genotype imputation, or used with non-SNP data.

**Supplementary Figure 38** shows that our empirical SPA solver is effective at controlling type 1 error when used either for rare binary phenotypes or for datasets with rare variants, while **Supplementary Figure 39** shows that the default values for the numbers of bin centres and knots suffice, in the sense that results are almost unchanged if we increase either of these values (e.g., increase the number of bin centres from 41 to 100, or the number of knots from 256 to 512).

### Comparison between our SPA solver and EmpSPA

Our SPA solver is inspired by EmpSPA,^39^ which was developed in order to compute p-values when performing Cox Regression (i.e., when analyzing survival data). In particular, we use the same formulae for KB(at), K^1^_B_(at) and K^2^_B_(at), and also copy their use of a Cauchy distribution when deciding the pre-determined knots. Saying this, a key difference is that our solver uses two-dimensional look-up tables (i.e., considers 41 values for a and 256 values for t), whereas EmpSPA uses one-dimensional look-up tables (in our notation, EmpSPA would be equivalent to setting a=1, then considering 10,000 values for t). Although, these two approaches have a similar one-off complexity (both involve evaluating the CGF and its derivatives for approximately 10,000 values), we find that using a two-dimensional look-up table is substantially more efficient. This is because, having mapped each value of X_ij_ to the nearest bin (i.e., obtained the count vectors C_1_, …, C_9_), our solver requires only 3×41 operations to evaluate K^1^_A_(t) for each knot, and having found the two closest knots, a further 6×41 operations to evaluate the corresponding values for KB(at) and K^2^_B_(at). Additionally, because we use uniformly-spaced bins, the (base-0) indexes of the nearest bins can be obtained by computing the integer part of 10(X_ij_+2). By contrast, to evaluate K^1^_A_(t) for a given value of t, EmpSPA must map each value of a_i_X_ij_ to the nearest pre-computed knot, then repeat this process for each t. As well as requiring more operations, the mapping is made more challenging by the fact that the pre-computed knots are not uniformly spaced (so requires a divide-and-conquer approach).

## REFERENCES

1. Uffelmann, E. et al. Genome-wide association studies. Nature Reviews Methods Primers 1, 59 (2021).

2. Visscher, P. M. et al. 10 Years of GWAS Discovery: Biology, Function, and Translation. Am J Hum Genet 101, 5–22 (2017).

3. Chen, Z. et al. China Kadoorie Biobank of 0.5 million people: survey methods, baseline characteristics and long-term follow-up. Int J Epidemiol 40, 1652–1666 (2011).

4. Keaton, J. M. et al. Genome-wide analysis in over 1 million individuals of European ancestry yields improved polygenic risk scores for blood pressure traits. Nature Genetics 56, 778–791 (2024).

5. Haines, J. L. et al. Complement factor H variant increases the risk of age-related macular degeneration. Science 308, 419–421 (2005).

6. Kurki, M. I. et al. FinnGen provides genetic insights from a well-phenotyped isolated population. Nature 613, 508–518 (2023).

7. Leitsalu, L. et al. Cohort Profile: Estonian Biobank of the Estonian Genome Center, University of Tartu. International Journal of Epidemiology 44, 1137–1147 (2014).

8. Lippert, C. et al. FaST linear mixed models for genome-wide association studies. Nature Methods 8, 833–835 (2011).

9. Zhou, X. & Stephens, M. Efficient multivariate linear mixed model algorithms for genome-wide association studies. Nature Methods 11, 407–409 (2014).

10. Yang, J., Zaitlen, N. A., Goddard, M. E., Visscher, P. M. & Price, A. L. Advantages and pitfalls in the application of mixed-model association methods. Nature Genetics 46, 100–106 (2014).

11. Svishcheva, G. R., Axenovich, T. I., Belonogova, N. M., van Duijn, C. M. & Aulchenko, Y. S. Rapid variance components–based method for whole-genome association analysis. Nature Genetics 44, 1166–1170 (2012).

12. Loh, P.-R. et al. Efficient Bayesian mixed-model analysis increases association power in large cohorts. Nature Genetics 47, 284–290 (2015).

13. Mbatchou, J. et al. Computationally efficient whole-genome regression for quantitative and binary traits. Nature Genetics 53, 1097–1103 (2021).

14. Zhou, W. et al. Efficiently controlling for case-control imbalance and sample relatedness in large-scale genetic association studies. Nat Genet 50, 1335–1341 (2018).

15. Loh, P. Mixed-model association for biobank-scale datasets. 50, 906–908 (2018).

16. Campos, A. I. et al. Boosting the power of genome-wide association studies within and across ancestries by using polygenic scores. Nature Genetics 55, 1769–1776 (2023).

17. Bycroft, C. et al. The UK Biobank resource with deep phenotyping and genomic data. (2018) doi:10.1038/s41586-018-0579-z.

18. Sudlow, C. et al. The UK Biobank resource with deep phenotyping and genomic data. PLoS Med. 12, e1001779 (2015).

19. Jiang, L. et al. A resource-efficient tool for mixed model association analysis of large-scale data. Nature Genetics 51, 1749–1755 (2019).

20. Berrandou, T., Balding, D. & Speed, D. LDAK-GBAT: Fast and powerful gene-based association testing using summary statistics. Am. J. Hum. Genet. 110, 23–29 (2023).

21. MacKay, D. J. Information Theory, Inference and Learning Algorithms. (Cambridge university press, 2003).

22. Dey, R., Schmidt, E. M., Abecasis, G. R. & Lee, S. A Fast and Accurate Algorithm to Test for Binary Phenotypes and Its Application to PheWAS. Am J Hum Genet 101, 37–49 (2017).

23. Ma, Y., Bi, W. & Zhang, J.-F. Empirical Saddlepoint Approximation and Its Application to Genome-Wide Association Studies. in 2021 40th Chinese Control Conference (CCC) 6380–6385 (2021). doi:10.23919/CCC52363.2021.9549628.

24. Bi, W., Fritsche, L. G., Mukherjee, B., Kim, S. & Lee, S. A Fast and Accurate Method for Genome-Wide Time-to-Event Data Analysis and Its Application to UK Biobank. The American Journal of Human Genetics 107, 222–233 (2020).

25. Speed, D., Holmes, J. & Balding, D. Evaluating and improving heritability models using summary statistics. Nat. Genet. 52, 458–462 (2020).

26. Zhang, Q., Privé, F., Vilhjálmsson, B. & Speed, D. Improved genetic prediction of complex traits from individual-level data or summary statistics. Nature Communications 12, 1–9 (2021).

27. Speed, D. et al. Reevaluation of SNP heritability in complex human traits. Nat. Genet. 49, 986– 992 (2017).

28. Speed, D. & Balding, D. SumHer better estimates the SNP heritability of complex traits from summary statistics. Nat. Genet. 51, 277–284 (2019).

29. Zou, H. & Hastie, T. Regularization and variable selection via the elastic net. J. R. Statist. Soc. B 67, 301–320 (2005).

30. Organization, W. H. The ICD-10 Classification of Mental and Behavioural Disorders: Clinical Descriptions and Diagnostic Guidelines. vol. 1 (World Health Organization, 1992).

31. de Leeuw, C. A., Mooij, J. M., Heskes, T. & Posthuma, D. MAGMA: generalized gene-set analysis of GWAS data. PLoS Comput Biol 11, e1004219 (2015).

32. Bakshi, A. et al. Fast set-based association analysis using summary data from GWAS identifies novel gene loci for human complex traits. Scientific Reports 6, 32894 (2016).

33. Momin, Md. M. et al. A method for an unbiased estimate of cross-ancestry genetic correlation using individual-level data. Nature Communications 14, 722 (2023).

34. Zeng, J. et al. Signatures of negative selection in the genetic architecture of human complex traits. Nat. Genet. 50, 746–753 (2018).

35. Schoech, A. et al. Quantification of frequency-dependent genetic architectures and action of negative selection in 25 UK Biobank traits. (2017).

36. Pruitt, K. D., Tatusova, T. & Maglott, D. R. NCBI reference sequences (RefSeq): a curated non-redundant sequence database of genomes, transcripts and proteins. Nucleic Acids Res 35, D61–65 (2007).

37. Pazokitoroudi, A. et al. Efficient variance components analysis across millions of genomes. Nat Commun 11, 4020 (2020).

38. Tseng, P. Convergence of a Block Coordinate Descent Method for Nondifferentiable Minimization. Journal of Optimization Theory and Applications 109, 475–494 (2001).

39. Ma, Y., Bi, W. & Zhang, J.-F. Empirical Saddlepoint Approximation and Its Application to Genome-Wide Association Studies. in 2021 40th Chinese Control Conference (CCC) 6380–6385 (2021).doi:10.23919/CCC52363.2021.9549628.

