## Supplementary Material for "LDAK-KVIK performs fast and powerful mixed-model association analysis of quantitative and binary phenotypes"

### Supplementary Notes

**1 Example LDAK-KVIK commands** **4**

**2 Technical details of LDAK-KVIK** **4**

**3 Using residual genotypes and phenotypes** **15**

**4 Expected residuals for the ridge regression PGS.** **16**

**5 Approximate logistic regression** **16**

**6 Data** **18**

**7 Existing MMAA Tools** **20**

**8 Attempts to estimate $\lambda$** **22**

#### Supplementary Figures

1 Illustration of our chunk-based variational Bayes solver. . . . . 26
2 Accuracy and speed of our variational Bayes solver. . . . . 27
3 Simulating phenotypes. . . . . 28
4 Type 1 error of LDAK-KVIK. . . . . 29
5 Type 1 error of LDAK-KVIK-GBAT. . . . . 30
6 Type 1 error of BOLT-LMM. . . . . 31
7 Type 1 error of REGENIE. . . . . 32
8 Type 1 error of fastGWA. . . . . 33
9 Type 1 error of GCTA-LOCO. . . . . 34
10 Type 1 error of SAIGE. . . . . 35
11 Type 1 error of all MMAA tools for the homogeneous dataset. . . . . 36
12 Fine-scale population structure. . . . . 37
13 Phenotypes where causal SNPs vary across ancestries. . . . . 38
14 Phenotypes where $\alpha$ varies across ancestries. . . . . 39
15 Runtime of LDAK-KVIK when analyzing 368 k individuals. . . . . 40
16 Computational requirements of LDAK-KVIK for very large datasets. . . . . 41
17 Power of MMAA tools for quantitative phenotypes. . . . . 42
18 Power of MMAA tools for common binary phenotypes. . . . . 43
19 Power of MMAA tools for rare binary phenotypes. . . . . 44
20 Power of gene-based association analysis for homogeneous data. . . . . 45
21 Estimates of $\alpha$ for the UK Biobank phenotypes. . . . . 46
22 Power of MMAA tools for the 40 quantitative UK Biobank phenotypes. . . . . 47
23 Power of MMAA tools for the 20 binary UK Biobank phenotypes. . . . . 48
24 Accuracy of Step 1 PGS when analyzing the UK Biobank phenotypes. . . . . 49
25 Gene-based association analysis of UK Biobank phenotypes. . . . . 50
26 Enlarged analysis of 62 quantitative phenotypes. . . . . 51
27 Approximate version of fastGWA. . . . . 52
28 LDAK-KVIK accurately estimates $\lambda$ . . . . . 54
29 BOLT-LMM can produce inflated test statistics. . . . . 55
30 Sensitivity of BOLT-LMM to choice of LD Scores. . . . . 56
31 Analyzing ascertained data. . . . . 57
32 Phenotypes with a strong environmental component. . . . . 58
33 Including non-European individuals in the analysis. . . . . 59
34 Reducing the number of Step 1 SNPs. . . . . 60
35 Robustness of our test for structure . . . . . 61
36 Adjusting estimates of relatedness for winners' curse. . . . . 62
37 Sensitivity of our variational Bayes solver to the convergence criterion. . . . . 63
38 Our SPA solver controls type 1 error. . . . . 64

39 Sensitivity of our SPA solver to parameter choices. . . . 66
40 LDAK-KVIK accurately estimates $\alpha$ and $h^2$. . . . 67
41 Padding missing phentoypes . . . . 68
42 Principal component axes of the UK Biobank data. . . . 69

#### Supplementary Tables

1 LDAK-KVIK runtimes when analyzing 368 k Individuals. . . . 70
2 The benefit of using multiple processors. . . . 71
3 The benefit of analyzing multiple phenotypes. . . . 72
4 Performance of our novel SPA solver. . . . 73
5 Ethnic compositions of the UK Biobank datasets. . . . 74
6 Baseline characteristics of the 40 quantitative UK Biobank phenotypes. . . . 75
7 Baseline characteristics of the 20 binary UK Biobank phenotypes. . . . 76

### Supplementary Note 1: Example LDAK-KVIK commands

Here we provide a simple example of how to run LDAK-KVIK; for more extensive documentation, see the LDAK-KVIK website (`www.ldak-kvik.com`). The following commands assume you have downloaded the Linux version of LDAK 6.1 (but note that we also provide a MAC version). Further, they assume that all data files are stored in PLINK format: specifically, that SNP genotypes are stored in the files `data.bed`, `data.bim` and `data.fam`, that measurements for a quantitative phenotype are stored in the file `data.pheno`, and that covariates are stored in the file `data.covar`. Finally, if performing a gene-based analysis (Step 3), you require gene annotations; this example uses the file `RefSeq_GRCh37.txt`, which can be downloaded from `www.dougspeed.com/resources`.

```
#Step 1 - Construct the LOCO PGS and estimate the test statistic scaling factor
./ldak6.1.linux --kvik-step1 kvik --bfile data --pheno data.pheno --covar data.covar

#Step 2 - Perform single-SNP association analysis
./ldak6.1.linux --kvik-step2 kvik --bfile data --pheno data.pheno --covar data.covar

#Step 3 - Perform gene-based association analysis
./ldak6.1.linux --kvik-step3 kvik --bfile data --genefile RefSeq_GRCh37.txt
```

Note that when analysing a binary phenotype, you should add `--binary YES` to Step 1.

The full results from single-SNP association analysis will be saved in the file `kvik.step2.assoc` (with a more concise version in `kvik.step2.summaries`). The results from the gene-based association analysis will be saved in the file `kvik.step3.remls.all`.

### Supplementary Note 2: Technical details of LDAK-KVIK

First we provide some definitions and summarize some existing methodology, then we explain the key innovations of LDAK-KVIK, and lastly we describe in detail the LDAK-KVIK algorithm. Note that most of this section assumes that the phenotype is quantitative, then at the end we explain the modifications used when the phenotype is binary.

#### 2.1 Definitions and existing methodology

We assume there are $n$ individuals, each genotyped for $m$ SNPs, recorded for $q$ covariates and measured for a quantitative phenotype. Let the $(n \times m)$ matrix $X'$ contain the genotypes, let the length-$n$ vector $Y'$ contain phenotypes, and let the $(n \times q)$ matrix $Z$ contain covariates. Note that we always include an intercept, so that $q \geq 1$ and $Z_{i,1} = 1$ $\forall i$. We use $C$ to denote the total number of chromosomes, use $X$ and $Y$ to denote, respectively, the genotypes and phenotypes after regressing out the covariates (via ordinary least-squares regression), and use $\lambda$ to denote the test statistic scaling factor. Without loss of generality, we standardize $X_j$, the $j$th column of $X$, to have mean zero and variance one. We similarly standardize $Y$.

Note that the regression models in this section describe how the residual phenotypes $Y$ depend on the residual genotypes $X$. In general, these regressions are equivalent to instead modelling how the original phenotypes $Y'$ depend on the original genotypes $X'$ and including covariates $Z$ (see Supplementary Note 3 for a justification).

**Linear random-effects regression model.** LDAK-KVIK repeatedly uses models of the form

$$Y = X_1\gamma_1 + ... + X_m\gamma_m + e = X\gamma + e, \tag{1}$$

where $\gamma$ and $e$ are, respectively, length-$m$ and length-$n$ vectors of random effects representing SNP effect sizes and environmental noise contributions. The models vary according to the assumed prior distributions for $\gamma$ and $e$. For example, ridge regression models use prior distributions of the form

$$\gamma_j \sim N(0, a_j) \quad \text{and} \quad e_i \sim N(0, \sigma_e^2),$$

where $N(0, a_j)$ and $N(0, \sigma_e^2)$ are mean-zero normal distributions with variance $a_j$ and $\sigma_e^2$, respectively, while elastic net models use prior distributions of the form

$$\gamma_j \sim pDE(b_j) + (1-p)N(0, c_j) \quad \text{and} \quad e_i \sim N(0, \sigma_e^2),$$

where $DE(b_j)$ denotes a double exponential distribution with rate parameter $b_j$ (note that calling this an elastic net model is an abuse of notation; to exactly mirror elastic net regularization, the prior distribution should be the product of a normal distribution and a double exponential distribution, rather than the sum).

We solve the linear random-effects regression models (i.e., estimate $\gamma$) using variational Bayes. We provide a brief description of variational Bayes in the Online Methods, while a detailed description is provided in the supplementary material of the BOLT-LMM publication [1].

**Heritability models.** The heritability model describes how $h_j^2$, the proportion of phenotypic variance explained by SNP $j$ is expected to vary across the genome [2]. If $f_j$ denotes the minor allele frequency of SNP $j$, then LDAK-KVIK uses heritability models of the form

$$\mathbb{E}[h_j^2] = w_j h^2/W, \quad \text{where} \quad w_j = [f_j(1-f_j)]^{1+\alpha} \quad \text{and} \quad W = \sum_{j=1}^{m} w_j. \tag{2}$$

The parameter $h^2$ is the phenotypic variance explained by all SNPs ($h^2 = \sum \mathbb{E}[h_j^2]$); when the data are homogeneous, $h^2$ equals the SNP heritability, but for heterogeneous data, $h^2$ is intermediate of the SNP heritability and family heritability[3–5]. The parameter $\alpha$ determines the relationship between the expected per-SNP heritabilities and MAF; if $\alpha > -1$, then $\mathbb{E}[h_j^2]$ increases with $f_j$, indicating that more common SNPs tend to contribute more heritability than less common SNPs, and vice versa [4]. Setting $\alpha = -1$ corresponds to assuming that $\mathbb{E}[h_j^2]$ is constant; we refer to this as the Uniform Heritability Model, and it is the model (implicitly) assumed by most software in statistical genetics (including BOLT-LMM [1], REGENIE [6], fastGWA [7], GCTA-LOCO [8] and SAIGE[9]). By contrast, our previous works, and those of others, have shown that values of $\alpha$ close to -0.25 are usually more appropriate when analyzing complex human traits [10–14].

Note that when using the linear random-effects regression model defined in Equation 1, the expected per-SNP heritability is $\mathbb{E}[h_j^2] = \mathbb{E}[\gamma_j^2] \times Var(X_j)/Var(Y) = \mathbb{E}[\gamma_j^2]$. Therefore, we can incorporate different heritability models by varying the parameters of the prior distributions for $\gamma$ and $e$. For example, if using a ridge regression model, we can incorporate the heritability model defined in Equation 2 by setting $a_j = w_j h^2/W$ and $\sigma_e^2 = 1 - h^2$ (because then $\mathbb{E}[h_j^2] = \mathbb{E}[\gamma_j^2] = w_j h^2/W$, as desired).

When using a linear random-effects regression model, the choice of heritability model determines the expected covariance between elements of $Y$. For example, if we assume the heritability model defined in Equation 2, and let $\Omega$ denote an $m \times m$ diagonal matrix such that $\Omega_{j,j} = w_j/W$, then the assumed phenotypic variance matrix is $V = Kh^2 + I(1 - h^2)$, where the genomic relatedness matrix $K = X\Omega X^T$.

**Partitioned randomized Haseman-Elston Regression.** Haseman-Elston Regression is a method-of-moment approach for estimating heritability, that searches for parameter values so that the expected phenotypic variance matrix most closely matches the observed phenotypic variance matrix [15]. First we describe the basic version of Haseman-Elston Regression, then we explain how it can be randomized and partitioned.

As explained above, the expected phenotypic variance matrix is determined by the heritability model, and takes the form $V = Kh^2 + I(1-h^2)$. Meanwhile, the observed variance matrix is $O = YY^T$ (note that $Y$ has mean zero, so does not need to be centred). We can estimate $h^2$ by regressing the upper-triangular values of values of $O$ onto the corresponding upper-triangular values of $K$ (note that it suffices to ignore the diagonal values because both $Tr(O)$ and $Tr(K)$, the traces of $O$ and $K$, respectively, are very close to $n$, while it suffices to ignore the lower-triangular values because both $O$ and $K$ are symmetric). The ordinary least-squares estimate of $h^2$ is

$$\hat{h}^2 = (K_U^T K_U)^{-1}(K_U^T O_U),$$

where $K_U$ and $O_U$ are vectors containing the upper-triangular values of $K$ and $O$, respectively.

Instead of computing $K_U^T K_U$ directly, randomized Haseman-Elston Regression writes this term as $K_U^T K_U = (Tr(KK) - \sum_i K_{i,i}^2)/2$, then uses the Monte Carlo approximation $Tr(KK) = \sum_r v_r^T KK v_r/R$, where $v_1, \ldots, v_R$ are length-$n$ random vectors with expected mean zero and expected variance one (in practice, the elements of each $v_r$ are sampled from a standard normal distribution). Note that with randomized Haseman-Elston Regression, it is not necessary to store all genotypes in memory. This is because if we divide the genome into $D$ chunks, we can write $Kv_r = X\Omega X^T v_r = X_1 \Omega_1 X_1^T v_r + \ldots + X_D \Omega_D X_D^T v_r$, where $X_d$ contains the genotypes for the $d$th chunk, and $\Omega_d$ is the corresponding sub-matrix of $\Omega$. Therefore, it suffices to read the data one chunk at a time, which substantially reduces memory requirements.

The partitioned version of Haseman-Elston Regression arises from generalizing the heritability model. For example, suppose we divide the genome into $B$ partitions, then assume

$$\mathbb{E}[h_j^2] = I(j,1) w_j H_1/W_1 + \ldots + I(j,b) w_j H_b/W_b + \ldots + I(j,B) w_j H_B/W_B, \quad \text{where} \quad W_b = \sum_j I(j,b) w_j. \tag{3}$$

Here, $I(j,b)$ is an indicator function that equals one (zero) if SNP $j$ is inside (outside) Partition $b$, while $H_b$ is the heritability of Partition $b$. The expected phenotypic variance matrix now takes the form $V = K_1 H_1 + \ldots + K_b H_b + \ldots + K_B H_B + I(1 - h^2)$, where $K_b$ is a genomic relatedness matrix computed using only the SNPs in

Partition $b$. Partitioned Haseman-Elston Regression estimates $H_1, \ldots, H_b, \ldots, H_B$ (and so also their sum, $h^2$), by regressing the upper-triangular values of $O$ jointly on the corresponding upper-triangular values of $K_1, \ldots, K_b, \ldots, K_B$.

Partitioned Haseman-Elston Regression has previously been advocated because it enables more realistic heritability models [15], but our motivation for using it is slightly different. Namely, we observe that the accuracy of basic Haseman-Elston Regression depends on the magnitude of $h^2$ (it tends to be more accurate when $h^2$ is small, and less accurate when $h^2$ is large). To understand why, note that the estimate of $h^2$ from basic Haseman-Elston Regression is equivalent to that you would obtain from one-step zero-start Newton-Raphson REML (i.e., from running Newton-Raphson REML if you required that the starting estimate of $h^2$ is zero, and stop after only one iteration). When $h^2$ is close to zero, one-step zero-start Newton-Raphson REML will likely be reasonably accurate (because the algorithm does not need to move far). However, when $h^2$ is moderate or large, one-step zero-start Newton-Raphson REML can perform poorly (because it needs to make a very large jump). It follows that the accuracy of partitioned Haseman-Elston Regression will depend on the magnitude of each $H_b$. However, provided we use sufficient partitions, $H_b$ will tend to be small, and therefore partitioned Haseman-Elston Regression will tend to be accurate.

**Monte Carlo REML.** Consider the mixed model $Y \sim N(0, Kh^2 + I(1-h^2))$. The REML estimate of $h^2$ maximizes

$$l(Y|h^2) = -\frac{1}{2}Y^T V^{-1} Y - \frac{1}{2}\log|V|, \quad \text{where} \quad V = Kh^2 + I(1-h^2).$$

We can find the maximum by treating the likelihood as a function of $h^2$ and $1-h^2$ (even though the two parameters are linearly dependent), then setting the (partial) derivatives with respect to $h^2$ and $1-h^2$ to zero:

$$\frac{dl}{dh^2} = 0 \Rightarrow Y^T V^{-1} K V^{-1} Y = Tr(KV^{-1}) \quad \text{while} \quad \frac{dl}{d(1-h^2)} = 0 \Rightarrow Y^T V^{-1} V^{-1} Y = Tr(V^{-1}).$$

It follows that $\hat{h}^2$, the REML estimate of $h^2$, satisfies

$$\delta(\hat{h}^2) = \frac{Y^T V^{-1} K V^{-1} Y}{Y^T V^{-1} V^{-1} Y} \Big/ \frac{Tr(KV^{-1})}{Tr(V^{-1})} = 1.$$

As noted by the authors of BOLT-LMM[1], the task of finding $\hat{h}^2$ is simplified by the fact that $\delta(h^2)$ tends to vary monotonically with $h^2$. Instead of computing the traces exactly, we can use the Monte Carlo estimates $Tr(KV^{-1}) = \sum_r v_r^T K V^{-1} v_r / R$ and $Tr(V^{-1}) = \sum_r v_r^T V^{-1} v_r / R$, where $v_1, \ldots, v_R$ are length-$n$ random vectors with expected mean zero and expected variance one (the same strategy utilized by randomized Haseman-Elston Regression).

**The two-step MMAA approach.** In the following paragraphs, we explain the rationale behind the two-step MMAA approach (e.g., that used by BOLT-LMM and REGENIE [1, 6]). Note that, for mathematical simplicity, we ignore the fact that most modern MMAA tools use LOCO (e.g., we treat the estimated variance matrix $\hat{V}$ and the PGS $P$ as fixed, when in practice, these will usually vary depending on the chromosome being tested).

Early MMAA tools (e.g., EMMA, GEMMA and FastLMM [16–18]) tested SNPs using models of the form

$$Y \sim N(X_j\beta_j, \hat{V}\sigma^2) \quad \text{with} \quad \hat{V} = K\hat{h}^2 + I(1-\hat{h}^2),$$

where $X_j$ is the SNP being tested and $\beta_j$ is its effect size. The generalized least-squares estimate of $\beta_j$ is

$$\hat{\beta}_j = \frac{X_j^T \hat{V}^{-1} Y}{X_j^T \hat{V}^{-1} X_j},$$

whose estimated variance is

$$Var(\hat{\beta}_j) = \frac{\hat{\sigma}^2}{(X_j^T \hat{V}^{-1} X_j)} \quad \text{where} \quad \hat{\sigma}^2 = \frac{Y^T \hat{V}^{-1} Y}{n-q},$$

and thus a $\chi^2(1)$ test statistic is

$$S_j = \frac{\hat{\beta}_j^2}{Var(\hat{\beta}_j)} = \frac{(X_j^T \hat{V}^{-1} Y)^2}{X_j^T \hat{V}^{-1} X_j \times \hat{\sigma}^2}.$$

Note that many existing two-step MMAA tools exclude $\sigma^2$, reflecting that if $\hat{V}$ accurately models the variance of $Y$, then $\sigma^2 = 1$ (and so its estimate can be ignored). However, we prefer to include this term, because we believe it guards against misspecification of $\hat{V}$ (e.g., imperfect estimation of $h^2$). Note also that the inclusion of $q$ in the denominator of $\hat{\sigma}^2$ reflects that we are using residual genotypes and phenotypes [1].

The early MMAA tools were computationally demanding due to the fact that when calculating the generalized least-squares estimates, they would explicitly compute $\hat{V}$, then its inverse. However, later software (e.g., GRAMMAR [19] and BOLT-LMM [1]), recognised that an equivalent (but much more efficient) alternative is to compute a PGS assuming the ridge regression model, then perform ordinary least-squares regression using the PGS as an offset. Although the resulting test statistics may be biased, this can be corrected for by scaling the test statistics by a constant.

Specifically, the two-step approach works as follows. Suppose we assume the ridge regression model $Y = X\gamma + e$, with $\gamma_j \sim N(0, \hat{h}_j^2)$ and $e_i \sim N(0, 1 - \hat{h}^2)$, then construct the PGS $P = X\hat{\gamma}$, where $\hat{\gamma}$ is the posterior mean of $\gamma$. It can be shown that $\hat{\gamma} = \hat{h}^2 K \hat{V}^{-1} Y$, and in turn that $Y - P = (1 - \hat{h}^2)\hat{V}^{-1} Y$ (see Supplementary Note 4 for a proof). An (unscaled) $\chi^2(1)$ test statistic from regressing $Y - P$ on $X_j$ is

$$T_j = \frac{(X_j^T \hat{V}^{-1} Y)^2}{X_j^T X_j \times \hat{s}^2} \quad \text{with} \quad \hat{s}^2 = \frac{Y^T \hat{V}^{-1} \hat{V}^{-1} Y}{n-q},$$

where $\hat{s}^2$ arises as an unbiased estimate of $Var(\hat{V}^{-1} Y)$. We can therefore write

$$S_j = \lambda'_j T_j, \quad \text{where} \quad \lambda'_j = \frac{X_j^T X_j \times \hat{s}^2}{X_j^T \hat{V}^{-1} X_j \times \hat{\sigma}^2}.$$

Empirical studies have found that $\lambda'_j$ tends to be almost constant across the genome [1], and can therefore be replaced by $\lambda'$, the mean value of $\lambda'_j$ computed across a small number of randomly-picked SNPs (we refer to this as the "Grammar-Gamma formula" [19]).

The authors of BOLT-LMM realised that the two-step approach described above can be improved by using a mixture prior for SNP effect sizes. They therefore proposed assuming the linear random-effects regression model

$$Y = X\gamma + e, \quad \text{with} \quad \gamma_j \sim pN\left(0, \frac{1-F}{p} \times \frac{\hat{h}^2}{m}\right) + (1-p)N\left(0, \frac{F}{1-p} \times \frac{\hat{h}^2}{m}\right) \quad \text{and} \quad e_i \sim N(0, 1 - \hat{h}^2),$$

where the hyperparameters $p$ and $F$ are set via cross-validation. Let $U_j$ denote the resulting test statistics (i.e., those obtained by computing $P = X\hat{\gamma}$, where $\hat{\gamma}$ is the revised posterior mean of $\gamma$, then regressing $Y - P$ on each $X_j$). Like $T_j$, these test statistics must be scaled by a constant, which we denote $\lambda$.

BOLT-LMM estimates $\lambda$ using a tool called LD Score Regression (LDSC)[20]. BOLT-LMM first runs LDSC using the scaled test statistics $\lambda' T_1, \ldots, \lambda' T_m$, where $T_1, \ldots, T_m$ are obtained by performing the two-step MMAA approach using a ridge regression PGS, and $\lambda'$ is estimated via the Grammar-Gamma formula[19]. BOLT-LMM then runs LDSC using the unscaled test statistics $U_1, \ldots, U_m$. If $A_1$ and $A_2$ denote the estimated intercepts from the two analyses, then BOLT-LMM set $\lambda = A_1/A_2$ (i.e., this value ensures that the two sets of scaled test statistics have the same estimated intercept).

**fastGWA.** When applied to quantitative phenotypes, fastGWA [7] performs generalized least-squares regression using the model $Y' \sim N(Z\theta + X_j'\beta_j, \hat{V}_S)$, where $\hat{V}_S$ is a sparse approximation of the estimated genome-wide variance matrix $\hat{V}$ (the authors of fastGWA recommend obtaining $\hat{V}_S$ by setting elements of $\hat{V}$ below 0.05 to zero)[7]. The sparsity of $\hat{V}_S$ enables fastGWA to rapidly compute $\hat{V}_S^{-1}Y$ and terms of the form $\hat{V}_S^{-1}X_j$, that are required when estimating SNP effect sizes and the corresponding variances. fastGWA reduces runtime further by using the Grammar-Gamma formula[19]. When applied to binary phenotypes, fastGWA switches from a linear mixed model to a logistic mixed model, then solves the latter by maximizing the quasi-likelihood[9, 21].

Note that LDAK-KVIK includes an approximate version of fastGWA (described in the Online Methods), that is used when analyzing binary phenotypes for datasets determined to have high structure.

**LDAK-GBAT.** We have previously developed LDAK-GBAT, a tool for gene-based association analysis using GWAS summary statistics and a reference panel [22]. LDAK-GBAT uses REML to solve the model

$$Y \sim N(0, K_S\sigma_S^2 + I(1 - \sigma_S^2)),$$

where $K_S$ is a "genomic" relatedness matrix constructed using only SNPs within the gene being tested, and $\sigma_S^2$ is the corresponding variance component, then performs a likelihood ratio test of $\sigma_S^2 > 0$ (using permutations to estimate the null distribution of the likelihood ratio test statistic). When applied to 109 phenotypes from the UK Biobank [23, 24], Million Veterans Program[25] and Psychiatric Genomics Consortium [26], LDAK-GBAT found at least 19% more significant genes than the existing tools MAGMA [27], fastBat [27], SKAT-O, PCA and ACAT (the last three tools are contained within the sumFREGAT software[28]).

Note that LDAK-GBAT requires a gene annotations file, which should be in Browser Extensible Data format[29]; i.e., it should contains one row for each gene, and four columns, that report the gene name and chromosome, and its start and end basepairs. For example, if the file contained a single line, with entries "ABC 7 0 10", this would indicate there is one gene, called ABC, which spans the first ten basepairs of Chromosome 7. We provide human gene annotations for both the GRCh37 and GRCh38 genome assemblies, constructed using RefSeq[30] annotations obtained from the UCSC Genome Browser[29], at `www.dougspeed.com/resources`.

#### 2.2 Key Innovations

LDAK-KVIK has six key innovations (note that these innovations are described in the Online Methods):

**1 - A novel variational Bayes Solver.** We have developed a chunk-based variational Bayes solver. This is not only used to construct PGS, but also to compute terms of the form $V^{-1}A$, where $V$ is a variance matrix and $A$ is a length-$n$ vector. Supplementary Figure 2 shows that when constructing PGS, our variational Bayes solver is 5-20 times faster than a standard (genome-wide) variational Bayes solver, and that when calculating $V^{-1}A$, our solver is 3-10 times faster than conjugate gradient descent. Furthermore, our variational Bayes solver has a trivial memory footprint (less than 1 Gb), because it never needs to read in more than 256 SNPs at a time.

**2 - A fast empirical SPA Solver.** We have developed an SPA solver, which is used by default in Step 2 when analyzing binary phenotypes. As shown in Supplementary Table 4, our solver has similar speed to fastSPA, the SPA solver implemented within REGENIE [6, 31]. However, we believe our solver has two advantages. Firstly, it is empirical, so can be applied to all types of phenotype (by contrast, fastSPA assumes a Bernoulli Distribution, so can only be used for binary phenotypes). Secondly, our solver does not require that the genotypes are sparse (whereas the efficiency of fastSPA relies on relatively few individuals having non-zero genotypes for each SNP). We expect that future work can exploit these two advantages, for example, by applying our SPA solver to non-binary phenotypes (e.g., count, ordinal and survival phenotypes) or by applying our SPA solver to non-sparse predictors (e.g., genotype probabilities or transcriptomic measurements).

**3 - Incorporates more realistic heritability models.** Most existing MMAA tools assume that expected per-SNP heritabilities are constant, which is equivalent to assuming the heritability model defined in Equation 2 with $\alpha = -1$. Supplementary Figure 24 shows that when analyzing the 40 quantitative UK Biobank phenotypes, the power advantage of LDAK-KVIK over BOLT-LMM [1] is mainly because the former constructs more accurate LOCO PGS, which in turn is because LDAK-KVIK estimates $\alpha$ from the data, whereas BOLT-LMM fixes its value to -1.

**4 - Uses a prior distribution for SNP effect sizes inspired by elastic net regularization.** While the use of the elastic net is fairly common in statistical genetics software [32, 33], we are not aware of it being used in MMAA tools. Supplementary Figure 24 shows that, when analyzing the 40 quantitative UK Biobank phenotypes, using the elastic net prior distribution provides a small advantage over using a mixture of two normal distributions (the assumption of Bolt-LMM [1]), and a large advantage over using a single normal distribution (similar to the assumption of REGENIE [6]).

**5 - A novel method for estimating $\lambda$.** We propose a general method for estimating $\lambda$ (whereas the Grammar-Gamma formula is only valid when using LOCO ridge regression PGS[19]). We believe that our method is superior to the general method proposed used by BOLT-LMM, because the latter requires strong assumptions both regarding how causal variation is distributed across the genome and how confounding inflates test statistics.

**6 - A novel test for structure.** We have developed a test for heterogeneity, based on the average squared correlation of SNPs on different chromosomes. For quantitative phenotypes, the result of this test influences how LDAK-KVIK calculates $\lambda$, while for binary phenotypes, the result dictates whether LDAK-KVIK switches to an approximate version of fastGWA.

#### 2.3 LDAK-KVIK Algorithm

LDAK-KVIK has two steps when used for single-SNP association analysis, and three steps when used for both single-SNP and gene-based association analysis. As a reminder, for now we assume the phenotype is quantitative, then later explain the changes for binary phenotypes.

**Step 1: Construct LOCO PGS and estimate $\lambda$**

Operation 1a - Test for structure

Operation 1b - Estimate $\alpha$ and $h_j^2$ using partitioned randomized Haseman-Elston Regression

Operation 1c - Revise the estimates of $h_j^2$ using Monte Carlo REML

Operation 1d - Determine suitable elastic net hyperparameters via cross-validation

Operation 1e - Construct LOCO PGS and estimate $\lambda$

**Step 2: Single-SNP association analysis**

Operation 2a - Calculate raw test statistics

Operation 2b - Scale test statistics

**Step 3: Gene-based association analysis**

Operation 3a - Run LDAK-GBAT using the results from Steps 1 & 2

First we describe each operation in turn, then we explain some implementation details. As a reminder, all regressions use residual genotypes and phenotypes, because this removes the need to include the covariate matrix $Z$ (see Supplementary Note 3 for a justification). The residual phenotypes (genotypes) are obtained by computing $Y = HY'$ ($X = HX'$), where $H = I - Z(Z^TZ)^{-1}Z^T$, then scaling $Y$ (columns of $X$) to have variance one.

**Operation 1a - Test for structure.** This test is fully described in Online Methods. However, in brief, LDAK-KVIK picks 512 SNPs randomly from across the genome, then computes $\hat{\rho}^2 = \bar{\rho}^2 - 1/(n-1)$, where $\bar{\rho}^2$ is the average squared correlation between pairs of SNPs on different chromosomes. LDAK-KVIK determines there is high structure if both $n\hat{\rho}^2 > 0.1$ and $\hat{\rho}^2$ is significantly greater than zero ($P < 0.001$).

**Operation 1b - Estimate $\alpha$ and $h_j^2$ using partitioned randomized Haseman-Elston Regression.** LDAK-KVIK first obtains $\hat{\alpha}$ and $\hat{h}^2$, estimates of $\alpha$ and $h^2$, respectively, by running randomized Haseman-Elston Regression using the partitioned heritability model defined in Equation 3. LDAK-KVIK considers five different values for $\alpha$ (-1, -0.75, -0.5, -0.25 & 0), then sets $\hat{\alpha}$ to the value that results in best-fitting $V$ (measured by the sum of squared differences between off-diagonal elements of $V$ and $O$), and sets $\hat{h}^2$ to the corresponding estimate of $h^2$. LDAK-KVIK then obtains $\hat{h}_j^2$, estimates of the per-SNP heritabilties, by setting $\hat{h}_j^2 = w_j\hat{h}^2/W$, where $w_j = [f_j(1-f_j)]^{1+\hat{\alpha}}$.

By default, LDAK-KVIK uses 40 equally-sized partitions and either ten random vectors (if $n < 40\,000$) or three random vectors (if $n \geq 40\,000$). Supplementary Figure 40 shows that this operation produces reasonable estimates of both $\alpha$ and $h^2$.

**Operation 1c - Revise the estimates of $h_j^2$ using Monte Carlo REML.** LDAK-KVIK first evaluates $\delta(0.5\times\hat{h}^2)$, $\delta(0.75\times\hat{h}^2)$ and $\delta(\hat{h}^2)$, where $\hat{h}^2$ is the estimate of $h^2$ from randomized Haseman-Elston Regression (e.g., if the estimate of $h^2$ from randomized Haseman-Elston Regression is 0.4, then LDAK-KVIK evaluates $\delta(0.2)$, $\delta(0.3)$ and

$\delta(0.4)$). LDAK-KVIK then uses linear interpolation or extrapolation to find an approximate solution to $\delta(h^2) = 1$. Finally, LDAK-KVIK recomputes the estimates of $h^2_j$ using the revised estimate of $h^2$ (continuing to use the estimate of $\alpha$ from Operation 1b). Supplementary Figure 40 shows that this operation reliably estimates $h^2$.

**Operation 1d - Determine suitable elastic net hyperparameters via cross-validation.** When constructing elastic net PGS, LDAK-KVIK assumes

$$\gamma_j \sim pDE\left(\left[\frac{2p}{(1-F)\hat{h}^2_j}\right]^{0.5}\right) + (1-p)N\left(0, \frac{F\hat{h}^2_j}{1-p}\right) \quad \text{and} \quad e \sim N(0, 1-\hat{h}^2). \tag{4}$$

In the prior distribution for SNP effect sizes, the parameter $p$ determines the contribution of the lasso component, while the parameter $F$ determines the expected contribution to variance from the ridge regression component. Note that this prior distribution is constructed so that $\mathbb{E}[h^2_j] = \hat{h}^2_j$ (i.e., the expected per-SNP heritabilities match their estimates from Operation 1c).

LDAK-KVIK uses cross-validation to obtain suitable values for $p$ and $F$. By default, it uses 90% of individuals to construct ten genome-wide PGS, for $(p, F)$ equal to (0.5,0.5), (0.5,0.3), (0.5,0.1), (0.1,0.5), (0.1,0.3), (0.1,0.1), (0.01,0.5), (0.01,0.3), (0.01,0.1) and (0,1). It then uses the remaining 10% of individuals to measure the accuracy of these PGS, selecting the values for $p$ and $F$ that result in the PGS with smallest mean-squared error. Note that if $MSE$ denotes the smallest mean-squared error, then LDAK-KVIK uses $n/MSE$ as an estimate of the effective sample size for the final association analysis (recall that $Var(Y) = 1$, so $1/MSE$ estimates the reduction in phenotypic variance when using the LOCO elastic net PGS as offsets).

**Operation 1e - Construct LOCO PGS and estimate $\lambda$.** Let $I(j, c)$ denote whether SNP $j$ is on Chromosome $c$, and let $S_c = (\sum_j I(j, c)\hat{h}^2_j)/\hat{h}^2$. LDAK-KVIK constructs $P_c$, the $c$th LOCO PGS, assuming the elastic net prior distribution for SNP effect sizes defined in Equation (4), with $p$ and $F$ set to the values identified in Operation 1d, and with $\hat{h}^2_j$ replaced by $\hat{h}^2_j I(j, c)/S_c$ (note that multiplying the estimated per-SNP heritabilities by $I(j, c)/S_c$ ensures that they will be zero for SNPs on Chromosome $c$, but continue to sum to $\hat{h}^2$).

If Operation 1a determines the dataset has low structure, then LDAK-KVIK sets $\lambda = 1$. Otherwise, LDAK-KVIK constructs a second set of LOCO PGS assuming the ridge regression prior distribution $\gamma_j \sim N(0, \hat{h}^2_j I(j, c)/S_c)$ and $e \sim N(0, 1-\hat{h}^2)$. It then uses least-squares linear regression with the model $\mathbb{E}[Y - P_c] = X_j\beta_j$ to compute two sets of unscaled $\chi^2(1)$ test statistics for 65,000 randomly-picked SNPs: for the first set (denoted by $U_j$), $P_c$ is the LOCO elastic net PGS corresponding to SNP $j$, while for the second set (denoted by $T_j$), $P_c$ is the corresponding LOCO ridge regression PGS. Next, LDAK-KVIK uses the Grammar-Gamma formula to compute $\lambda'$, the scaling factor corresponding to the second set of test statistics (by default it uses 20 randomly-picked SNPs)[19]. Finally, it finds $\lambda$ such that $\lambda' \sum_{j\in\mathbb{S}} T_j = \lambda \sum_{j\in\mathbb{S}} U_j$, where $\mathbb{S}$ contains the SNPs with both $\lambda U_j < 6$ and $\lambda' T_j < 6$ (because the definition of $\mathbb{S}$ depends on $\lambda$, the equality is solved iteratively).

**Operation 2a - Calculate raw test statistics**. If SNP $j$ is on Chromosome $c$, then LDAK-KVIK tests it for association using ordinary least-squares regression with the model $\mathbb{E}[Y - P_c] = A_j\beta_j$, where $P_c$ is the LOCO PGS constructed in Operation 1e, and either $A_j = X_j$ (the "exact model") or $A_j = X'_j$ (the "approximate model"). When using the exact model, the estimated effect size is

$$\hat{\beta}_j = \frac{X_j^T(Y - P_c)}{X_j^T X_j},$$

with estimated variance

$$Var(\hat{\beta}_j) = \frac{\hat{s}^2}{X_j^T X_j} \quad \text{where} \quad \hat{s}^2 = \frac{(Y - P_c)^T(Y - P_c)}{n - q},$$

and the corresponding $\chi^2(1)$ test statistic is

$$U_j = \frac{(X_j^T(Y - P_c))^2}{X_j^T X_j \times \hat{s}^2}.$$

For the approximate model, the formulae are the same, except that $X_j$ is replaced by $X'_j$. By default, LDAK-KVIK first tests all SNPs using the approximate model, then retests the SNPs with $P < 0.05$ using the exact model. This strategy mirrors the approach used by fastGWA[7], and can be substantially faster than using the exact model for all SNPs when there are many covariates (because it is only necessary to compute $X_j$ for a subset of SNPs).

**Operation 2b - Scale test statistics**. LDAK-KVIK reports three values for each SNP: an effect size estimate $\epsilon_1$, an estimate of the variance of the effect size estimate $\epsilon_2$, and a $\chi^2(1)$ test statistic $\epsilon_3$. LDAK-KVIK sets $\epsilon_1 = \lambda\hat{\beta}_j$ and $\epsilon_3 = \lambda U_j$, where $\hat{\beta}_j$ and $U_j$ are, respectively, the estimated effect size and test statistic calculated in Operation 2a, while $\lambda$ is the estimated scaling factor from Operation 1e. LDAK-KVIK then sets $\epsilon_2 = \epsilon_1^2/\epsilon_3$, which ensures that the three reported values are consistent (i.e., that $\epsilon_3 = \epsilon_1^2/\epsilon_2$).

**Operation 3a - Run LDAK-GBAT using the results from Steps 1 & 2**. LDAK-GBAT requires four inputs, a file containing gene annotations, an estimate of $\alpha$, GWAS summary statistics, and a reference panel. The gene annotations must be provided by the user (if analyzing human data, the user can download RefSeq[30] annotations from `www.dougspeed.com/resources`). LDAK-KVIK uses the estimate of $\alpha$ from Operation 1b and the summary statistics from Operation 2b. Finally, LDAK-KVIK randomly picks 5000 of the $n$ individuals, and uses their genotypes as an (in-sample) reference panel.

**Reducing runtime.** LDAK-KVIK minimizes memory usage by reading the SNP data on-the-fly. However, this means that a key determinant of its runtime is the number of times it must read each SNP once (we refer to this as the "number of scans"). For large datasets, the time to perform a scan is non-trivial (e.g., when analyzing 400 k individuals and 600 k SNPs, one scan takes about 0.25 CPU hours). Therefore, we have implemented LDAK-KVIK in a way that aims to minimize the total number of scans. For example, in Operation 1b, we run all the Haseman-Elston Regressions simultaneously, while we perform Operations 1c & 1d together. Furthermore, when performing Operations 1c & 1d, instead of waiting for convergence of the approximate log likelihood (the requirement in Operation 1e), we stop the Variational Bayes solver when the revised estimate of $h^2$ and the accuracy of the best-fitting PGS have both converged (changed by less than 0.005).

**Customizing LDAK-KVIK.** The above description of LDAK-KVIK assumed the default settings, which is what we recommend. However, if desired, all operations can be modified by adding options to the command line. For example, in Operation 1a, the user can change the number of SNPs used when testing for structure using the option "–num-pedigree-predictors", change the thresholds for determining a dataset has high structure using the options

"–MAI-threshold" and "–MAI-significance", or skip the test by adding "–check-pedigree NO" (the latter will result in LDAK-KVIK always estimating $\lambda$). In Operation 1b, the user can change the numbers of random vectors and partitions using the options "–num-MCMC" and "–num-divides", respectively (note that the former will also change the number of random vectors used in Operation 1c). In Operation 1c, the user can change the maximum number of scans performed by the variational Bayes solver using the option "–num-scans" (note that this will also change the maximum number of scans used in Operations 1d & 1e). In Operation 1d, the user can change the proportion of test individuals using the option "–cv-proportion". In Operation 1e, the variational Bayes solver considers a chunk converged when the approximate log likelihood changes by less than $n \times tol$; by default, $tol = 10^{-6}$, but the user can change this value using the option "–tolerance". Also in this operation, the user can change the number of SNPs used when estimating $\lambda'$ and $\lambda$ using the options "–num-calibration-predictors" and "–num-comparison-predictors", respectively (these options only have an impact if Operation 1a determines there is high structure).

**Multi-phenotype version of LDAK-KVIK.** LDAK-KVIK can accommodate multiple phenotypes in both Step 1 & 2. When analyzing more than one phenotype, LDAK-KVIK makes the following changes. If a phenotype has missing values, then the missing values are replaced by the observed mean for this phenotype (whereas when analyzing a single phenotype, the corresponding individuals are excluded from all analyses). Supplementary Figure 41 shows that this "padding" of missing phenotypes has minimal impact on test statistics, especially when the missing proportion is small (say, less than 10%). In general, analyzing multiple phenotypes reduces runtime, relative to analyzing each phenotype separately. This is mainly because many of the operations perform matrix-matrix multiplications, which are more efficient when combined, and because analyzing multiple phenotypes has limited impact on the number of scans. For example, Supplementary Table 3 shows that LDAK-KVIK takes the same time to analysis five phenotypes simultaneously as it does to analysis two of the phenotypes individually. Nonetheless, we usually recommend analyzing phenotypes individually, because we find that when using a high-performance cluster, it is normally more efficient to run many low-resource jobs, than fewer high-resource jobs.

#### 2.4 Binary Phenotypes

Suppose now the phenotype is binary, and let the length-$n$ vector $\mu$ contain the probabilities that each individual is a case (i.e., $\mu_i = P(Y'_i = 1)$). LDAK-KVIK obtains $\mu'$, an estimate of $\mu$, by regressing $Y'$ logistically on $Z$, then constructs $D$, an $(n \times n)$ diagonal matrix such that $D_{i,i} \propto \mu'_i(1 - \mu'_i)$ and $Tr(D^{-1}) = n$.

LDAK-KVIK then performs the operations described above, with three general changes (see Supplementary Note 5 for a justification). Firstly, the standardized residual genotypes are obtained by setting $X = H'X'$, where $H' = I - Z(Z^TDZ)^{-1}DZ^T$, then scaling columns of $X$ to have variance one. Secondly, $Y$ contains standardized adjusted phenotypes, obtained by setting $Y = D^{-1}(Y - \mu')$, then scaling $Y$ to have variance one. Thirdly, whenever using a linear random-effects model of the form $Y = X\gamma + e$, LDAK-KVIK assumes that the covariance matrix of the noise term is proportional to $D^{-1}$ (i.e., the assumption $e \sim N(0, I(1-h^2))$ is replaced by $e \sim N(0, D^{-1}(1-h^2))$). The third change means that the variance matrices have the revised form $V = Kh^2 + D^{-1}(1-h^2)$, that when performing Monte Carlo REML in Operation 1c, the derivative ratio becomes

$$\delta(h^2) = \frac{Y^TV^{-1}KV^{-1}Y}{Y^TV^{-1}D^{-1}V^{-1}Y} \Big/ \frac{Tr(KV^{-1})}{Tr(D^{-1}V^{-1})},$$

and that when computing test statistics in Operation 2a, LDAK-KVIK switches from ordinary least-squares regression to weighted least-squares regression (using $D$ as a weight matrix).

LDAK-KVIK makes two further changes when analyzing binary phenotypes. Firstly, for SNPs with association $p$-value below 0.05, it recomputes their test statistic using our novel SPA solver (i.e., replaces $\lambda U_j$ with $\lambda U_j'$, the SPA test statistic defined in the Online Methods). Secondly, when the test in Operation 1a determines there is high structure, LDAK-KVIK switches to an approximate version of fastGWA (described in the Online Methods). The second change is motivated by the observations that when analyzing binary phenotypes, all MMAA tools tend to have very similar power, and that the choice of prior distribution for SNP effect sizes is less important when there is high structure. Therefore, we find that switching to fastGWA has limited impact on power, but has the advantage of being about three times faster.

#### Supplementary Note 3: Using residual genotypes and phenotypes

Here we justify why our regressions use residual genotypes and phenotypes (or in the case of binary phenotypes, residual genotypes and adjusted phenotypes).

Suppose we regress the quantitative phenotype $Y'$ on $Z$ and $X'$ using ordinary least-squares regression. The estimated effect sizes are

$$\hat{\theta} = \begin{pmatrix} Z^TZ & Z^TX' \\ X'^TZ & X'^TX' \end{pmatrix}^{-1} \begin{pmatrix} Z^TY' \\ X'^TY' \end{pmatrix}.$$

We can evaluate the first term using a blockwise inversion. If we let $H = I - Z(Z^TZ)^{-1}Z^T$, then

$$\hat{\theta} = \begin{pmatrix} (Z^TZ)^{-1} + (Z^TZ)^{-1}Z^TX'(X'^THX')^{-1}X'^TZ(Z^TZ)^{-1} & (Z^TZ)^{-1}Z^TX'(X'^THX')^{-1} \\ -(X'^THX')^{-1}X'^TZ(Z^TZ)^{-1} & -(X'^THX')^{-1} \end{pmatrix} \begin{pmatrix} Z^TY' \\ X'^TY' \end{pmatrix}.$$

If $\theta_X$ denotes the sub-vector of $\theta$ corresponding to the genotypes, then its estimate is

$$\hat{\theta}_X = -(X'^THX')^{-1}X'^TZ(Z^TZ)^{-1}Z^TY' + (X'^THX')^{-1}X'^TY' = (X'^THX')^{-1}X'^THY'.$$

The matrix $H$ is the complement of the hat matrix when performing ordinary least-squares regression with predictor matrix $Z$ (i.e., $HA$ is the residual of the matrix $A$ after regressing on $Z$). Further, $H$ is idempotent (i.e., $HH = H$). It follows that

$$\hat{\theta}_X = ((HX)^T(HX))^{-1}(HX)^T(HY') = (X^TX)^{-1}X^TY.$$

Thus we can see that the estimated SNP effect sizes from regressing $Y'$ on $Z$ and $X'$ are identical to those from regressing $Y$ on $X$. The only complication with this approach is that when estimating the variance of the effect sizes, we must allow for the fact we have regressed out covariates (i.e., remember to divide by $n - q$ instead of $n - 1$).

Note that the above mathematics can be used to show that when performing weighted least-squares regression (with weight matrix $D$), the estimated effect sizes from regressing $Y'$ on $Z$ and $X'$ are identical to those from regressing $H'Y'$ on $H'X'$, where $H' = I - Z(Z^TDZ)^{-1}Z^TD$. However, when we perform weighted least-squares regression, we take a slightly different approach. Specifically, while we do use residual genotypes (i.e., set $X = H'X'$), we use adjusted phenotypes instead of residual phenotypes (i.e., set $Y = D^{-1}(Y' - \mu)$ instead of $Y = H'Y'$). This reflects that we use weighted least-squares regression as an approximation to logistic regression (as explained in Supplementary Note 5). Furthermore, it can be shown that when using residual genotypes and adjusted phenotypes, the resulting score test statistic matches that obtained from regressing $Y'$ logistically on $X'$ and $Z$ (see Appendix A of Dey *et al.*[31] for a proof). However, we recognise that, in practice, the impact of replacing residual phenotypes with adjusted phenotypes is likely to be small (essentially, we are replacing residual phenotypes from weighted linear regression with weighted residual phenotypes from logistic regression).

#### Supplementary Note 4: Expected residuals for the ridge regression PGS.

Let $V = Kh^2 + I(1 - h^2)$, with $K = X\Omega X^T$. Here we prove the statement in the Online Methods that when constructing a ridge regression PGS $Y = X\hat{\gamma}$, assuming

$$Y = X\gamma + e, \quad \text{with} \quad \gamma_j \sim N(0, \Omega_{j,j}h^2) \quad \text{and} \quad e_i \sim N(0, 1 - h^2),$$

then the PGS takes the form $P = h^2KV^{-1}Y$ and the corresponding residuals are $Y - P = (1 - h^2)V^{-1}Y$.

With the priors defined above, the posterior distribution of SNP effect sizes satisfies

$$P(\gamma|Y) \propto \exp\left(-\frac{(Y - X\gamma)^T(Y - X\gamma)}{2(1 - h^2)}\right) \times \exp\left(-\frac{\gamma^T\Omega^{-1}\gamma}{2h^2}\right),$$

which can be rewritten as

$$P(\gamma|Y) \propto \exp\left(-\frac{(\gamma - A)^TB^{-1}(\gamma - A)}{2}\right) \quad \text{where} \quad B^{-1} = \frac{X^TX}{1 - h^2} + \frac{\Omega^{-1}}{h^2} \quad \text{and} \quad A = (X^TX + \frac{1 - h^2}{h^2}\Omega^{-1})^{-1}X^TY.$$

Therefore, the posterior distribution of $\gamma$ is multivariate normal, with mean $A$ and variance $B$. By applying the Woodbury matrix identify, we can write

$$A = \left(\frac{h^2}{1 - h^2}\Omega - \frac{h^2}{1 - h^2}\Omega X^T\left(\frac{h^2}{1 - h^2}X\Omega X^T + I\right)^{-1} X\frac{h^2}{1 - h^2}\Omega\right) X^TY$$

Using the definitions of $V$ and $K$ from above, it follows that the PGS constructed using the posterior mean of $\gamma$ is

$$P = X\hat{\gamma} = XA = \frac{h^2}{1 - h^2}K\left[I - \left(\frac{V}{1 - h^2}\right)^{-1}\frac{h^2}{1 - h^2}K\right]Y = h^2KV^{-1}Y,$$

and the corresponding vector of residuals is

$$Y - P = \left(I - h^2KV^{-1}\right)Y = (1 - h^2)V^{-1}Y.$$

Note that when the phenotype is binary, we instead obtain

$$P = h^2KV^{-1}Y \quad \text{and} \quad Y - P = (1 - h^2)D^{-1}V^{-1}Y,$$

where the variance matrix now takes the form $V = Kh^2 + D^{-1}(1 - h^2)$.

#### Supplementary Note 5: Approximate logistic regression

In this section, we assume $Y'$ is a binary phenotype, and explain our reason for switching from ordinary least-squares regression to weighted least-squares regression. As a reminder, we use the length-$n$ vector $\mu$ to denote the probabilities that each individual is a case, while $\mu'$ is an estimate of $\mu$ from regressing $Y'$ logistically on $Z$ (i.e., $\mu'_i$ is the estimated probability that Individual $i$ is a case given their covariates).

Suppose we were to regress $Y'$ logistically on the matrices $Z$ and $X'$ using the model $\log(\mu/(1-\mu)) = A\theta$, where the matrix $A$ contains both the covariates and the genotypes (i.e., $A = [Z\ X']$). The corresponding log likelihood would be

$$l(Y'|A) = \sum_i \big(Y_i \log(\mu_i) + (1-Y_i)\log(1-\mu_i)\big),$$

whose first and second derivatives can be expressed as

$$\frac{dl}{d\theta} = A^T(Y' - \mu) \quad \text{and} \quad \frac{d^2 l}{d\theta^2} = -A^T D' A,$$

where $D'$ is a diagonal matrix with $D'_{i,i} = \mu_i(1-\mu_i)$. We could estimate $\theta$ using Newton-Raphson iterations of the form

$$\theta^{k+1} = \theta^k + (A^T D' A)^{-1} A^T (Y' - \mu),$$

where $\theta^k$ denotes the estimated coefficients after the $k$th iteration. If we rewrite this as

$$\theta^{k+1} = \theta^k + (A^T D' A)^{-1} A^T D' [D'^{-1}(Y' - \mu)],$$

then we can see that the updated estimate of $\theta$ is similar to that obtained by regressing the adjusted phenotype $D'^{-1}(Y' - \mu)$ linearly on $A$ with weight matrix $D'$ (in fact, the two estimates are equal when $\theta^k = 0$).

Now let $\theta_Z$ and $\theta_X$ denote the sub-vectors of $\theta$ corresponding to the covariates and genotypes, respectively. Further, suppose $\theta_Z^0$, the starting values for $\theta_Z$, are set to their values from regressing $Y'$ logistically on $Z$, while $\theta_X^0$, the starting values for $\theta_X$, are set to zero. It can be shown that the estimate of $\theta_X$ after one Newton-Raphson iteration is

$$\theta_X^1 = (X_j^T D X_j)^{-1} X_j^T D Y,$$

where $D$ is the realization of the weight matrix when $\mu = \mu'$, $X$ contains the residuals from regressing columns of $X'$ linearly on $Z$ with weight matrix $D$, and $Y = D^{-1}(Y' - \mu)$.

Therefore, our decision to switch to weighted least-squares regression when the phenotype is binary, is because the resulting estimates of SNP effect sizes can be considered an approximation of those obtained from logistic regression. We note that we could improve the accuracy of this approximation by updating the weight matrix $D$ during the algorithm (instead of fixing its value at the start), in which case our approach would mirror the quasi-likelihood approach used by SAIGE[9] and fastGWA[7]. However, updating $D$ would introduce computational challenges (e.g., there would need to be a separate $D$ for each PGS, which would prevent us from computing terms such as $X^T D(Y-P)$ for multiple PGS at once). Furthermore, we believe the benefit of updating $D$ would be small. This is because the major advantage of switching from linear to logistic regression is when there are relatively few cases, but in these circumstances, the proportion of variance explained by the SNPs tends to be small, and therefore the updated $D$ would be relatively close to its starting value. This is evidenced by the results in Supplementary Figure 27, which show that our approximate version of fastGWA (which does not update $D$) has well-controlled type 1 error.

### Supplementary Note 6: Data

We use genotypic and phenotypic data from the UK Biobank [23, 24] (obtained via application 21432).

#### 6.1 Genotypes

After excluding individuals missing phenotypic information, or those who have withdrawn consent, the UK Biobank contains approximately 487 k individuals, directly genotyped for approximately 784 k SNPs. We constructed six datasets: the "white dataset" contains 367 981 individuals and 690 264 SNPs, the "homogeneous dataset", "family dataset", "twins dataset" and "British-Irish dataset" each contain 63 000 individuals and 690 264 SNPs, while the "multi-ancestry dataset" contains 63 000 individuals and 639 727 SNPs. We primarily used the white dataset when analysing the real UK Biobank phenotypes, and used the five remaining datasets for simulations.

To create the white dataset, we first restricted to the 408 868 individuals listed in data field 22006 (those who self-identify as being white British, and whom UK Biobank determined "have very similar genetic ancestry based on a principal components analysis of the genotypes"), then randomly sampled 367 981 (90%) of these individuals (we use the remaining 40 887 individuals for testing the accuracy of Step 1 PGS). There were 690 264 autosomal, biallelic SNPs with MAF $> 0.001$ and genotype call rate $> 90\%$.

We constructed the homogeneous dataset by reducing the white dataset to 63 000 distantly-related individuals (we removed samples until there were no first- or second-degree relatives, based on relationships inferred by the software KING[34]). We constructed the family dataset by reducing the white dataset to 15 750 non-overlapping pairs of first-degree or second-degree relatives and 31 500 distantly-related individuals (again, using relationships inferred by KING). We constructed the twins dataset by randomly picking 31 500 individuals from the homogeneous dataset and duplicating their genomes.

We created the British-Irish dataset by combining 11 737 unrelated individuals who identified as being white Irish, with 51 263 unrelated individuals who identified as being white British. Lastly, we created the multi-ancestry dataset by identifying the 56 319 individuals who did not identify as being white British, and adding to these 6 681 individuals from the white dataset. For this dataset, there were 639 727 autosomal, biallelic SNPs with MAF $> 0.001$ and genotype call rate $> 90\%$.

Supplementary Figure 42 provides principal component plots for the UK Biobank data, while Supplementary Table 5 gives an overview of the ethnic composition of each dataset. Note that when analyzing simulated phenotypes, we included the top ten principal component axes as covariates, while when analyzing real phenotypes, we included the top ten axes, age, sex, the square of age, and age times sex.

#### 6.2 Simulated phenotypes

Note that in this section, there is a slight abuse of notation, because we use the index $j$ to denote the $j$th causal SNP (instead of the $j$th SNP overall).

We first partitioned the genome into three: the "causal partition" contains the starts of each chromosomes, the "buffer partition" contains the middle 4 Mb of each chromosome, while the "null partition" contains the ends of each chromosome (see Supplemental Figure 3). When simulating quantitative phenotypes, we specified $m'$, the number of causal SNPs (picked at random from the causal partition), $h^2$, the heritability contributed by all causal SNPs, and the power parameter $\alpha$. When simulating binary phenotypes, we also specified $K$, the prevalence. Note that the choices of $m'$, $h^2$ and $\alpha$ determine the values of $\mathbb{E}[h_j^2]$, the expected heritability contributed by the $j$th causal SNP.

We generated quantitative phenotype for the homogeneous dataset using the model

$$Y = \sum_{j=1}^{m'} X_j \gamma_j + A\theta_1 + e,$$

where $X_j$ and $\gamma_j$ denote the genotypes and effect size of the $j$th causal SNP, respectively, $A$ is a length-$n$ vector containing the first principal component axes, $\theta_1$ is a scalar, while $e$ is a length-$n$ vector representing environmental noise. We sampled $\gamma_j$ and $e_i$ independently from the mean-zero normal distributions $N(0, \hat{h}_j^2)$ and $N(0, 1 - h^2)$, respectively, which ensures that the expected heritability of the $j$th causal SNP equals $\mathbb{E}[h_j^2]$, then we set $\theta_1$ so that the $Var(Y - A\theta_1) = 0.95 \times Var(Y)$.

To generate binary phenotype for the homogeneous dataset, we first simulated liabilities using the model

$$L = \sum_{j=1}^{m'} X_j \gamma_j + A\theta_1 + e$$

with $\gamma_j$, $\theta_1$ and $e_i$ generated the same as for quantitative phenotypes. We then obtained phenotypes by dichotomizing the liabilities. Specifically, having decided the prevalence $K$, we set $Y_i = 1$ if $L_i > \Phi(K)^{-1}$, and $Y_i = 0$ if $L_i \leq \Phi(K)^{-1}$, where $\Phi^{-1}$ is the inverse CDF of a standard normal distribution.

When analyzing the family and twins datasets, we typically simulated quantitative and binary phenotypes using the same models we used for the homogeneous dataset. However, for Supplementary Figure 32, we simulated additional quantitative phenotypes where common environment explains 10% of the total phenotypic variance. We generated these phenotypes using the model

$$Y = \sum_{j=1}^{m'} X_j \gamma_j + B\theta + e,$$

where $B$ is an $(n \times R)$ matrix that indicates which individuals belong to each of the $R$ related pairs (for the family dataset, $R = 15\,750$, while for the twins dataset, $R = 31\,500$), and the elements of the length-$R$ vector $\theta$ are sampled from a standard normal distribution, then scaled so that $Var(Y - B\theta) = 0.9 \times Var(Y)$. This model is the same as that used by Jiang *et al.*, except that their dataset included individuals who were related to more than one other person, and so they first partitioned individuals into distinct families, then used columns $B$ to indicate which individuals belonged to each family.

We generated quantitative phenotypes for the British-Irish dataset using the model

$$Y = \sum_{j=1}^{m'} X_j \gamma_j + A\theta_1 + e,$$

where the vector $A$ now indicates which individuals are Irish, and we set $\theta_1$ so that the $Var(Y - A\theta_1) = 0.8 \times Var(Y)$.

We generated quantitative phenotypes for the multi-ancestry dataset using the model

$$Y = \sum_{j=1}^{m'} X_j \gamma_j + e,$$

allowing either $X_j$ or $\gamma_j$ to vary based on ancestry (specifically, we allowed either the choice of causal SNPs or the assumed value for $\alpha$ to depend on whether an individuals was classified as European, Asian or African).

##### 6.3 Real phenotypes

For most analyses of real phenotypes, we used either the 40 quantitative or 20 binary UK Biobank phenotypes detailed in Supplementary Tables 6 & 7, respectively. Note that across the 368 k individuals in the white dataset, the quantitative phenotypes have on average 5.1% missing values (range 0.2% to 13.6%), whereas the binary phenotypes are complete (because everyone not identified as being a case, was automatically assumed to be a control). However, for one analysis, we instead used 62 quantitative UK Biobank phenotypes, obtained by combining our 40 quantitative phenotypes with the 22 extra phenotypes listed in Supplementary Table 4 of Jiang *et al.*[7] (see `www.ldak-kvik.com/summaries` for more details).

#### Supplementary Note 7: Existing MMAA Tools

We benchmark the performance of LDAK-KVIK against classical regression and five existing MMAA tools: BOLT-LMM [1], REGENIE [6], fastGWA [7], GCTA-LOCO [8] and SAIGE[9]. Note that REGENIE and fastGWA are designed for both quantitative and binary phenotypes, whereas BOLT-LMM and GCTA-LOCO are designed for quantitative phenotypes, while SAIGE is designed for binary phenotypes. Here we provide a summary of each tool. Note that all five existing MMAA tools (implicitly) assume that $\mathbb{E}[h_j^2]$ is constant (equivalent to setting $\alpha = -1$ in Equation 2).

**BOLT-LMM.** BOLT-LMM [1] is a two-step MMAA tool. In Step 1, it creates LOCO Mixture-Normal PGS, where the SNP effect sizes are assigned the prior distribution

$$\gamma_j \sim pN\left(0, \frac{1-F}{p} \times \frac{\hat{h}^2}{m}\right) + (1-p)N\left(0, \frac{F}{1-p} \times \frac{\hat{h}^2}{m}\right).$$

BOLT-LMM estimates $h^2$ using Monte Carlo REML, and determines suitable values for $p$ and $F$ via cross-validation. BOLT-LMM estimates the test statistic scaling factor $\lambda$ using LDSC[20] (as explained in Supplementary Note 2).

We find that BOLT-LMM is generally the most powerful existing MMAA tool for quantitative phenotypes, reflecting the benefits of using a mixture prior distribution for SNP effect sizes when constructing PGS. However, we also find that BOLT-LMM is computationally demanding, due to its genome-wide variational Bayes solver, and its use of conjugate gradient descent (both when performing Monte Carlo REML and when calculating the test statistic scaling factor).

Note that BOLT-LMM provides the option to replace the mixture prior distribution described just above, with the ridge regression prior $\gamma_j \sim N(0, \hat{h}_j^2)$. The resulting tool is referred to as BOLT-LMM-inf, and is mathematically equivalent to, but substantially faster than, GCTA-LOCO (described below).

When running BOLT-LMM, we used the option "–lmmForceNonInf" to ensure it always computes LOCO PGS using the mixture prior distribution for $\gamma_j$ (otherwise, the software switches to the infinitissimal prior if the improvement from using the mixture prior is expected to be less than 1%). When analyzing the simulated phenotypes, we used the option "—LDscoresUseChip" so that BOLT-LMM computes LD scores from the data. This is appropriate because for our simulations, the causal variants were picked from the genotyped predictors. By contrast, when analyzing the UK Biobank phenotypes, we followed the recommendation of the authors and used the option "–LDscoresFile" to provide the European LD scores supplied with the software (these were computed using 1000 Genome Project[35] data for 379 individuals and approximately 10M common SNPs[36]). Further, we used the option "–predBetasFile" to save the estimated effect sizes of the (genome-wide) Bolt-LMM PGS (this allowed us to measure the accuracy of the BOLT-LMM PGS when applied to independent data, and compare this to the accuracy of the LDAK-KVIK PGS).

**REGENIE.** REGENIE [6] is a two-step MMAA tool. In Step 1, it creates PGS using stacked ridge regression. First, it divides the genome into blocks, then for each block it creates five PGS assuming the effect size prior distribution $\gamma_j \sim N(h^2/m)$, with $h^2$ set to 0.01, 0.25, 0.5, 0.75 or 0.99. Finally, it uses ridge regression with either a linear model (quantitative traits) or logistic model (binary traits) to combine the block-based PGS into LOCO PGS. REGENIE does not compute a test statistic scaling factor (so in effect, always assumes $\lambda = 1$).

REGENIE is computationally efficient, reflecting that it only needs to store one block of SNPs at a time, and that the block-based PGS are fast to compute and combine. It also parallelizes across phenotypes very efficiently, because the most time-consuming operations, reading and standardizing SNPs, and inverting a block-based correlation matrix, only need to be performed once regardless of the number of phenotypes. However, we find that REGENIE is less powerful than BOLT-LMM, due to the infinitesimal nature of its prior distribution for SNP effect sizes.

When running REGENIE, we set the block-size to 1000 SNPs. When analyzing binary phenotypes, REGENIE recomputes small $p$-values using either its implementation of fastSPA [31], or using an approximate Firth correction [37]; we followed the recommendation of the authors and selected the latter.

**fastGWA.** A description of fastGWA is provided in Supplementary Note 2. While our applications of fastGWA demonstrated its computational efficiency, they also found that it tended to be the least-powerful MMAA tool. The latter reflects that a sparse variance matrix will poorly capture the contributions of SNPs distant from the SNP being tested (consider the extreme case, where all off-diagonal terms in $K$ are set to zero, in which case the model used by fastGWA would be almost identical to that used by classical linear regression). Furthermore, restricting to a sparse variance matrix means that fastGWA can only account for heterogeneity due to familial relatedness, but not due to population structure.

**GCTA-LOCO.** GCTA-LOCO [8] performs generalized least-squares regression using the model $Y \sim N(Z\theta +$

$X_j\beta_j, \hat{V}_c)$, where $\hat{V}_c$ is an estimated LOCO variance matrix taking the form $\hat{V}_c = Kh^2 + I(1-h^2)$. GCTA-LOCO tends to be more powerful than fastGWA because it does not require the variance matrices to be sparse (therefore, it better captures the contributions of SNPs distant from the SNP being tested). However, GCTA-LOCO tends to be less powerful than BOLT-LMM, reflecting that its model arises from assuming an infinitesimal prior distribution for SNP effect sizes. Further, GCTA-LOCO is the most computationally demanding method, because it requires construction of the $n \times n$ variance matrix (whereas fastGWA uses a compact approximation of the variance matrix, while the other tools use algorithmns that do not explicitly construct the variance matrix).

**SAIGE.** SAIGE [9] uses logistic mixed-model regression. Specifically, it assumes $\log(\frac{\mu_i}{1-\mu_i}) = Z\theta + X_j\beta_j + X\gamma$, where $\mu_i = P(Y_i' = 1)$ is probability that the $i$th individual is a case, and the SNP effect sizes are assumed to have the prior distribution $\gamma_j \sim N(0, \hat{h}^2/m)$. SAIGE uses REML to obtain the estimate of $h^2$, uses variational Bayes to construct the PGS $X\gamma$, and uses conjugate gradient descent to compute the Grammar-Gamma estimate of the test statistic scaling factor $\lambda$ (these choices mirror those used by BOLT-LMM, except that for computational reasons, SAIGE replaces the true model likelihood with a quasi-likelihood). SAIGE tends to have similar power to REGENIE, because both methods assume an infinitesimal prior distribution for SNP effect sizes (and because the difficulty of constructing accurate PGS for binary phenotypes means that all binary MMAA tools tend to have similar power). However, SAIGE tends to be substantially slower than REGENIE, reflecting that its algorithms are based on those used by BOLT-LMM.

When running Step 1 of SAIGE, we used the LOCO option.

### Supplementary Note 8: Attempts to estimate $\lambda$

As explained in Supplementary Note 2, the Grammar-Gamma formula provides a way to estimate $\lambda$ when performing two-step MMAA using ridge regression PGS[19]. However, we are not aware of a corresponding formula for the general case (e.g., when using elastic net PGS). Although BOLT-LMM proposed estimating $\lambda$ based on LDSC, Supplementary Figures 29 & 30 show that this approach can result in substantially-inflated test statistics.

The following paragraphs summarize eight attempts we made to estimate $\lambda$. Note that they use $U_j$ and $T_j$ to denote $\chi^2(1)$ test statistics obtained by regressing $Y - P_c$ on $X_j$, where $P_c$ is either a LOCO ridge regression PGS ($U_j$) or a LOCO elastic net PGS ($T_j$). Further, they use $\lambda'$ to denote the estimated test statistic scaling factor corresponding to $T_j$ (obtained using the Grammar-Gamma formula[19]).

**1 - Estimating $\lambda$ using SumHer.** We tried modifying the approach proposed by BOLT-LMM, so that instead of measuring confounding using LDSC (which assumes $\alpha = -1$), we used our software SumHer (which allows the user to specify $\alpha$)[38]. We therefore ran SumHer twice, first using the scaled test statistics $\lambda' T_1, \ldots, \lambda' T_m$, then using the unscaled test statistics $U_1, \ldots, U_m$. For both runs, we used the LDAK-KVIK estimate of $\alpha$ (obtained in Operation 1b). Finally, we set $\lambda = A_1/A_2$, where $A_1$ and $A_2$ were the estimated intercepts from the two runs of SumHer.

Although we found improvement using SumHer instead of LDSC (in particular, it greatly reduced the inflation of test statistics when analyzing phenotypes generated assuming $\alpha = -0.25$), our modified approach still appeared to produce biased test statistics. Additionally, we had two further concerns with this approach. Firstly, the estimates of confounding (both from LDSC and SumHer) are based on strong assumptions regarding how causal variation is distributed across the genome and the impact of confounding on test statistics, but it is challenging to test the validity of these assumptions on real data. Secondly, this approach can only be applied to datasets with linkage disequilibrium (i.e., where nearby predictors tend to be strongly correlated), whereas we wanted an approach that could be universally applied.

**2 - Estimating $\lambda$ based on the test statistics of weakly-associated SNPs.** This is our chosen approach when analyzing quantitative phenotypes for datasets with high structure. It is similar to Approach 1, except that it measures confounding based on the average test statistic of SNPs weakly-associated with the phenotype (see Supplementary Note 2 for a full description).

**3 - Estimating $\lambda$ based on the test statistics of strongly-associated SNPs.** When using a two-step MMAA tool, the expected increase of the average $\chi^2(1)$ test statistic of associated SNPs is proportional to 1/MSE, where MSE is the average mean-squared error of the Step 1 LOCO PGS. This motivated us to set $\lambda$ so that $\sum \lambda U_j / \sum \lambda' T_j = M_1/M_2$, where the summations are strongly-associated SNPs (e.g., SNPs with $\lambda' T_j > 30$), while $M_1$ and $M_2$ are, respectively, the estimated MSE of the LOCO ridge regression and elastic net PGS (obtained from the cross-validation performed in Operation 1d). We found this approach performed poorly, reflecting the large variance of test statistics (e.g., the standard deviations of test statistics with expectation 30 and 50 are approximately 11 and 14, respectively).

**4 - Generalized least-squares regression with mixture priors** Suppose we have only two SNPs, and test SNP 1 for association using the model $Y = X_1\beta_j + X_2\gamma_2 + e$, with $\gamma_2 \sim N(0, \hat{h}^2)$ and $e_i \sim N(0, 1-\hat{h}^2)$. This would be equivalent to assuming the model

$$Y \sim N(X_1\beta_1, \hat{V}) \quad \text{with} \quad \hat{V} = K\hat{h}^2 + I(1-\hat{h}^2) \quad \text{where} \quad K = X_2X_2^T.$$

The generalized least-squares estimate of $\beta_1$ is

$$\hat{\beta}_1 = \frac{X_1^T\hat{V}^{-1}Y}{X_1^T\hat{V}^{-1}X_1},$$

which is the value of $\beta_1$ that maximizes the model likelihood

$$L(Y|\beta_1) = (2\pi)^{\frac{-(n-q)}{2}} |\hat{V}|^{\frac{-1}{2}} \exp\left(-\frac{(Y - X_1\beta_1)^T\hat{V}^{-1}(Y - X_1\beta_1)}{2}\right).$$

Now suppose we replace the infinitesimal prior distribution for $\gamma_2$ with the mixture prior distribution $\gamma_2 \sim pN(0, h_A^2) + (1-p)N(0, h_A^2)$. This would be equivalent to assuming the model

$$Y \sim pN(X_1\beta_1, \hat{V}_A) + (1-p)N(X_1\beta_1, \hat{V}_B), \quad \text{with} \quad \hat{V}_A = Kh_A^2 + I(1-h^2) \quad \text{and} \quad \hat{V}_B = Kh_B^2 + I(1-h^2),$$

and the corresponding likelihood takes the form

$$L(Y|\beta_1) = pL_A(Y|\beta_1) + (1-p)L_B(Y|\beta_1),$$

where $L_A(Y|\beta_1)$ and $L_B(Y|\beta_1)$ are the likelihoods when assuming $Y \sim N(X_1\beta_1, \hat{V}_A)$ and $Y \sim N(X_1\beta_1, \hat{V}_B)$, respectively. One way to estimate $\beta_1$ would be to maximize the revised model likelihood, which could be considered the generalized least-squares estimate assuming a mixture prior. If we did this directly, by differentiating the model likelihood with respect to $\beta_1$ and setting to zero, it would be necessary to solve

$$-X_1^T\hat{V}_A^{-1}(Y - X_1\beta_1) \times pL_A(Y|\beta_1) - X_1^T\hat{V}_B^{-1}(Y - X_1\beta_1) \times (1-p)L_B(Y|\beta_1) = 0.$$

It is not obvious how to find $\beta_1$. Furthermore, consider that this is the simplest case, because there are only two SNPs, and hence only one $\gamma_j$; for a proper application, there would be approximately $m$ $\gamma_j$, and the model likelihood (and its derivative) would have approximately $2^m$ terms. We wondered whether it would be feasible to replace the (true) model likelihood with the approximate likelihood from variational Bayes. However, by our calculations, this would be equivalent to assuming $Y - P_c \sim N(X_1\beta_1, I(1-h^2))$, and therefore very similar to performing classical linear regression.

**5 - Finding an equivalent ridge regression PGS.** As explained in Supplementary Note 2, a ridge regression PGS is obtained by assuming prior distributions of the form $\gamma_j \sim N(0, a_j)$ and $e_i \sim N(0, \sigma_e^2)$. We noticed that, given an arbitrary LOCO elastic net PGS $P_c$, our variational Bayes solver could find values for $a_j$ and $\sigma_e^2$ such that the resulting ridge regression PGS $P_c'$ was identical to $P_c$. It followed that if $\Omega'$ is an $m \times m$ diagonal matrix with $\Omega'_{j,j} = a_j$, then we could express each LOCO elastic net PGS constructed in Step 1 in the form $P_c = K'V'^{-1}Y$, where $K' = X\Omega'X^T$ and $V' = K' + I\sigma_e^2$. This suggested that we could use the Grammar-Gamma formula, but replacing $V$ with $V'$. Unfortunately, this approach did not work, which we suspect was due to overfitting, a consequence of the fact that the matrix $V'$ was constructed based on the data.

**6 - Estimating $\lambda$ via permutation.** The role of $\lambda$ can be better understood by examining the per-SNP components of the Grammar-Gamma formula

$$\lambda_j' = \frac{X_j^TX_j \times \hat{s}^2}{X_j^T\hat{V}_c^{-1}X_j \times \hat{\sigma}^2}, \quad \text{where} \quad \hat{s}^2 = \frac{Y^T\hat{V}_c^{-1}\hat{V}_c^{-1}Y}{n-q} \quad \text{and} \quad \hat{\sigma}^2 = \frac{Y^T\hat{V}_c^{-1}Y}{n-q}.$$

Note that this version is slightly different to that in Supplementary Note 2, because we previously ignored the LOCO aspect.

We believe that $\lambda$ has two main purposes. The first is to quantify the overfitting due to correlation between SNPs on different chromosomes, while the second is to quantify the the overfitting inherent in the LOCO PGS. Both these purposes are captured by the denominator term $X_j^TV_c^{-1}X_j$. To appreciate why, note that $V_c^{-1}X_j$ is proportional to the vector of residuals when constructing a ridge regression PGS for $X_j$ using SNPs on other chromosomes (Supplementary Note 4), and therefore its magnitude is a measure of overfitting.

We therefore considered computing $\lambda_j = G_j^TV'^{-1}G_j/X_j^TV'^{-1}X_j$, where $G_j$ is a permutation of $X_j$ and $V'$ is the variance matrix corresponding to the "equivalent ridge regression PGS" (explained in Approach 5). Specifically, our proposed estimate of $\lambda$ was the mean value of $\lambda_j$ computed for 20 randomly-picked SNPs. Our idea was that the numerator $X_j^TV'^{-1}X_j$ will be affected by both overfitting due to cross-chromosome correlations and inherent overfitting, whereas the denominator $G_j^TV'^{-1}G_j$ will only be affected by inherent overfitting (because permuting

SNP $j$ will ensure this SNP is uncorrelated with SNPs on other chromosomes). It follows that the ratio of these two terms will measure only overfitting due to cross-chromosome correlations.

We found that this approach performed well for the homogeneous dataset (in which case cross-chromosome correlations are slight, and $\lambda_j$ is close to one). However, it only partially worked for the family and twins datasets (in general, we found it underestimated $\lambda$ by 10-20%).

**7 - Combining pedigree and LOCO PGS.** We wondered if it is possible to first construct $P_K$, a pedigree PGS (copying the approach of fastGWA), then to construct the LOCO elastic net PGS using the offset phenotype $Y - P_K$. Further, we speculated that the test statistic scaling factor corresponding to regressing $Y - P_K - P_c$ on $X_j$, where $P_c$ is the LOCO PGS corresponding to SNP $j$, takes the form $\lambda = \lambda_1 \times \lambda_2$, where $\lambda_1$ and $\lambda_2$ are the scaling factors corresponding to the pedigree and LOCO PGS, respectively. $\lambda_1$ can be estimated via the Grammar-Gamma formula[19], while we thought it might be valid to set $\lambda_2 = 1$, due to that fact that $Y - P_K$ has been "corrected for structure." This approach did not work, which we believe was due to overfitting when constructing $P_K$, which resulted in $Y - P_K$ having negative estimated heritability (making it challenging to construct meaningful LOCO PGS). More formally, this approach corresponds to assuming the model $Y = X_j\beta_j + P_K + P_c + e$, which suggests that it is necessary to construct $P_K$ and $P_c$ jointly (instead of first constructing $P_K$, then constructing $P_c$ using the offset phenotype $Y - P_K$).

**8 - Setting $\lambda = 1$.** This is our chosen approach when analyzing quantitative phenotypes for datasets with low structure. As explained above, we view $\lambda$ as a measure of overfitting, and therefore we believe its true value is upper-bounded by one (because we do not see how negative overfitting is possible). While setting $\lambda = 1$ can lead to deflated test statistics (when the true value is below one), we believe it should not lead to inflated test statistics (because the true value is never above one).

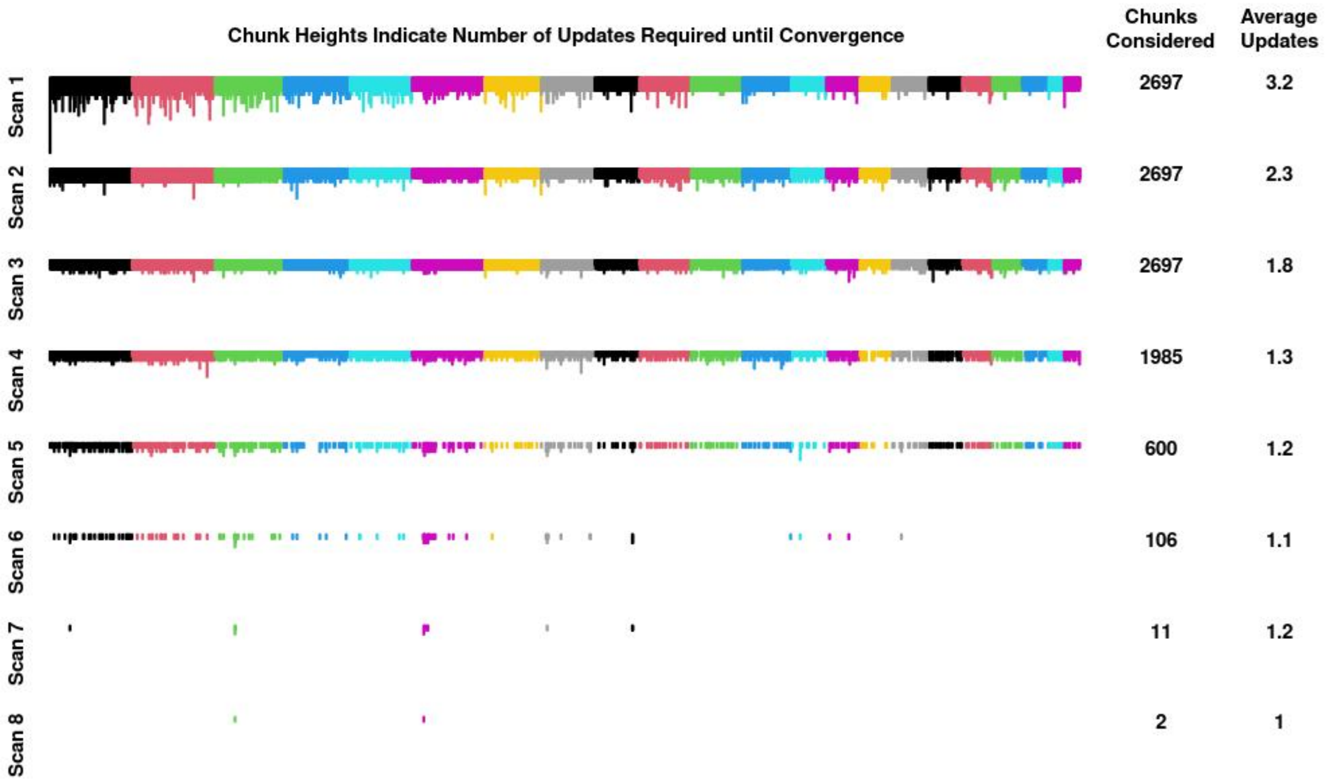

**Supplementary Figure 1: Illustration of our chunk-based variational Bayes solver.** We use variational Bayes to obtain $Q(\gamma) = \prod Q_j(\gamma_j)$, an approximation of the posterior distribution of SNP effect sizes. The variational Bayes solver contained within BOLT-LMM[1] uses sequential genome-wide scans. On Scan 1, it visits each SNP once (i.e., update $Q_1(\gamma_1)$, then $Q_2(\gamma_2)$, and so on, until $Q_m(\gamma_m)$). Then it repeats this process on subsequent scans (i.e., visits each SNP once), continuing until the (genome-wide) approximate log likelihood has converged. By contrast, our solver divides the genome into chunks of 256 SNPs, then uses chunk-based scans. On Scan 1, it first only updates $Q_j(\gamma_j)$ for SNPs in Chunk 1, continuing until the (chunk-based) approximate log likelihood for Chunk 1 has converged. It then moves on to SNPs in Chunk 2 (continuing until convergence), then to SNPs in Chunk 3, and so on until it reaches the final chunk in the genome. On subsequent scans, our solver only visits chunks that had a sizeable impact on the approximate log likelihood in the previous scan.

This figure shows a scenario where there are 2697 chunks. On Scan 1 our solver visits all chunks, and the same for Scans 2 & 3. However, on Scan 4, it only visits 1985 chunks, indicating that the remaining 712 achieved convergence on Scan 3. We see that by the end of Scan 8, all chunks have converged, so our solver stops. Within each scan, the height of each chunk indicates the number of updates required until each convergence, while the numbers in the right column record the average number of updates across all active chunks. For example, we see that on Scan 5, the average number of updates was 1.2 (i.e., for the approximately 153 k SNPs contained within the 600 chunks visited on this scan, it was on average necessary to update $Q_j(\gamma_j)$ 1.2 times).

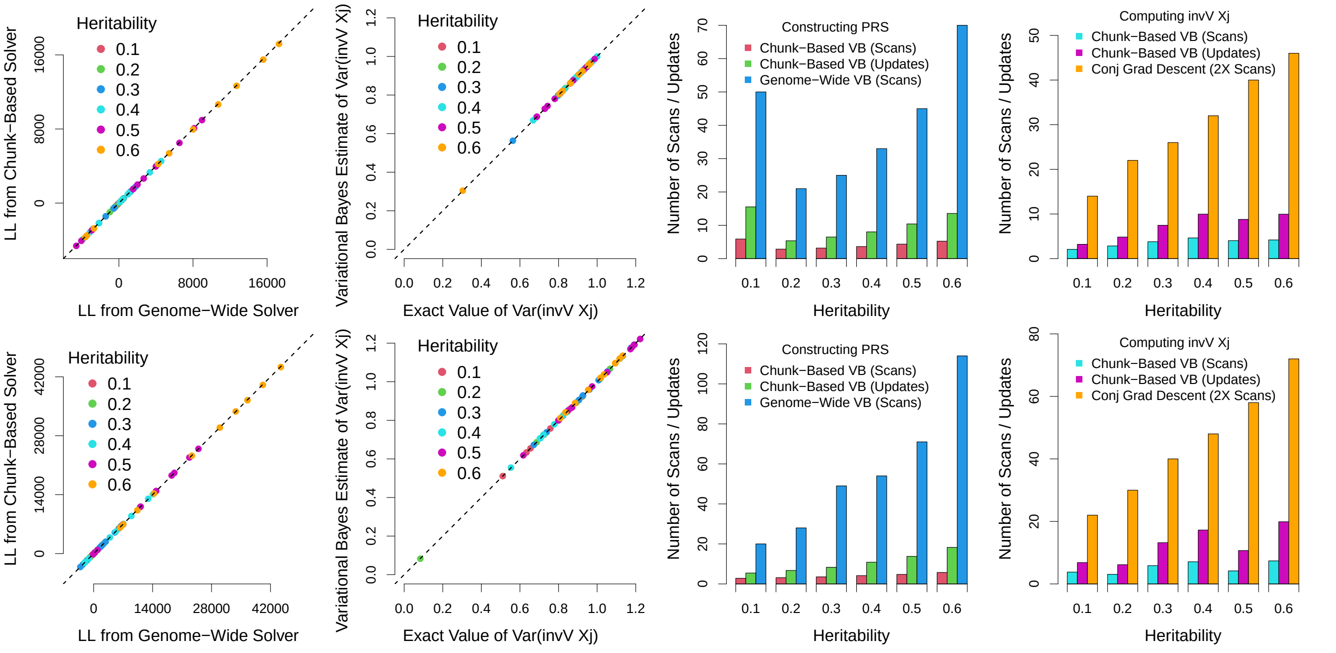

**Supplementary Figure 2: Accuracy and speed of our variational Bayes solver.** For this figure, we generate six phenotypes each with 10 000 causal SNPs, and with heritabilities ranging from 0.1 to 0.6. The top panels report results from analyzing 50 k individuals, while the bottom panels report results from analyzing 100 k individuals. The first column shows that when constructing elastic net PGS, the approximate log likelihood (LL) from our chunk-based variational Bayes solver matches very closely that from a conventional, genome-wide solver. Note that for each phenotype, there are 10 points, corresponding to ten different sets of elastic net hyperparameters. Next we compute $V^{-1}X_j$ for twenty randomly-picked SNPs, where $V$ is the (phenotype-specific) estimated variance matrix. The second column shows that the variances of the estimates of $V^{-1}X_j$ from our variational Bayes solver are very close to the true values (which we computed using conjugate gradient descent).

The third column reports the numbers of scans and per-SNP updates required by our chunk-based variational Bayes solver to construct the elastic net PGS for each phenotype, as well as the number of scans required by a genome-wide variational Bayes solver. For example, the first bar in the top panel shows that when analyzing the phenotype with heritability 0.1, our solver requires 5.9 scans. Note that this is not an integer, because our solver can perform partial scans (e.g., it may have been the case that our solver performed five scans that visited all chunks, then two scans that visited only 45% of chunks). The second bar shows that our solver performed 15.5 per-SNP updates (i.e., it updated $Q_j(\gamma_j)$ on average 15.5 times for each SNP). The third bar shows that the genome-wide solver required 50 scans (as each scan updates $Q_j(\gamma_j)$ once per SNP, this means the genome-wide solver required about three times more updates than our chunk-based solver). The fourth column is similar to the third, except it reports the numbers of scans and per-SNP updates our solver required when computing $V^{-1}X_j$, and compares this to twice the number of scans required when using conjugate gradient descent (we report twice the number of scans because each conjugate gradient descent scan performs approximately twice the number of algebraic operations as each scan of our variational Bayes solver).

In summary, we found that our chunk-based variational Bayes solver constructs PGS that match very closely those computed by a genome-wide variational Bayes solver, but requires substantially fewer updates of $Q_j(\gamma_j)$. We additionally found that our solver produces accurate estimates of $V^{-1}X_j$, and that it does so substantially faster than conjugate gradient descent.

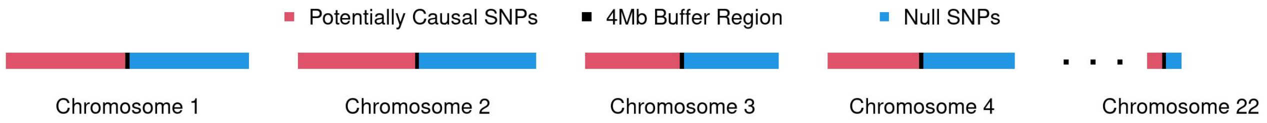

**Supplementary Figure 3: Simulating phenotypes.**

We partitioned the genome into three, as illustrated above. SNPs within the central 4 Mb of each chromosome were assigned to the black partition, while the SNPs upstream and downstream were assigned to the red and blue partitions, respectively. When simulating phenotypes, we ensured that all causal SNPs were located within the red partition (46% of SNPs), then used SNPs within the blue partition (51% of SNPs) for measuring type 1 error (i.e., these were considered "null SNPs"). We included the black partition (3% of SNPs) as a buffer, to ensure the null and causal SNPs were in approximate linkage equilibrium.

We assess the type 1 error of different MMAA tools based on four measures: the mean $\chi^2(1)$ test statistic across null SNPs, and the proportions of null SNPs with $p$-value below 0.05, 0.001 or $5 \times 10^{-5}$. We derived 95% confidence intervals by performing classical single-SNP analysis for 1000 non-heritable phenotypes (we sampled these phenotypes from a normal distribution, independently of genotypes and covariates). The confidence intervals vary slightly by dataset, such that the intervals for the family and twins datasets are approximately 10% and 50% wider than those for the homogeneous dataset. However, for simplicity, our figures only mark the 95% confidence intervals for the homogeneous dataset (which are approximately 92% and 80% confidence intervals for the family and twins dataset, respectively). When testing if a tool has significant inflation, we assume the mean test statistic across null SNPs has the null distribution $\Gamma(N_e/2, 2/N_e)$, where $N_e$ is the effective number of independent null SNPs. We estimate $N_e$ based on the variance of the mean test statistic when analyzing the non-heritable phenotypes (each of the homogeneous, family and twins datasets has 354 k null SNPs, and the estimates of $N_e$ are 79.9 k, 65.2 k and 35.2 k, respectively). Using the significance threshold $P < 0.05/36$ (we divide by 36 because our main simulations construct 12 sets of phenotypes for each of three datasets), a tool has significant inflation if the mean test statistic across null SNPs, when averaged across ten replicates, is above 1.0047 (homogeneous dataset), above 1.0052 (family dataset) or above 1.0071 (twins dataset). We recognize that our significance testing is not exact. For example, our choice of significance threshold ($P < 0.05/36$) is somewhat arbitrary, and should be tool-specific (e.g., increased for BOLT-LMM, because we only apply this tool to quantitative phenotypes). Furthermore, we appreciate that we may have uncovered more scenarios where a tool was significantly inflated had we used more than ten replicates.

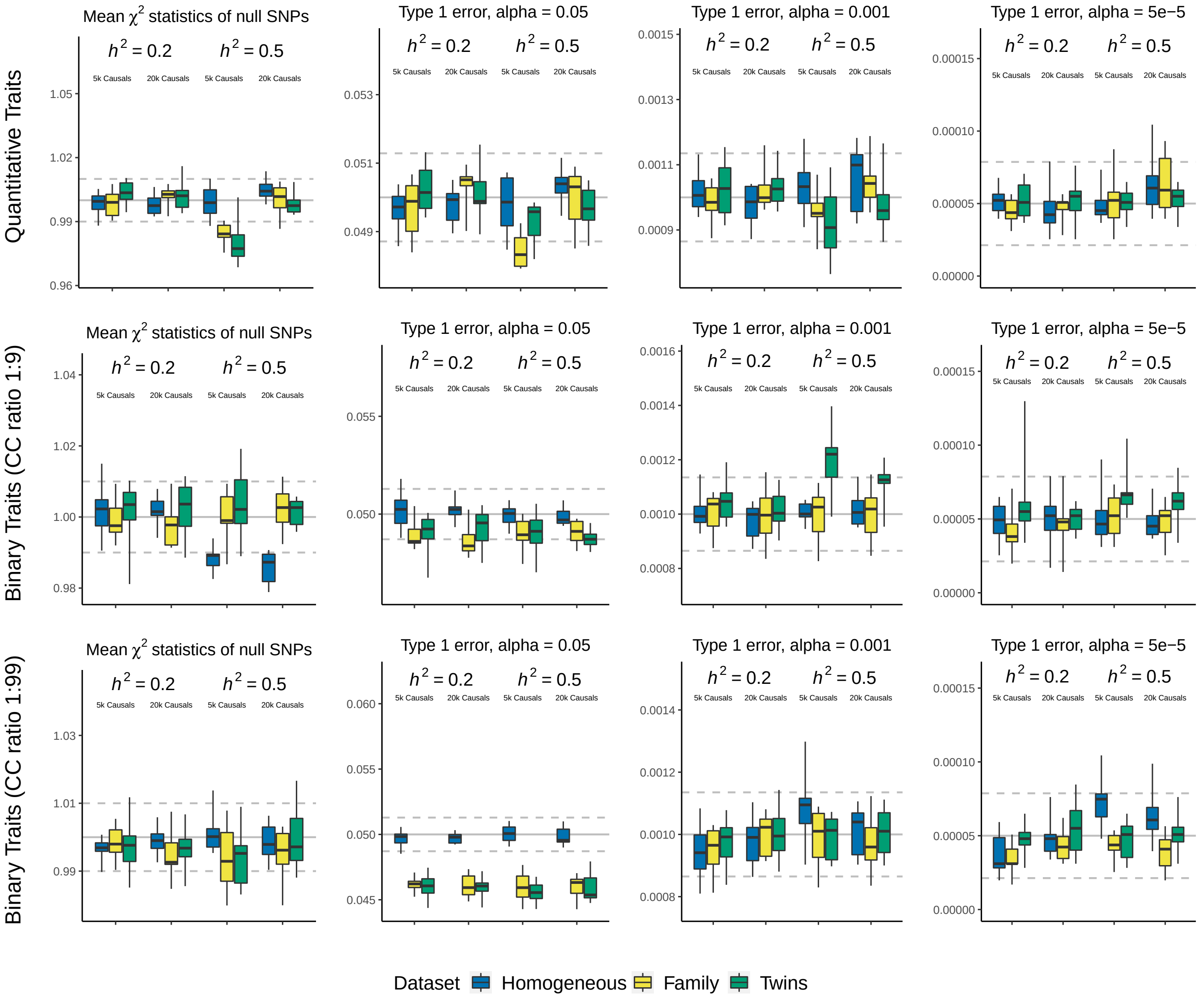

**Supplementary Figure 4: Type 1 error of LDAK-KVIK.**

We simulate quantitative and binary phenotypes for the homogeneous, family and twins datasets (each of which contains 63 k individuals). For each dataset, we consider 12 different scenarios, obtained by varying the heritability (0.2 or 0.5), the number of causal SNPs (5 k or 20 k), and for binary phenotypes, also the prevalence (10% or 1%). When generating causal SNP effect sizes, we assume $\alpha = -1$. We perform single-SNP analysis using LDAK-KVIK, then measure the type 1 error based on the mean $\chi^2(1)$ test statistic of null SNPs (Column 1), and based on the proportions of null SNPs with $p$-value below 0.05, 0.001 and $5 \times 10^{-5}$ (Columns 2, 3 & 4, respectively). In each box, the three horizontal lines mark the median and inter-quartile range across ten replicates. The solid grey lines mark the expected value of each measure under the null hypothesis, while the dashed grey lines provide a 95% confidence interval (derived by analyzing 1000 permuted phenotypes, as explained in Supplementary Figure 3).

We find that LDAK-KVIK controls type 1 error for all datasets and scenarios considered. For example, the highest mean test statistic of null SNPs, averaged across ten replicates, is 1.0049, which occurs when analyzing the quantitative phenotypes with heritability 0.5 and 20 k causal SNPs for the homogeneous dataset (the tenth box in the first panel); the corresponding one-sided $p$-value is 0.002, which is not significant when allowing for multiple testing (see Supplementary Figure 3 for an explanation of how we obtained $p$-values, and our choice of significance threshold).

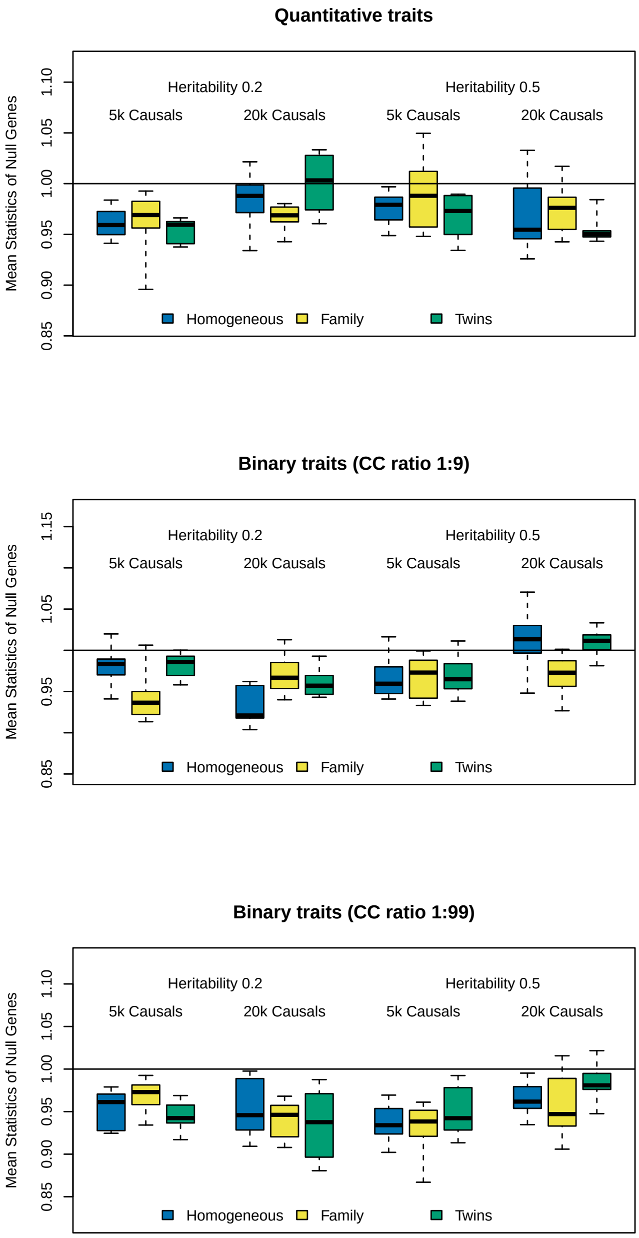

**Supplementary Figure 5: Type 1 error of LDAK-KVIK-GBAT.**
We simulate quantitative and binary phenotypes for the homogeneous, family and twins datasets (each of which contains 63 k individuals). For each dataset, we consider 12 different scenarios, obtained by varying the heritability (0.2 or 0.5), the number of causal SNPs (5 k or 20 k), and for binary phenotypes, also the prevalence (10% or 1%). When generating causal SNP effect sizes, we assume $\alpha = -1$. We perform gene-based analysis using LDAK-KVIK-GBAT, then measure the type 1 error based on the mean $\chi^2(1)$ test statistic of null genes. In each box, the three horizontal lines mark the median and inter-quartile range across ten replicates. The solid grey lines mark the expected mean $\chi^2(1)$ test statistic under the null hypothesis.

We find that LDAK-KVIK-GBAT has well-controlled type 1 error for all datasets and scenarios considered (although we recognise that its test statistics can be deflated, which is because the algorithm LDAK-GBAT uses for estimating the null distribution is slightly conservative[22]).

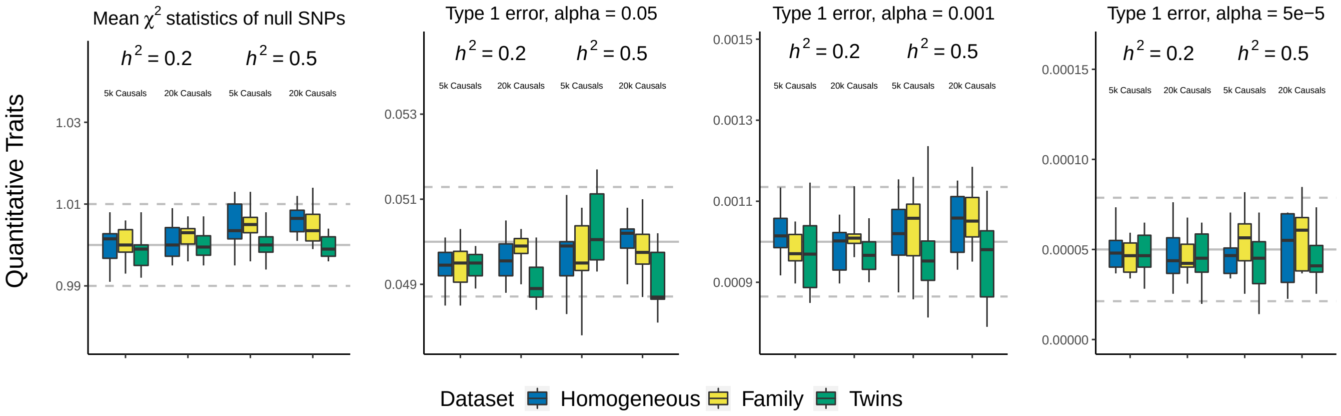

**Supplementary Figure 6: Type 1 error of BOLT-LMM.**

We simulate quantitative phenotypes for the homogeneous, family and twins datasets (each of which contains 63 k individuals). For each dataset, we consider 4 different scenarios, obtained by varying the heritability (0.2 or 0.5) and the number of causal SNPs (5 k or 20 k). When generating causal SNP effect sizes, we assume $\alpha = -1$. We perform single-SNP analysis using BOLT-LMM, then measure the type 1 error based on the mean $\chi^2(1)$ test statistic of null SNPs (Column 1), and based on the proportions of null SNPs with $p$-value below 0.05, 0.001 and $5 \times 10^{-5}$ (Columns 2, 3 & 4, respectively). In each box, the three horizontal lines mark the median and inter-quartile range across ten replicates. The solid grey lines mark the expected value of each measure under the null hypothesis, while the dashed grey lines provide an approximate 95% confidence interval (derived by analyzing 1000 permuted phenotypes, as explained in Supplementary Figure 3).

While this figure shows that BOLT-LMM has well-controlled type 1 error for all datasets and scenarios considered, this reflects that we have restricted to phenotypes simulated with $\alpha = -1$ (matching the assumption of BOLT-LMM). In Supplementary Figure 29, we demonstrate that BOLT-LMM can produce substantially-inflated test statistics when applied to phenotypes simulated with $\alpha = -0.25$, which more closely reflects what we observe for real human traits.

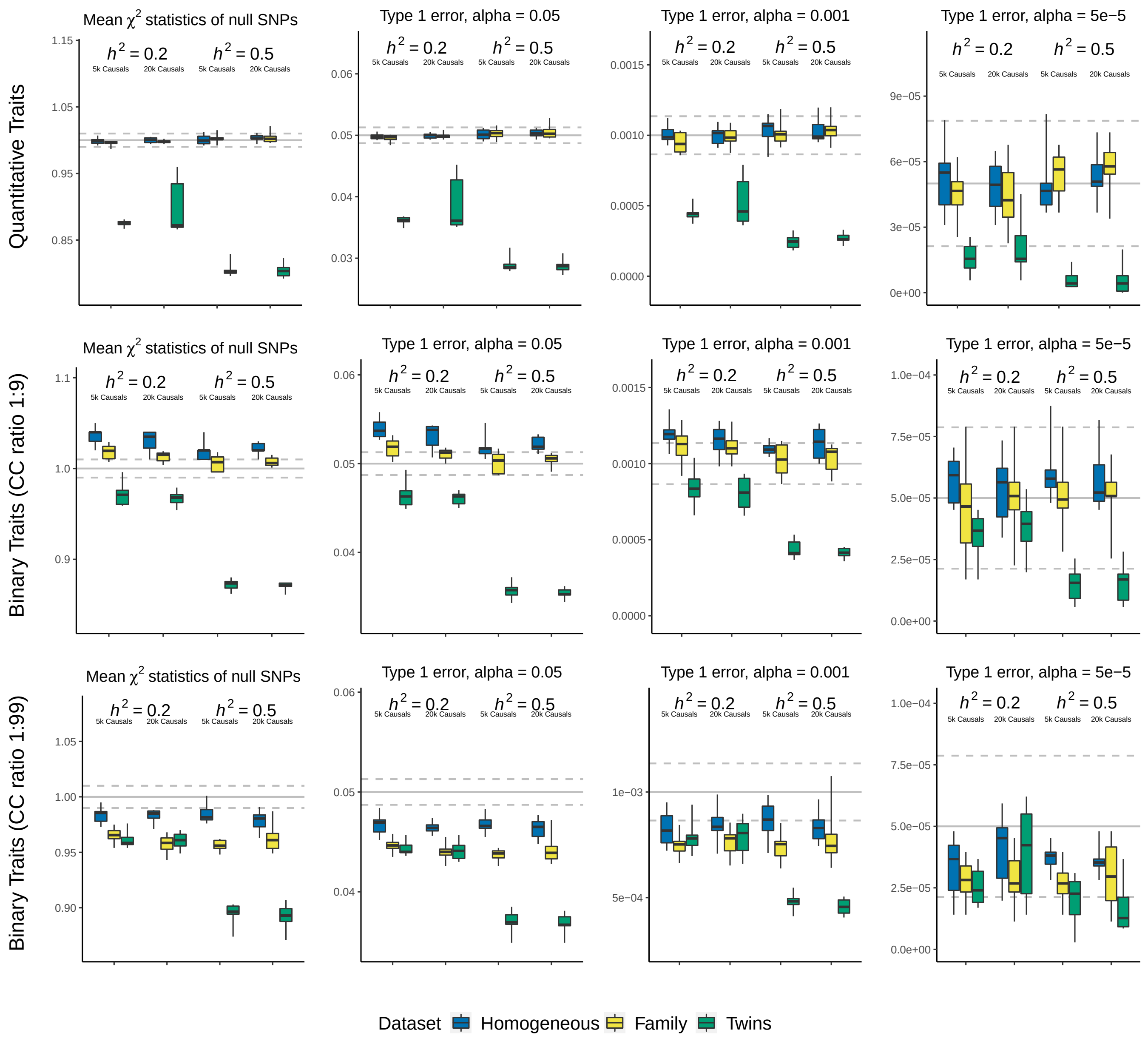

**Supplementary Figure 7: Type 1 error of REGENIE.**

We simulate quantitative and binary phenotypes for the homogeneous, family and twins datasets (each of which contains 63 k individuals). For each dataset, we consider 12 different scenarios, obtained by varying the heritability (0.2 or 0.5), the number of causal SNPs (5 k or 20 k), and for binary phenotypes, also the prevalence (10% or 1%). When generating causal SNP effect sizes, we assume $\alpha = -1$. We perform single-SNP analysis using REGENIE, then measure the type 1 error based on the mean $\chi^2(1)$ test statistic of null SNPs (Column 1), and based on the proportions of null SNPs with $p$-values below 0.05, 0.001 and $5 \times 10^{-5}$ (Columns 2, 3 & 4, respectively). In each box, the three horizontal lines mark the median and inter-quartile range across ten replicates. The solid grey lines mark the expected value of each measure under the null hypothesis, while the dashed grey lines provide a 95% confidence interval (derived by analyzing 1000 permuted phenotypes, as explained in Supplementary Figure 3).

We find that for quantitative phenotypes and rare binary phenotypes, REGENIE controls type 1 error for all datasets and scenarios considered. However, we see that REGENIE tends to produce inflated test statistics when applied to common binary phenotypes. We also note that REGENIE can have deflated test statistics when applied to datasets containing related samples, which is a consequence of it (implicitly) assuming $\lambda = 1$.

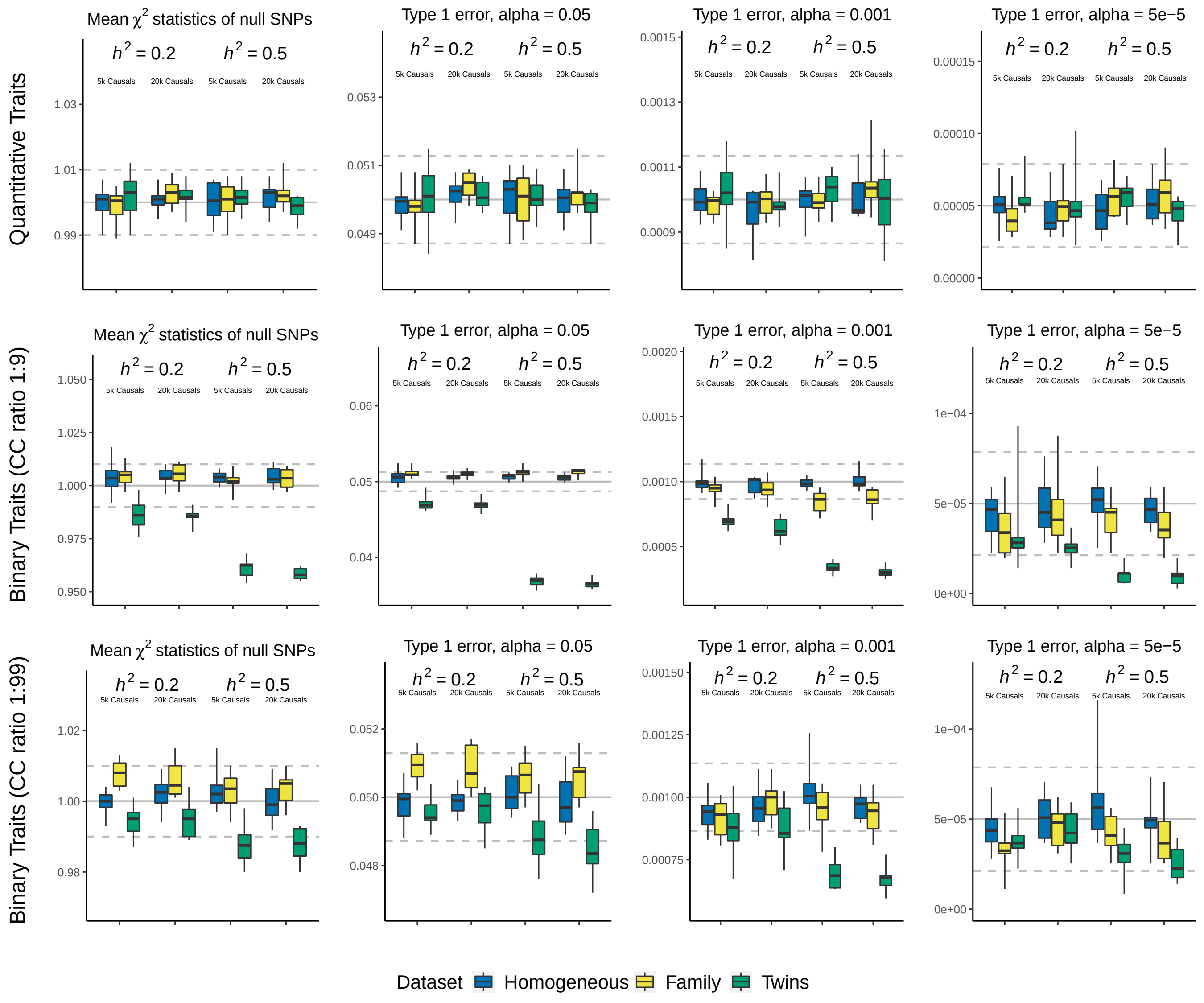

**Supplementary Figure 8: Type 1 error of fastGWA.**

We simulate quantitative and binary phenotypes for the homogeneous, family and twins datasets (each of which contains 63 k individuals). For each dataset, we consider 12 different scenarios, obtained by varying the heritability (0.2 or 0.5), the number of causal SNPs (5 k or 20 k), and for binary phenotypes, also the prevalence (10% or 1%). When generating causal SNP effect sizes, we assume $\alpha = -1$. We perform single-SNP analysis using fastGWA, then measure the type 1 error based on the mean $\chi^2(1)$ test statistic of null SNPs (Column 1), and based on the proportions of null SNPs with $p$-values below 0.05, 0.001 and $5 \times 10^{-5}$ (Columns 2, 3 & 4, respectively). In each box, the three horizontal lines mark the median and inter-quartile range across ten replicates. The solid grey lines mark the expected value of each measure under the null hypothesis, while the dashed grey lines provide a 95% confidence interval (derived by analyzing 1000 permuted phenotypes, as explained in Supplementary Figure 3).

We find that fastGWA has well-controlled type 1 error for all datasets and scenarios considered (although we note that its test statistics can be deflated when analyzing common binary phenotypes for datasets with high relatedness).

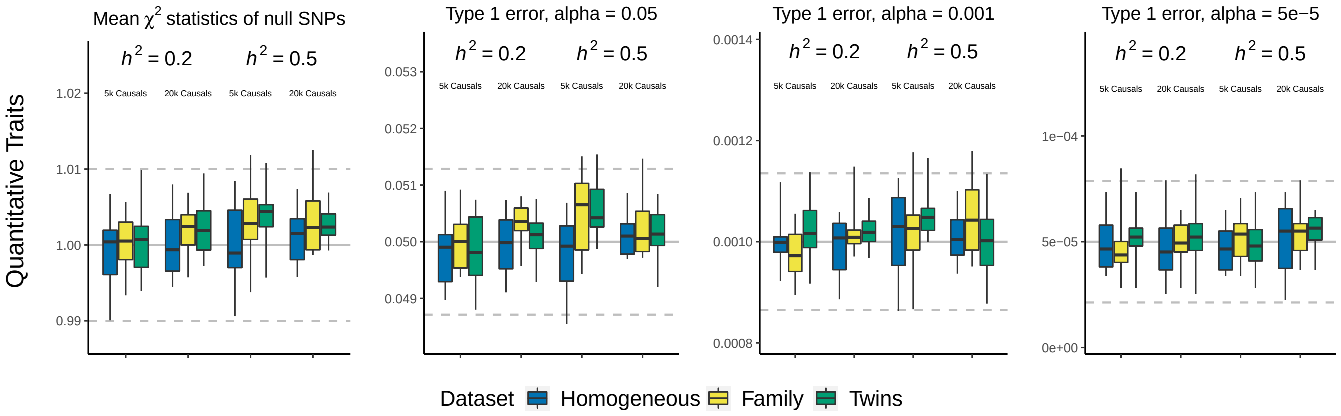

**Supplementary Figure 9: Type 1 error of GCTA-LOCO.**

We simulate quantitative phenotypes for the homogeneous, family and twins datasets (each of which contains 63 k individuals). For each dataset, we consider 4 different scenarios, obtained by varying the heritability (0.2 or 0.5) and the number of causal SNPs (5 k or 20 k). When generating causal SNP effect sizes, we assume $\alpha = -1$. We perform single-SNP analysis using GCTA-LOCO, then measure the type 1 error based on the mean $\chi^2(1)$ test statistic of null SNPs (Column 1), and based on the proportions of null SNPs with $p$-values below 0.05, 0.001 and $5 \times 10^{-5}$ (Columns 2, 3 & 4, respectively). In each box, the three horizontal lines mark the median and inter-quartile range across ten replicates. The solid grey lines mark the expected value of each measure under the null hypothesis, while the dashed grey lines provide an approximate 95% confidence interval (derived by analyzing 1000 permuted phenotypes, as explained in Supplementary Figure 3).

We find that GCTA-LOCO has well-controlled type 1 error for all datasets and scenarios considered.

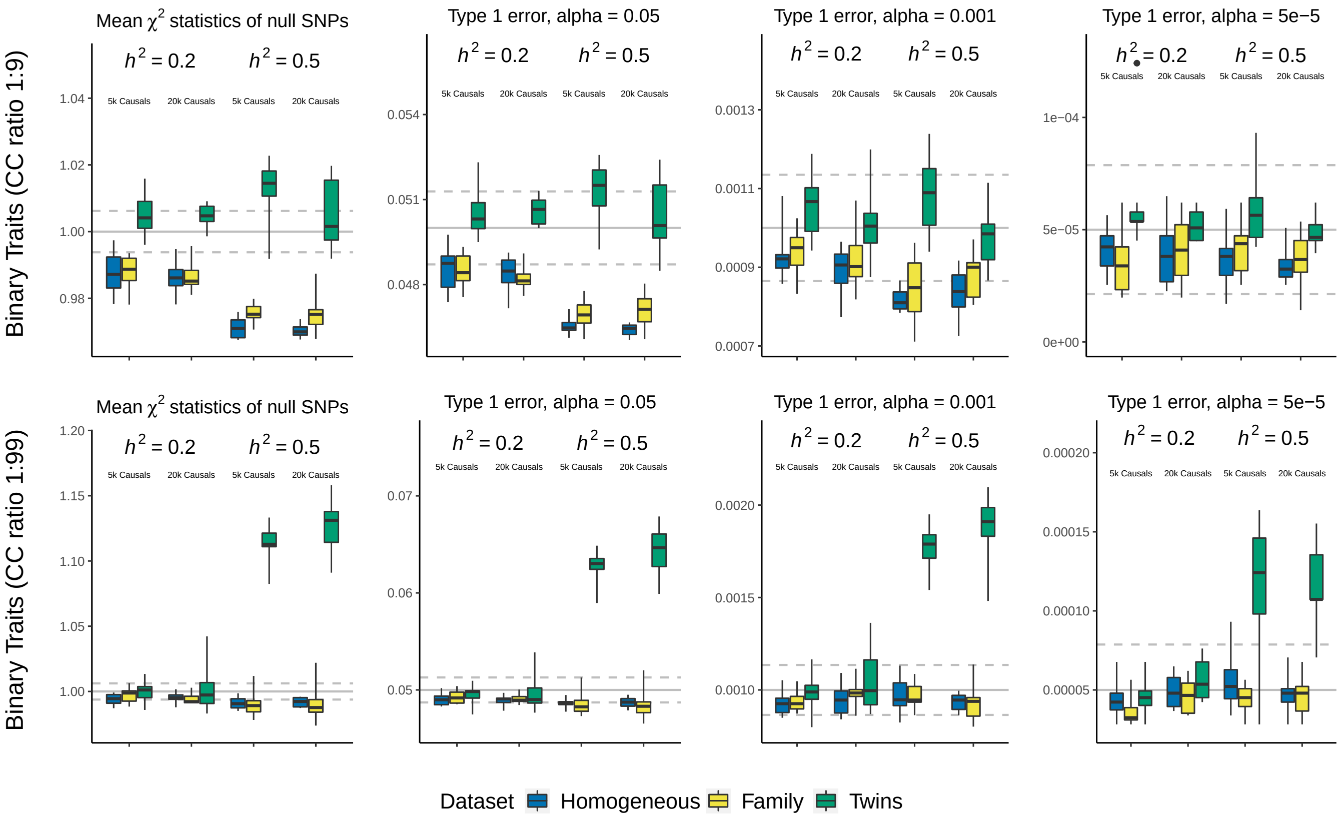

**Supplementary Figure 10: Type 1 error of SAIGE.**

We simulate binary phenotypes for the homogeneous, family and twins datasets (each of which contains 63 k individuals). For each dataset, we consider 8 different scenarios, obtained by varying the heritability (0.2 or 0.5), the number of causal SNPs (5 k or 20 k) and the prevalence (10% or 1%). When generating causal SNP effect sizes, we assume $\alpha = -1$. We perform single-SNP analysis using SAIGE, then measure the type 1 error based on the mean $\chi^2(1)$ test statistic of null SNPs (Column 1), and based on the proportions of null SNPs with $p$-values below 0.05, 0.001 and $5 \times 10^{-5}$ (Columns 2, 3 & 4, respectively). In each box, the three horizontal lines mark the median and inter-quartile range across ten replicates. The solid grey lines mark the expected value of each measure under the null hypothesis, while the dashed grey lines provide an approximate 95% confidence interval (derived by analyzing 1000 permuted phenotypes, as explained in Supplementary Figure 3).

We find that SAIGE generally has well-controlled type 1 error for phenotypes with prevalence 10%. However, we see that for phenotypes with prevalence 1%, SAIGE can produce inflated test statistics when there is both high heritability and relatedness.

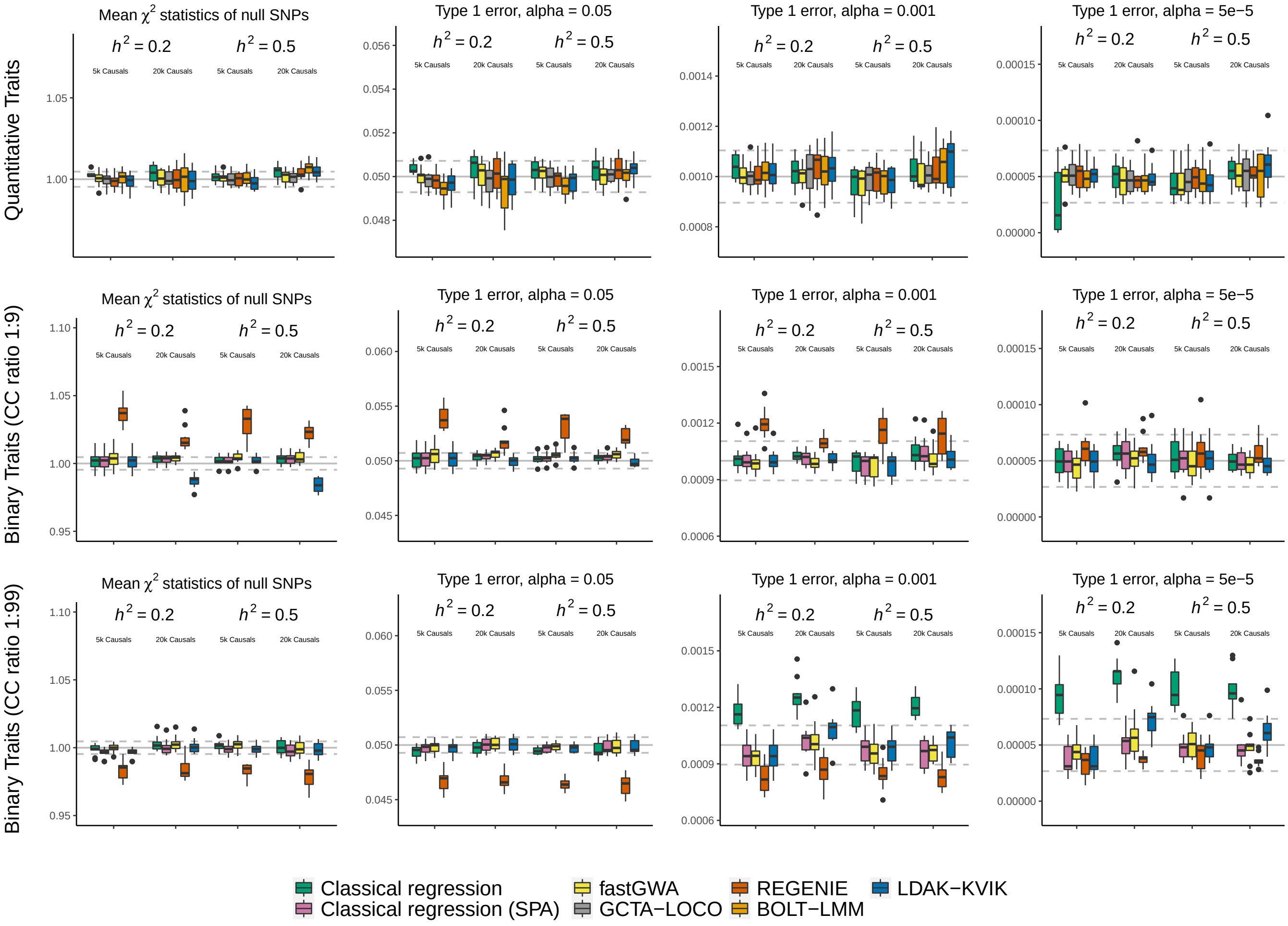

**Supplementary Figure 11: Type 1 error of all MMAA tools for the homogeneous dataset.** We simulate quantitative and binary phenotypes for the homogeneous dataset (63 k individuals). We consider 12 different scenarios, obtained by varying the heritability (0.2 or 0.5), the number of causal SNPs (5 k or 20 k), and for binary phenotypes, also the prevalence (10% or 1%). When generating causal SNP effect sizes, we assume $\alpha = -1$. We perform single-SNP analysis using classical linear regression, classical logistic regression (with and without SPA correction of $p$-values), BOLT-LMM, REGENIE, fastGWA, GCTA-LOCO and LDAK-KVIK, then measure the type 1 error based on the mean $\chi^2(1)$ test statistic of null SNPs (Column 1), and based on the proportions of null SNPs with $p$-values below 0.05, 0.001 and $5 \times 10^{-5}$ (Columns 2, 3 & 4, respectively). In each box, the three horizontal lines mark the median and inter-quartile range across ten replicates. The solid grey lines mark the expected value of each measure under the null hypothesis, while the dashed grey lines provide a 95% confidence interval (derived by analyzing 1000 permuted phenotypes, as explained in Supplementary Figure 3).

This figure is intended to provide a broad comparison of the type 1 error of different MMAA tools when applied to homogeneous data (note that except for the non-SPA logistic regression, all the information in this figure can be seen more clearly by examining the blue boxes in Supplementary Figures 4, 6, 7, 8 & 9). We find that the MMAA tools are generally well-calibrated across the different scenarios, with the notable exceptions being the inflation and deflation observed for REGENIE when analyzing common and rare binary phenotypes, respectively (the red boxes in Rows 2 & 3). Furthermore, the final panel demonstrates the importance of using the SPA when analyzing rare binary phenotypes (the green boxes show that without SPA correction, logistic regression can produce false positives, while the purple boxes indicate that with SPA correction, these are avoided).

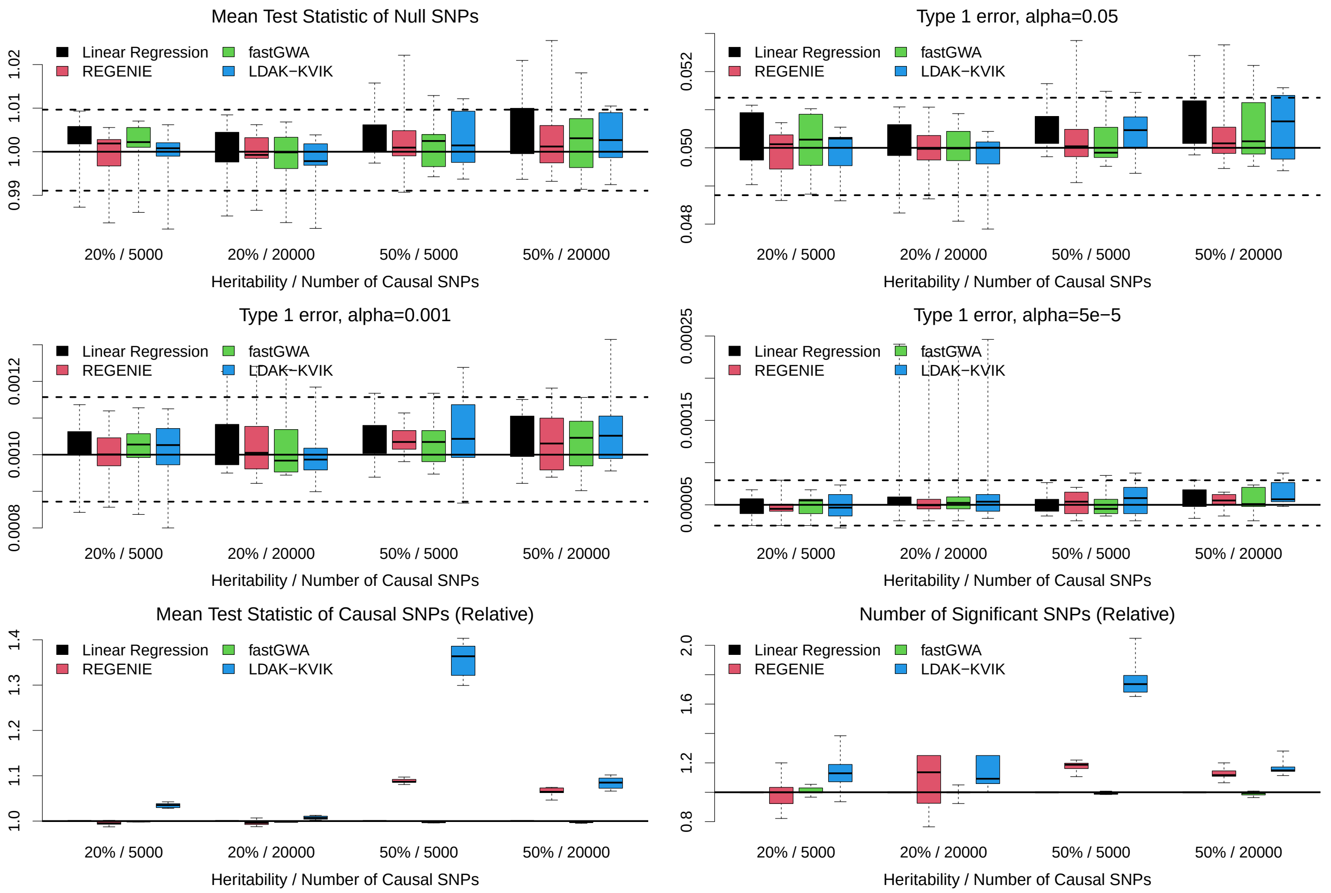

**Supplementary Figure 12: Fine-scale population structure.**

We simulate quantitative phenotypes for the British-Irish dataset (63 k individuals divided into 51 k British and 12 k Irish). For each phenotype, 20% of the total phenotypic variance is explained by a binary vector that indicates which individuals are Irish. We consider 4 different scenarios, obtained by varying the heritability (0.2 or 0.5) and the number of causal SNPs (5 k or 20 k). When generating causal SNP effect sizes, we assume $\alpha = -1$. We perform single-SNP analysis using classical linear regression, REGENIE, fastGWA and LDAK-KVIK. We measure the type 1 error based on the mean $\chi^2(1)$ test statistic of null SNPs (Panel 1), and based on the proportions of null SNPs with $p$-values below 0.05, 0.001 and $5 \times 10^{-5}$ (Panels 2, 3 & 4, respectively), while we measure the power based on the mean $\chi^2(1)$ test statistic of causal SNPs (Panel 5) and the number of causal SNPs with $p$-value below $5 \times 10^{-8}$ (Panel 6). In each box, the three horizontal lines mark the median and inter-quartile range across ten replicates. The solid grey lines in the top four panels mark the expected value of each measure under the null hypothesis, while the dashed grey lines provide an approximate 95% confidence interval (derived by analyzing 1000 permuted phenotypes, as explained in Supplementary Figure 3).

In general, the results are similar to those when analyzing simulated quantitative phenotypes for the homogeneous dataset. For example, we see that LDAK-KVIK has good control of type 1 error for all scenarios considered, and tends to be more powerful than REGENIE and fastGWA.

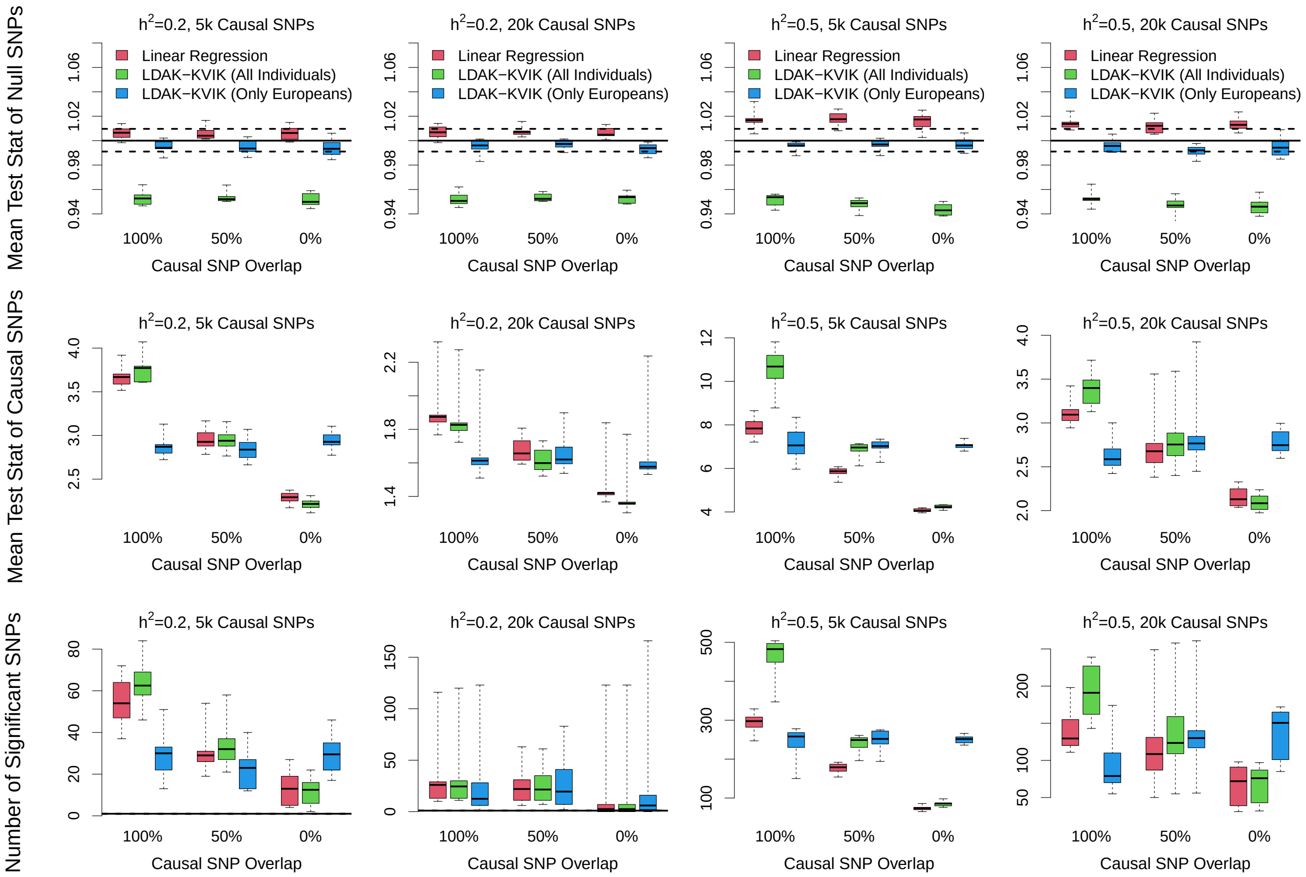

**Supplementary Figure 13: Phenotypes where causal SNPs vary across ancestries.** We simulate quantitative phenotypes for the multi-ancestry dataset (63k individuals, divided into 39 k Europeans, 15 k Asians and 8 k Africans). We consider 12 different scenarios, obtained by varying the heritability (0.2 or 0.5), the number of causal SNPs (5 k or 20 k) and the causal SNP overlap: 100% overlap means the causal SNPs (and their effect sizes) are the same across ancestries, 0% overlap means the causal SNPs are picked independently for Europeans and non-Europeans, while 50% overlap means half the causal SNPs are the same, while half are picked independently. When generating causal SNP effect sizes, we assume $\alpha = -1$. We perform single-SNP analysis using classical linear regression and LDAK-KVIK; for the former we analyze all individuals, while for the latter, we either analyze all individuals or only the European individuals. We measure the type 1 error based on the mean $\chi^2(1)$ test statistic of null SNPs (top row), and measure the power based on the mean $\chi^2(1)$ test statistic of causal SNPs (middle row) or the number of causal SNPs with $p$-value below $5 \times 10^{-8}$ (bottom row). Note that for the power comparisons, we consider only the SNPs causal for European individuals (meaning that for the scenarios with 50% or 0% overlap, we ignore the SNPs that are only causal for non-Europeans). In each box, the three horizontal lines mark the median and inter-quartile range across ten replicates. The solid grey lines in the top row mark the expected mean test statistic under the null hypothesis, while the dashed grey lines provide a 95% confidence interval (derived by analyzing 1000 permuted phenotypes, as explained in Supplementary Figure 3).

We find that LDAK-KVIK controls type 1 error for all scenarios (although we note that its test statistics are well-calibrated when considering only European individuals, but deflated when considering all individuals). As expected, the advantage of analyzing all individuals (instead of only Europeans) depends on the causal SNP overlap; when it is 100%, it is beneficial to consider all individuals, when it is 0%, it is detrimental to include the non-Europeans, while when it is 50%, both sets of analyses give similar results.

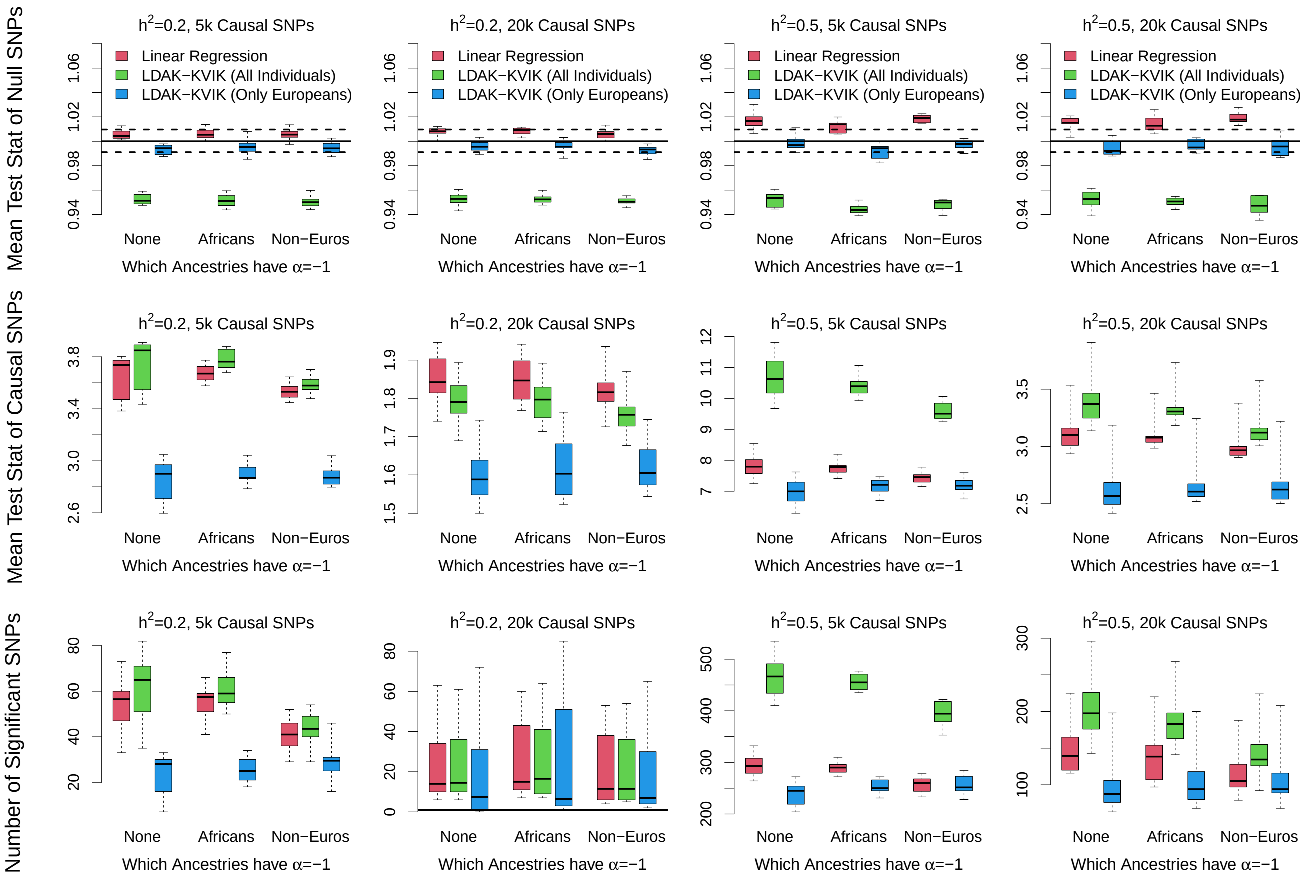

**Supplementary Figure 14: Phenotypes where $\alpha$ varies across ancestries.** We simulate quantitative phenotypes for the multi-ancestry dataset (63k individuals, divided into 39 k Europeans, 15 k Asians and 8 k Africans). We consider 12 different scenarios, obtained by varying the heritability (0.2 or 0.5), the number of causal SNPs (5 k or 20 k) and the choice of $\alpha$ when sampling causal SNP effect sizes: -0.25 for all individuals; -0.25 for Europeans and Asians, but -1 for Africans; -0.25 for Europeans, but -1 for Asians and Africans. We perform single-SNP analysis using classical linear regression and LDAK-KVIK; for the former we analyze all individuals, while for the latter, we either analyze all individuals or only the European individuals. We measure the type 1 error based on the mean $\chi^2(1)$ test statistic of null SNPs (top row), and measure the power based on the mean $\chi^2(1)$ test statistic of causal SNPs (middle row) or the number of causal SNPs with $p$-value below $5 \times 10^{-8}$ (bottom row). In each box, the three horizontal lines mark the median and inter-quartile range across ten replicates. The solid grey lines in the top row mark the expected mean test statistic under the null hypothesis, while the dashed grey lines provide a 95% confidence interval (derived by analyzing 1000 permuted phenotypes, as explained in Supplementary Figure 3).

We find that LDAK-KVIK controls type 1 error for all scenarios (although we note that its test statistics are well-calibrated when considering only European individuals, but deflated when considering all individuals). It is generally more powerful to analyze only European individuals, than to analyze all individuals. However, this likely reflects that we generated effect sizes independently for Europeans, Asians and Africans, meaning that although the same SNPs are causal for all individuals, their effects will often be in opposite directions (we realise it would be more realistic to encourage concordance of effect size direction across ancestries, but could not work out how to do this while allowing $\alpha$ to vary across ancestries).

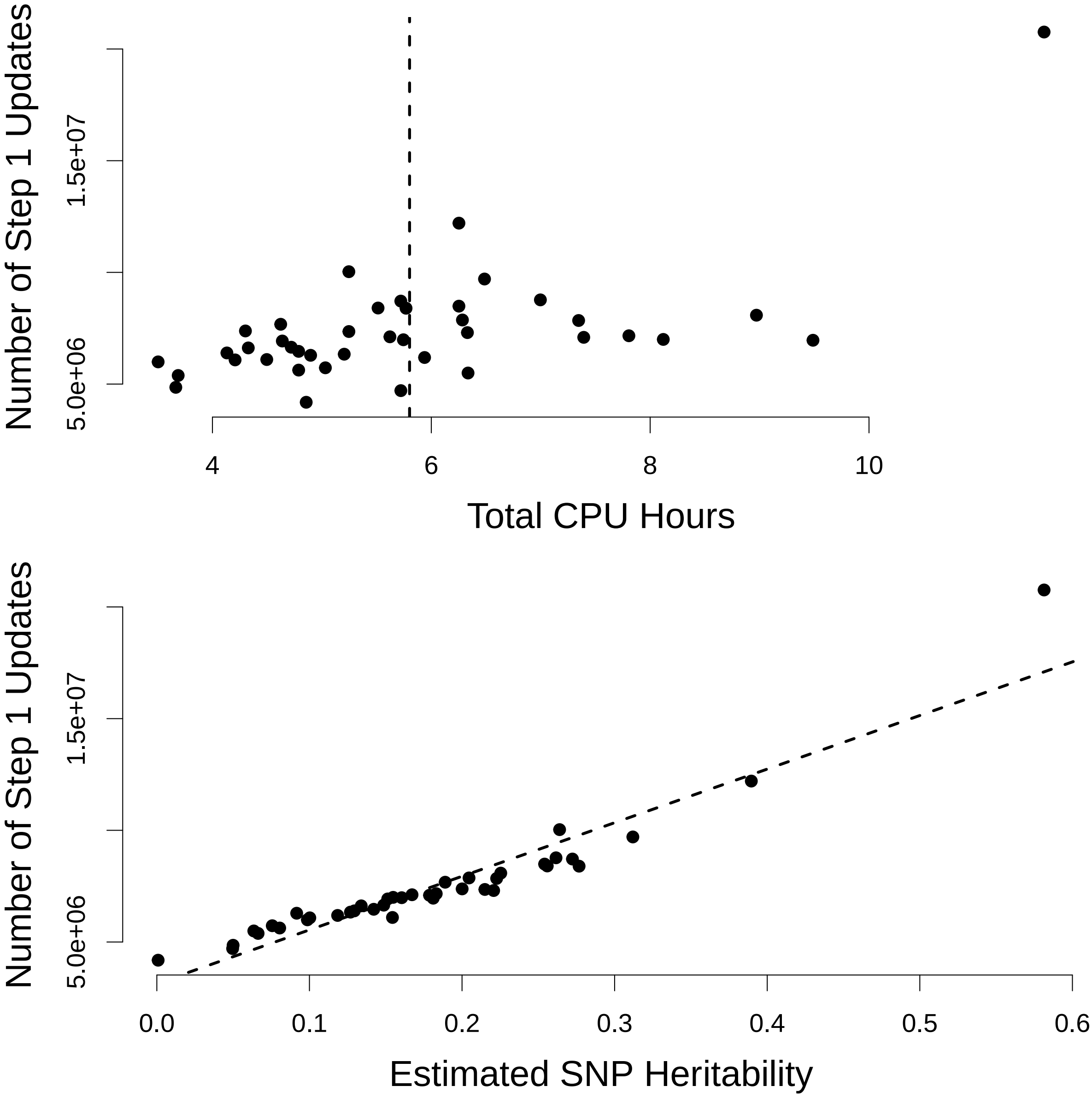

**Supplementary Figure 15: Runtime of LDAK-KVIK when analyzing 368 k individuals.** We analyze the 40 quantitative UK Biobank phenotypes using the white dataset (368 k individuals). The top panel compares the number of variational Bayes updates performed in Step 1 with the total runtime of LDAK-KVIK (i.e., the time taken for both Steps 1 & 2); the dashed vertical line marks the mean runtime across the 40 phenotypes. The bottom panel compares the number of variational Bayes updates performed in Step 1 with the estimated SNP heritability (obtained using our software SumHer[38]); the dashed diagonal line indicates the best fit from regressing the numbers of updates on the estimates of SNP heritability.

Overall, we see that the total runtime of LDAK-KVIK depends strongly on the number of updates our variational Bayes solver requires to achieve convergence, which in turn is strongly correlated with the SNP heritability of the phenotype. This explains why the slowest runtime was recorded for height (whose heritabilty is about 50% higher than the next most heritable phenotype). Note that the average runtime reported here is lower than for Table 1 in the Main Text, which is mainly because the latter considers only five phenotypes (including height, the phenotype with longest runtime), but also reflects random factors (for the analysis in the main text, we ensured all MMAA tools were run on the same processor, whereas here the choice of processor was left to our high-performance cluster).

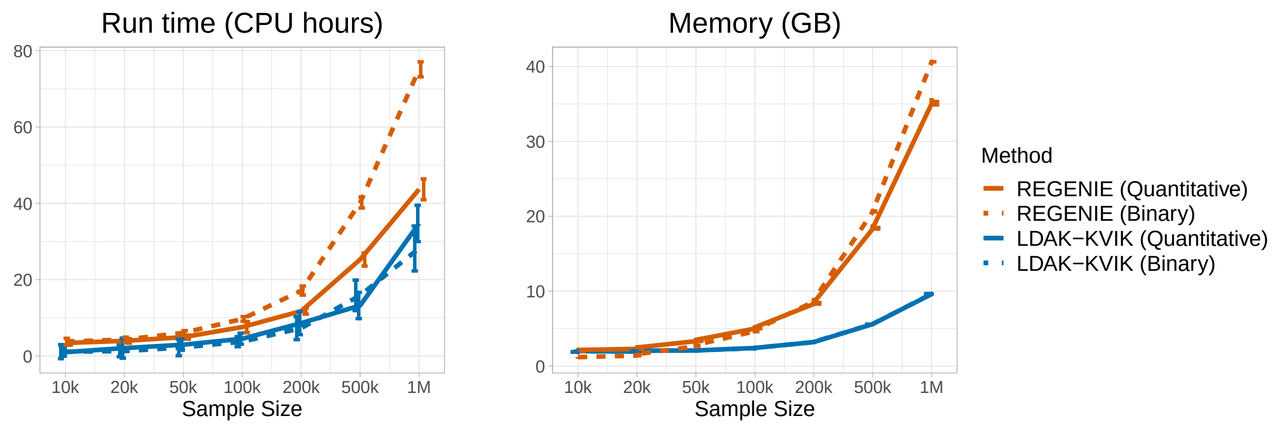

**Supplementary Figure 16: Computational requirements of LDAK-KVIK for very large datasets.** We first use the simulation feature of LDAK (`www.dougspeed.com/simulations`) to construct a dataset containing genotypes for 1 M individuals and 690 k SNPs, assuming that both individuals and SNPs are independent (i.e., no cryptic relatedness nor linkage disequilibrium). For each SNP, the MAF is randomly sampled from [0.01,0.5]. We then simulate 12 quantitative and 12 binary phenotypes (for the latter, the prevalence is assumed to be 10%), varying the heritability (0.2 or 0.5) and the number of causal SNPs (5 k or 20 k). When generating causal SNP effect sizes, we assume $\alpha = -1$. The left and right panels compare the runtimes and memory requirements, respectively, when analyzing the quantitative and binary phenotypes using REGENIE and LDAK-KVIK for sample sizes 10 k, 20 k, 50 k, 100 k, 200 k, 500 k and 1 M (the vertical segments show the variation across 12 phenotypes).

These results indicate that it is feasible to apply LDAK-KVIK to very large datasets (e.g., a million individuals). In general, we find that LDAK-KVIK is faster than REGENIE for all sample sizes, and for both quantitative and binary phenotypes. While the two tools have similar memory requirements for small samples sizes, LDAK-KVIK uses noticeably less memory than REGENIE for larger sample sizes (e.g., more than 50 k individuals). Note that the variation in runtimes across phenotypes mainly reflects the randomness in how our high-performance cluster schedules jobs, rather than differences between the genetic architectures of the phenotype being analyzed (this is evident from the variation in runtime observed for REGENIE, whose algorithmic complexity is largely independent of genetic architecture).

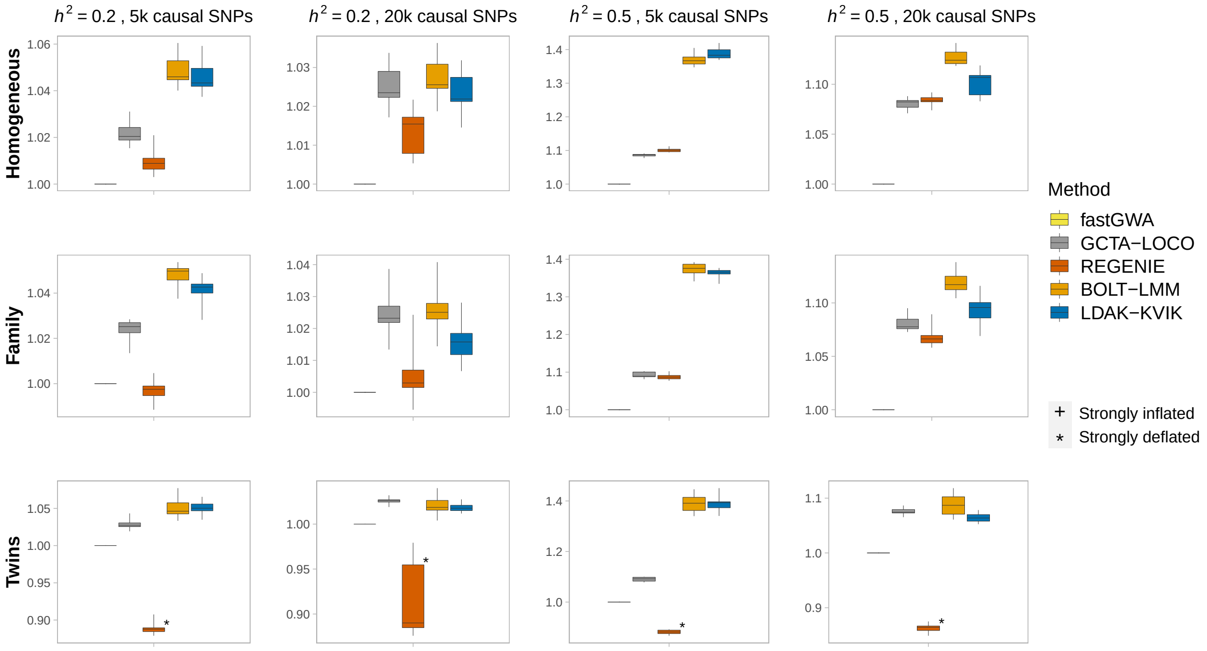

**Supplementary Figure 17: Power of MMAA tools for quantitative phenotypes.** We simulate quantitative phenotypes for the homogeneous, family, and twins datasets (each of which contains 63 k individuals). For each dataset, we consider four different scenarios, obtained by varying the heritability (0.2 or 0.5) and the number of causal SNPs (5 k or 20 k). When generating causal SNP effect sizes, we assume $\alpha = -1$. We perform single-SNP analysis using BOLT-LMM, REGENIE, GCTA-LOCO, fastGWA and LDAK-KVIK, then measure the statistical power based on the mean $\chi^2(1)$ test statistic of causal SNPs (reporting values relative to that of fastGWA for the same phenotype). In each box, the three horizontal lines mark the median and inter-quartile range across ten replicates. We use a plus sign (+) and an asterix (*), respectively, to highlight tools that show strong inflation (mean $\chi^2(1)$ of null SNPs greater than 1.02) or strong deflation (mean $\chi^2(1)$ of null SNPs less than 0.98) for a particular scenario (note that in this figure, there are no plus signs).

In general, we find that BOLT-LMM and LDAK-KVIK have the highest power across the range of scenarios considered. The power differences are largest for phenotypes with heritability 0.5 and 5 k causal SNPs (Column 3), where the mean test statistics from LDAK-KVIK and BOLT-LMM are up to 40% higher than those from fastGWA. REGENIE and GCTA-LOCO tend have intermediate power (i.e., lower than BOLT-LMM and LDAK-KVIK, but higher than fastGWA), however, we note that REGENIE has lower power than fastGWA when analysing the twins dataset, which is due to its deflated type 1 error (a consequence of the fact that REGENIE implicitly assumes $\lambda = 1$).

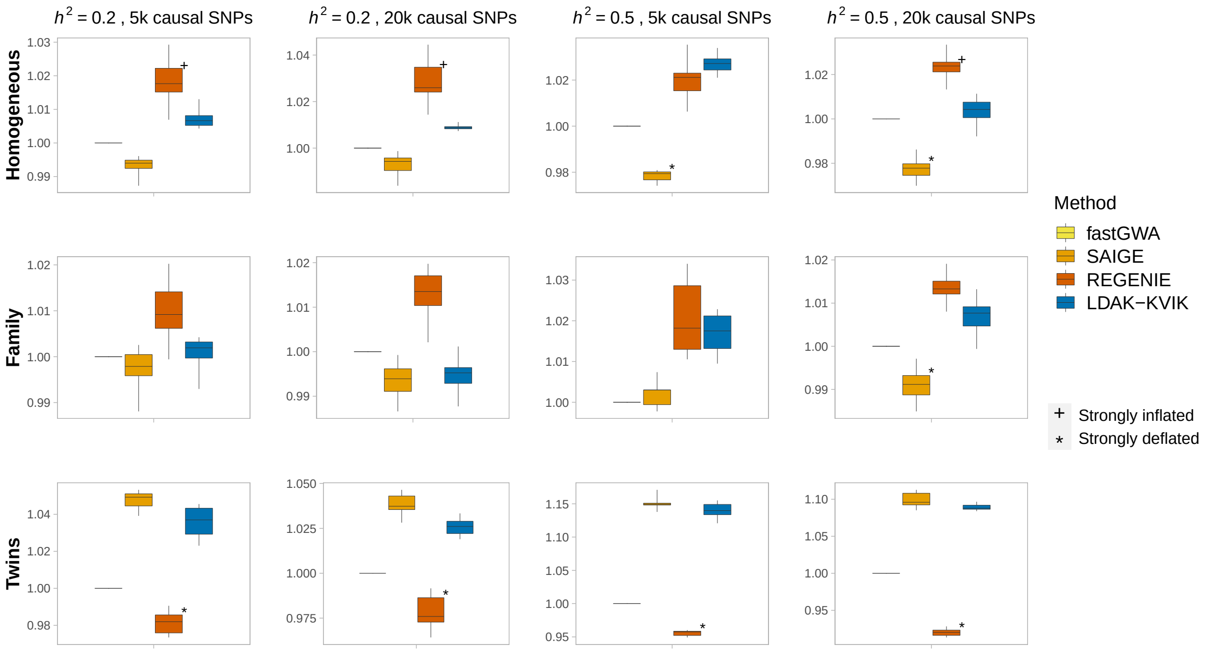

**Supplementary Figure 18: Power of MMAA tools for common binary phenotypes.** We simulate binary phenotypes with prevalence 10% for the homogeneous, family, and twins datasets (each of which contains 63 k individuals). For each dataset, we consider four different scenarios, obtained by varying the heritability (0.2 or 0.5) and the number of causal SNPs (5 k or 20 k). When generating causal SNP effect sizes, we assume $\alpha = -1$. We perform single-SNP analysis using REGENIE, GCTA-LOCO, fastGWA, SAIGE and LDAK-KVIK, then measure the statistical power based on the mean $\chi^2(1)$ test statistic of causal SNPs (reporting values relative to that of fastGWA for the same phenotype). In each box, the three horizontal lines mark the median and inter-quartile range across ten replicates. We use a plus sign (+) and an asterix (*), respectively, to highlight tools that show strong inflation (mean $\chi^2(1)$ of null SNPs greater than 1.02) or strong deflation (mean $\chi^2(1)$ of null SNPs less than 0.98) for a particular scenario.

In general, it is difficult to construct accurate PGS for binary phenotypes, and as a result, all tools tend to have very similar power. The exception is for the phenotypes with heritability 0.5 and 5 k causal SNPs (Column 3), where the genetic architecture is relatively simple, and so LDAK-KVIK tends to be more powerful than the other tools (due to it constructing more accurate PGS). Note that the apparent power advantage of REGENIE over LDAK-KVIK in the top row reflects that REGENIE tends to have strongly inflated type 1 error when analyzing common binary phenotypes for the homogeneous dataset (this is also the case for the family dataset, albeit to a lesser extent, which explains the apparent advantage for some scenarios in the middle row).

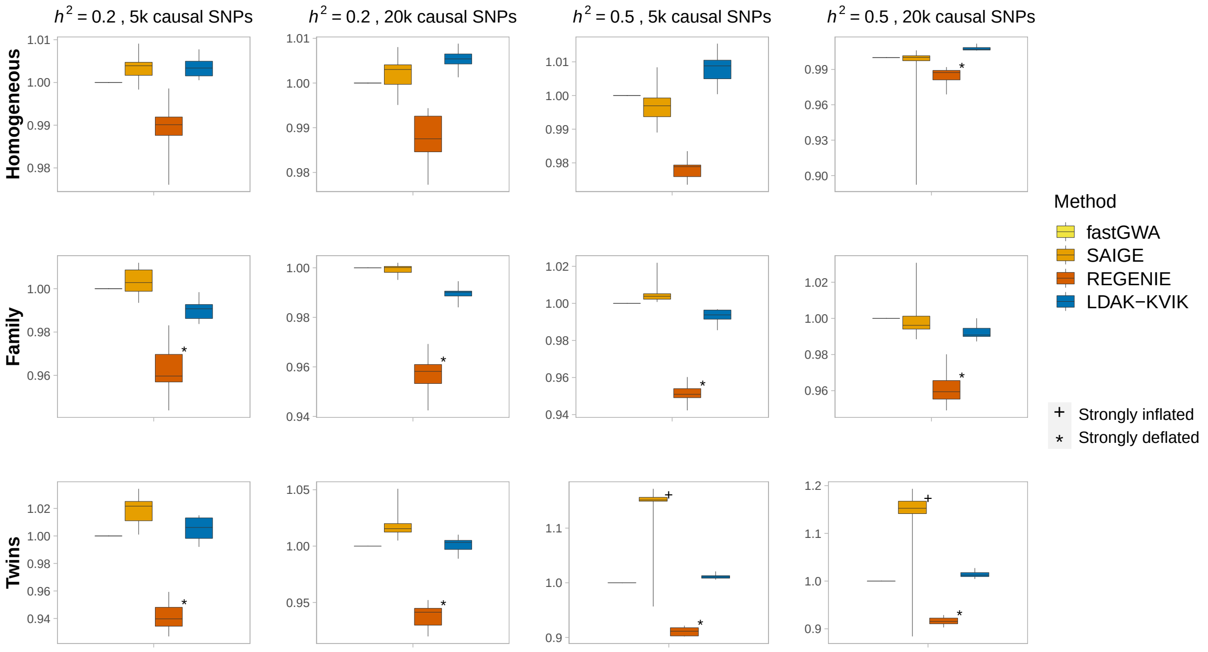

**Supplementary Figure 19: Power of MMAA tools for rare binary phenotypes.** We simulate binary phenotypes with prevalence 1% for the homogeneous, family, and twins datasets (each of which contains 63 k individuals). For each dataset, we consider four different scenarios, obtained by varying the heritability (0.2 or 0.5) and the number of causal SNPs (5 k or 20 k). When generating causal SNP effect sizes, we assume $\alpha = -1$. We perform single-SNP analysis using REGENIE, GCTA-LOCO, fastGWA, SAIGE and LDAK-KVIK, then measure the statistical power based on the mean $\chi^2(1)$ test statistic of causal SNPs (reporting values relative to that of fastGWA for the same phenotype). In each box, the three horizontal lines mark the median and inter-quartile range across ten replicates. We use a plus sign (+) and an asterix (*), respectively, to highlight tools that show strong inflation (mean $\chi^2(1)$ of null SNPs greater than 1.02) or strong deflation (mean $\chi^2(1)$ of null SNPs less than 0.98) for a particular scenario.

In general, it is difficult to construct accurate PGS for binary phenotypes, and as a result, all tools tend to have very similar power. Note that the apparent power advantage of SAIGE over LDAK-KVIK in the final two panels reflects that SAIGE tends to have strongly inflated type 1 error when analyzing rare binary phenotypes with high heritability for the twins dataset.

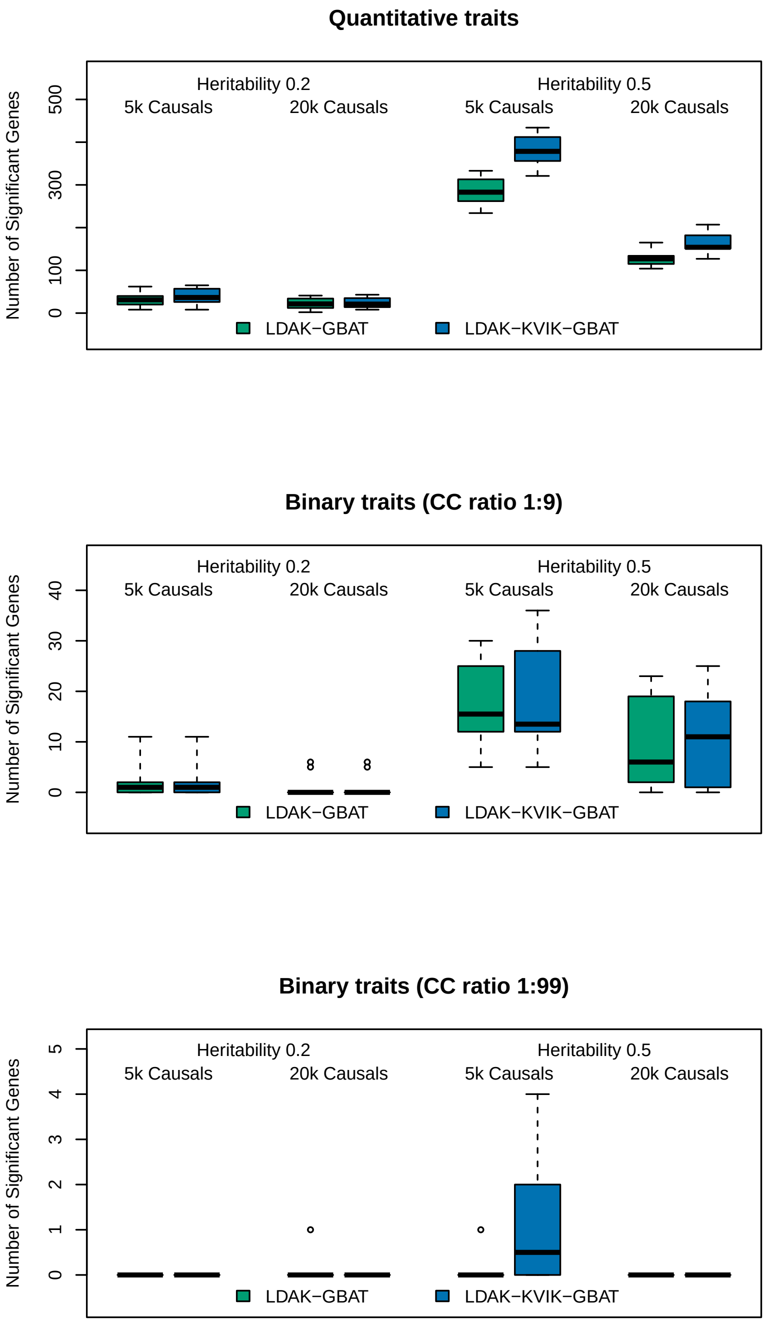

**Supplementary Figure 20: Power of gene-based association analysis for homogeneous data.** We simulate quantitative and binary phenotypes for the homogeneous dataset (63 k individuals). We consider 12 different scenarios, obtained by varying the heritability (0.2 or 0.5), the number of causal SNPs (5 k or 20 k), and for binary phenotypes, also the prevalence (10% or 1%). When generating causal SNP effect sizes, we assume $\alpha = -1$. We perform gene-based analysis using LDAK-GBAT and LDAK-KVIK-GBAT, then measure the power based on the number of causal genes with $p$-value below $0.05/17\,322 = 2.9 \times 10^{-6}$. In each box, the three horizontal lines mark the median and inter-quartile range across ten replicates.

We find that LDAK-KVIK-GBAT tends to be more powerful than LDAK-GBAT for quantitative phenotypes, whereas for binary phenotypes, the two tools have similar power.

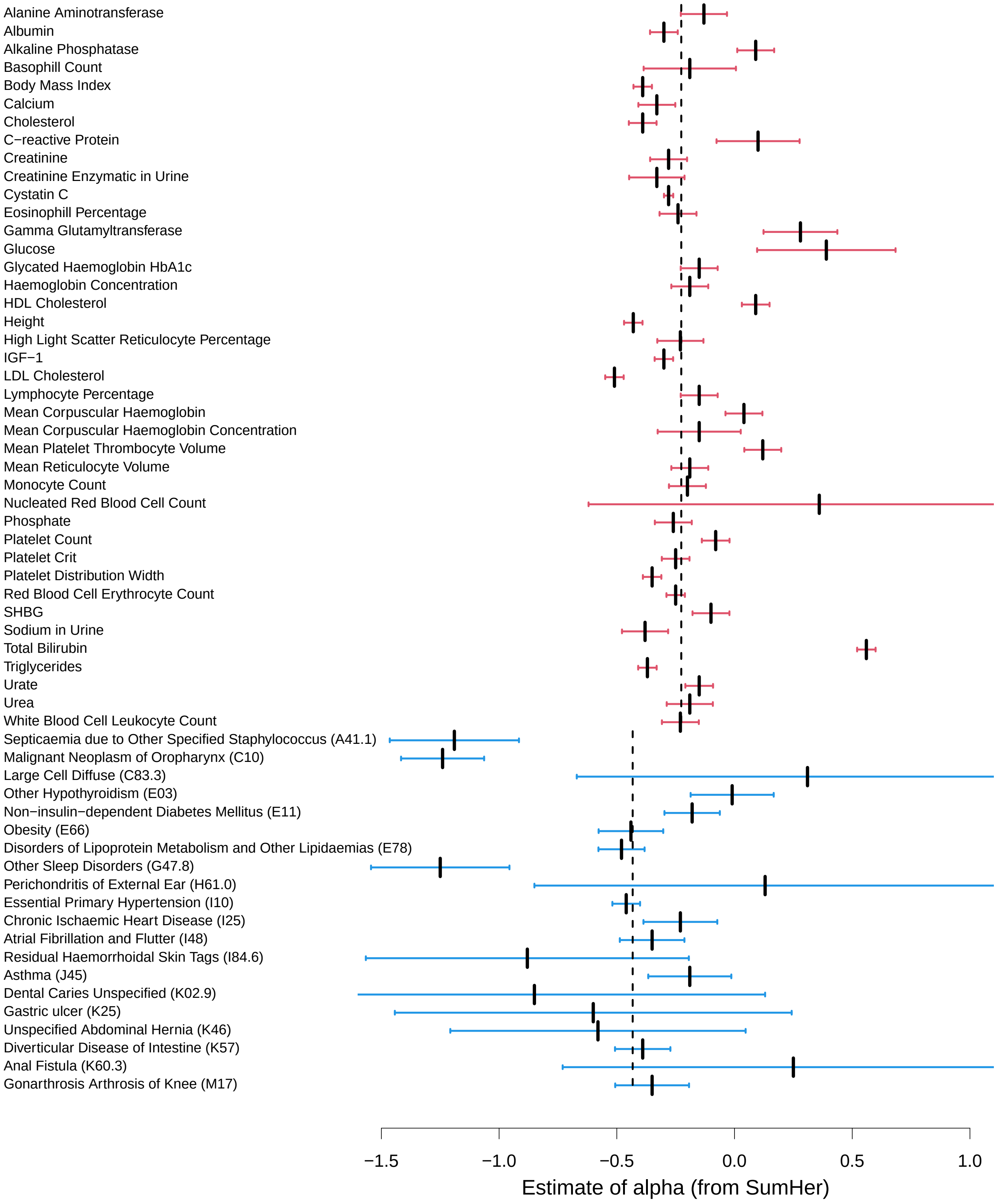

**Supplementary Figure 21: Estimates of $\alpha$ for the UK Biobank phenotypes.**

We report estimates of $\alpha$ for the 40 quantitative, then for the 20 binary phenotypes (obtained using our software SumHer[38]); the vertical dashed lines mark the inverse-variance-weighted mean estimates of $\alpha$ for the two sets of phenotypes.

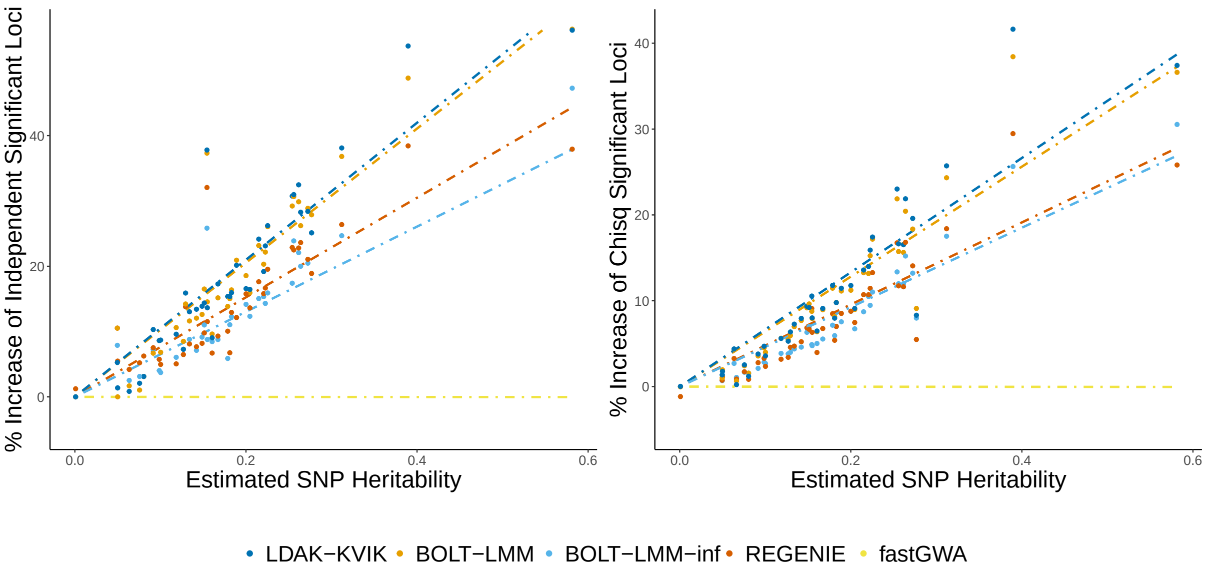

**Supplementary Figure 22: Power of MMAA tools for the 40 quantitative UK Biobank phenotypes.** We analyze each phenotype using BOLT-LMM, REGENIE, fastGWA, BOLT-LMM-inf and LDAK-KVIK, then report (left panel) the number of independent, genome-wide significant loci (SNPs with $P < 5 \times 10{-}8$, filtered so that no pair within 1 Mb has squared correlation above 0.1), and (right panel) the mean $\chi^2(1)$ test statistics of SNPs significant from fastGWA. For both plots, the values are relative to the results from fastGWA. The dashed lines are obtained by regressing the performance of each tool on the estimated SNP heritability (obtained using our software SumHer[38]).

The left panel matches the figure used in the main text, and shows that LDAK-KVIK finds most significant loci, slightly more than BOLT-LMM, and substantially more than REGENIE, fastGWA and BOLT-LMM-inf (we use BOLT-LMM-inf as a proxy for GCTA-LOCO, because it is not feasible to apply the latter to this dataset). The right panel shows that the ranking of MMAA tools is robust to changing the way we measure power.

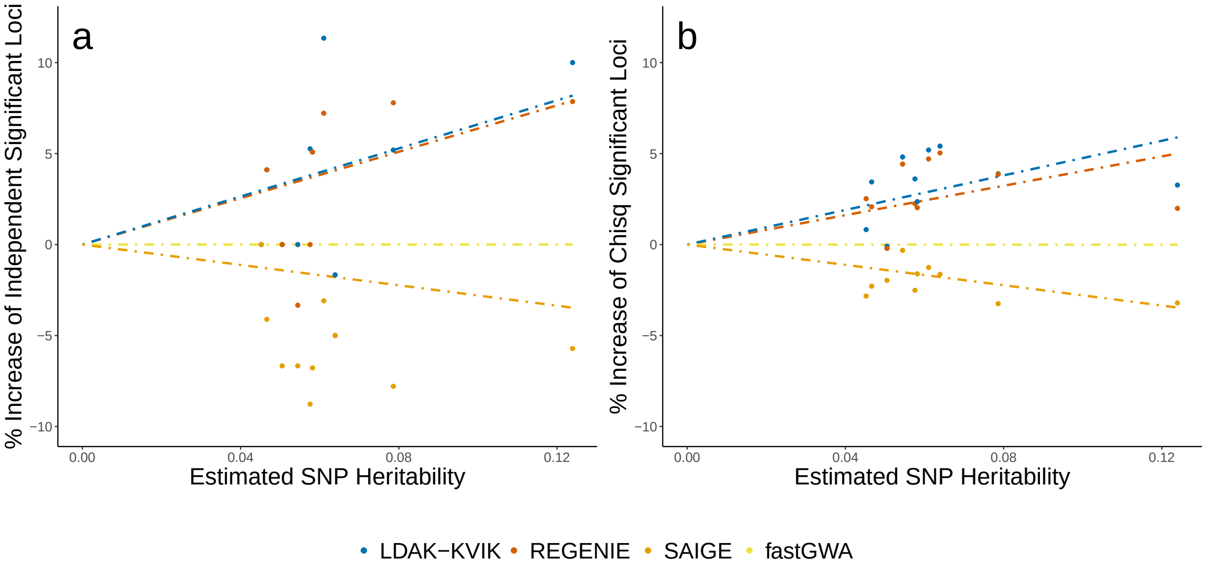

**Supplementary Figure 23: Power of MMAA tools for the 20 binary UK Biobank phenotypes.** We analyze each phenotype using REGENIE, fastGWA, SAIGE and LDAK-KVIK, then report (left panel) the number of independent, genome-wide significant loci (SNPs with $P < 5 \times 10{-}8$, filtered so that no pair within 1 Mb has squared correlation above 0.1), and (right panel) the mean $\chi^2(1)$ test statistics of SNPs significant from fastGWA. For both plots, the values are relative to the results from fastGWA. The dashed lines are obtained by regressing the performance of each tool on the estimated SNP heritability (obtained using our software SumHer[38]).

All the lines of best fit are close to the horizontal, indicating that REGENIE, SAIGE and LDAK-KVIK all have similar power to fastGWA, which reflects that their Step 1 PGS tend to have very low accuracy (Supplementary Figures 18 & 19).

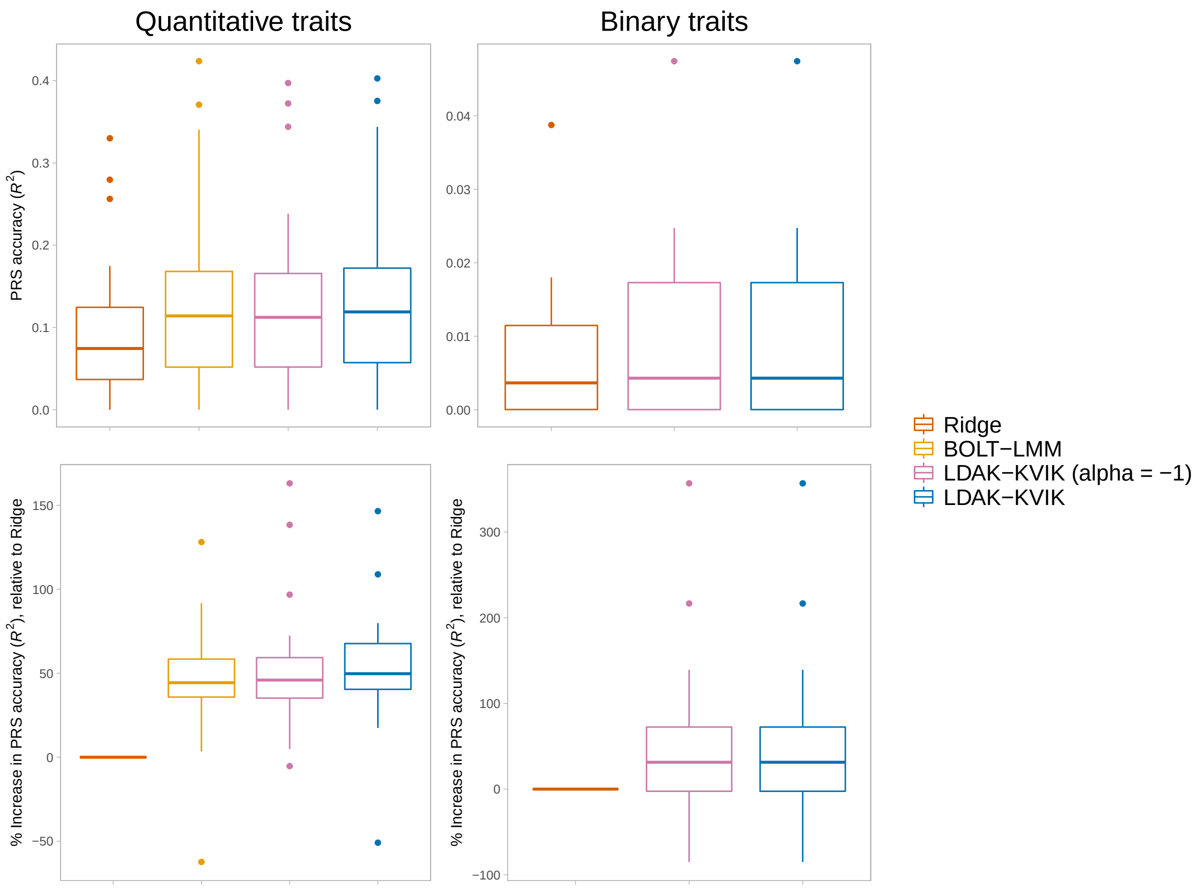

**Supplementary Figure 24: Accuracy of Step 1 PGS when analyzing the UK Biobank phenotypes.** We construct PGS for the 40 quantitative phenotypes using a ridge regression model, BOLT-LMM, a modified version of LDAK-KVIK that forces $\alpha = -1$, and the default version of LDAK-KVIK. We do the same for the 20 binary phenotypes, except that we exclude BOLT-LMM (which is only designed for quantitative phenotypes). Boxes report the accuracy of each PGS, measured by the squared correlation between predicted and observed phenotypes across 41k samples (distinct from those used to estimate the PGS effect sizes). In each box, the three horizontal lines mark the median and inter-quartile range. Note that the top panels report absolute accuracies, whereas the bottom panels report accuracies relative to those of the ridge regression PGS.

The bottom left panel demonstrates the two main reasons why LDAK-KVIK has slightly increased power relative to BOLT-LMM when analyzing the quantitative phenotypes. Firstly, LDAK-KVIK uses an elastic net prior distribution for SNP effect sizes, which results in slightly more accurate Step 1 PGS than BOLT-LMM, which assumes a mixture of normal distributions (this is evident from comparing the second and third boxes). Secondly, LDAK-KVIK estimates $\alpha$ from the data, which improves the accuracy of the Step 1 PGS relative to simply assuming $\alpha = -1$ (evident from comparing the third and fourth boxes). We include the ridge regression PGS because these are similar to those constructed by REGENIE (the latter does not report effect sizes, so we can not measure the accuracy of its PGS directly). Therefore, the panel also shows that the substantial power advantage of LDAK-KVIK (and BOLT-LMM) compared to REGENIE, is a consequence of using a mixture prior distribution for SNP effect sizes, instead of an infinitesimal distribution.

The top right panel shows that for the binary phenotypes, both REGENIE and LDAK-KVIK produce Step 1 PGS with low accuracy, explaining why their power is only slightly higher than that of classical logistic regression.

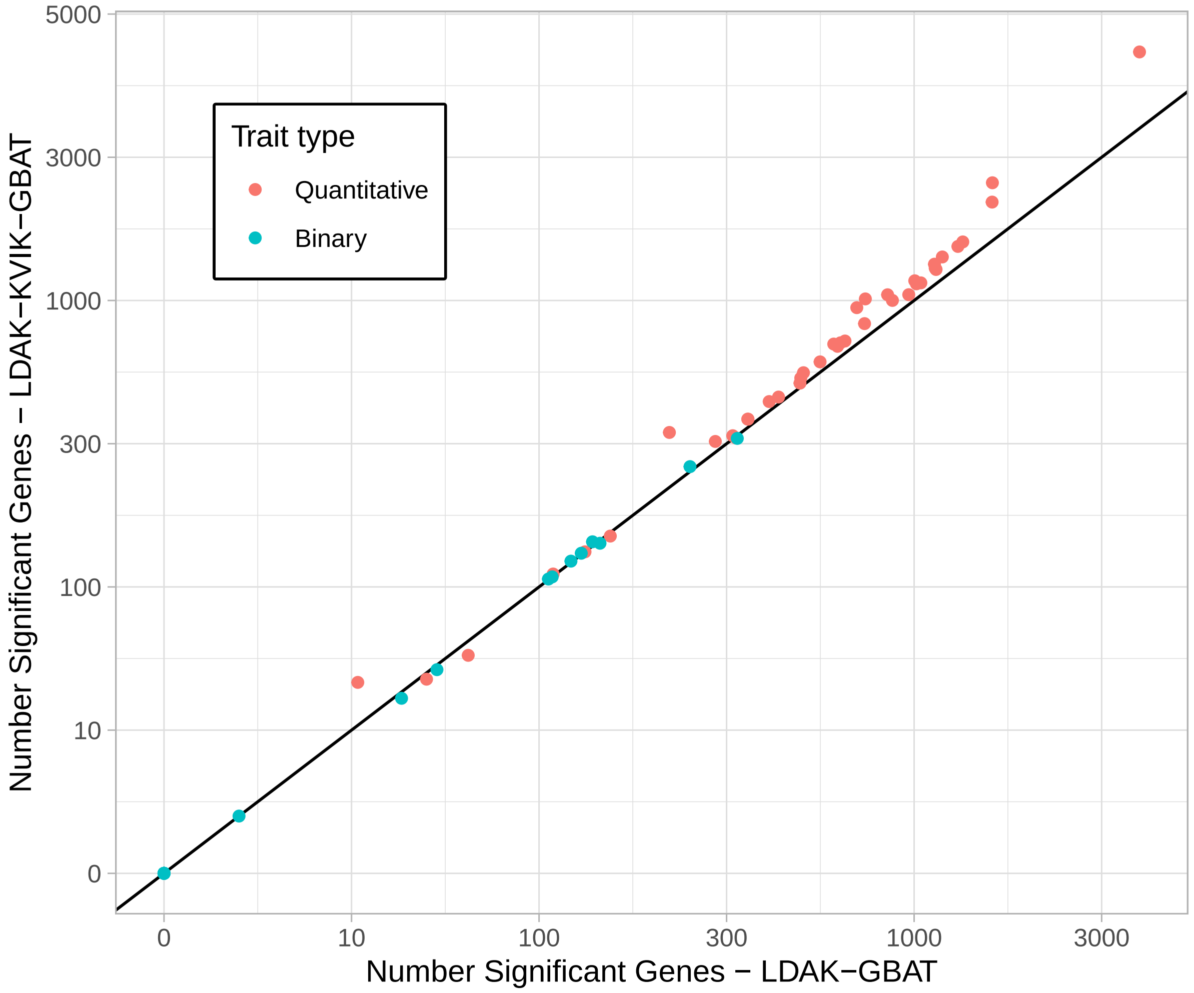

**Supplementary Figure 25: Gene-based association analysis of UK Biobank phenotypes.** Points compare the number of significant genes ($P < 0.05/17\,322 = 2.9 \times 10^{-6}$) from LDAK-GBAT and LDAK-KVIK-GBAT for each of the 40 quantitative phenotypes (red) and 20 binary phenotypes (blue). The diagonal line marks $y = x$.

When analyzing the quantitative phenotypes, LDAK-KVIK-GBAT finds on average 18% more significant genes than LDAK-GBAT, whereas for the binary phenotypes, the two tools find similar numbers of significant genes. These result mirror those for single-SNP analysis, where LDAK-KVIK was substantially more powerful than classical regression for quantitative phenotypes, but had similar power for binary phenotypes (reflecting that LDAK-KVIK produces more accurate Step 1 PGS for quantitative phenotypes than for binary phenotypes).

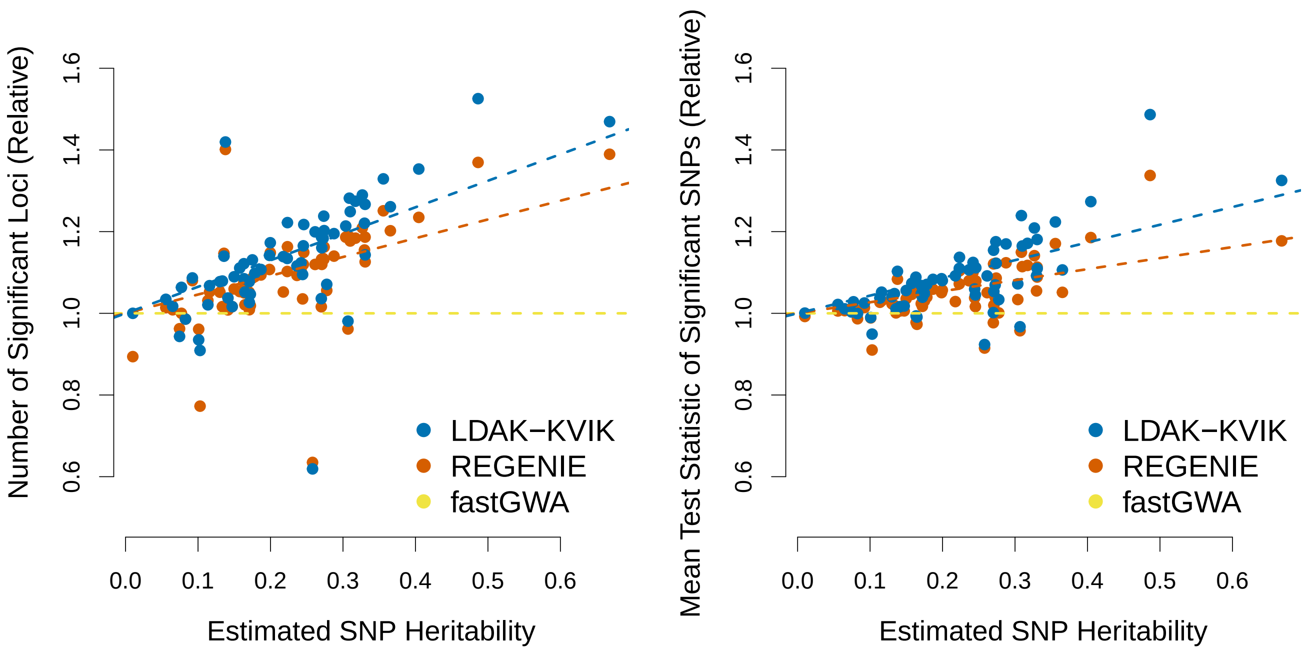

**Supplementary Figure 26: Enlarged analysis of 62 quantitative phenotypes.**

For this analysis, we increased the number of quantitative phenotypes from 40 to 62 (by adding 22 traits analyzed by the authors of fastGWA[7]), increased the total sample size from 368k to 459k (the latter represents all European UK Biobank individuals), and increased the number of SNPs from 690k to 9.0M (by including imputed variants). We analyzed each phenotype using REGENIE, fastGWA and LDAK-KVIK, then report (left panel) the number of independent, genome-wide significant loci (SNPs with $P < 5 \times 10{-}8$, filtered so that no pair within 1 Mb has squared correlation above 0.1), and (right panel) the mean $\chi^2(1)$ test statistics of SNPs significant from fastGWA. For both plots, the values are relative to the results from fastGWA. The dashed lines are obtained by regressing the performance of each tool on the estimated SNP heritability (obtained using our software SumHer[38]).

The results are consistent with those from analyzing fewer phenotypes, individuals and SNPs, in that LDAK-KVIK finds substantially more significant loci than REGENIE and fastGWA (and this remains the case if we restrict to the 22 phenotypes not considered in our previous analysis).

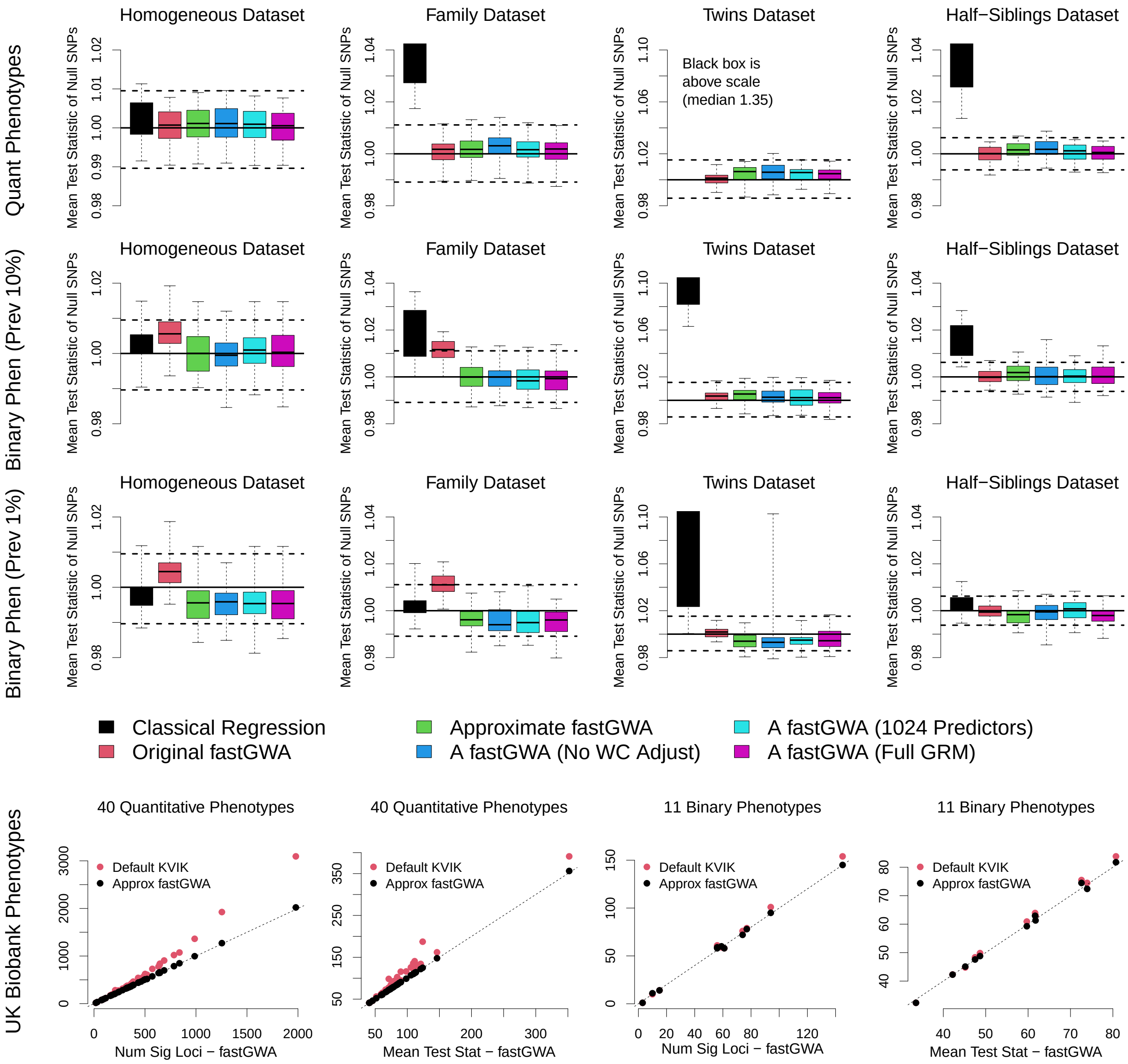

**Supplementary Figure 27: Approximate version of fastGWA.**

For the top three rows, we simulate quantitative and binary phenotypes for the homogeneous, family, twins and "half-siblings" datasets (each of which contains 63 k individuals); the half-siblings dataset is one we created just for this analysis, by generating genotypes for 21 000 families (each containing three half-siblings individuals) and 300 k SNPs in linkage equilibrium. For each dataset, we consider 12 different scenarios, obtained by varying the heritability (0.2 or 0.5), the number of causal SNPs (5 k or 20 k), and for binary phenotypes, also the prevalence (10% or 1%). When generating causal SNP effect sizes, we assume $\alpha = -1$. We perform single-SNP analysis using classical linear or logistic regression, original fastGWA, and four versions of approximate fastGWA contained within LDAK-KVIK: the default version, a version that does not adjust for winners' curse (Supplementary Figure 36), a version that increases the number of pedigree SNPs from 1024 to 2048, and a version that uses kinship estimates from the genome-wide GRM (i.e., the same as original fastGWA). We measure the type 1 error based on the mean $\chi^2(1)$ test statistic of null SNPs. In each box, the three horizontal lines mark the median and inter-quartile range across 40 phenotypes (ten replicates for each of four scenarios). The solid grey lines mark the expected value of each measure under the null hypothesis, while the dashed grey lines provide a 95% confidence interval (derived by analyzing 1000 permuted phenotypes, as explained in Supplementary Figure 3). [The description continues on the next page.]

We find that both original fastGWA and approximate fastGWA generally have good control of type 1 error for all four datasets (the exception is the small inflation when original fastGWA is applied to binary phenotypes for the family dataset). Note that it is reassuring that both original fastGWA and approximate fastGWA perform well for the half-siblings dataset, as we designed this to represent a worst-case scenario (i.e., a dataset that contains multiple, moderate relationships). For approximate fastGWA, we find that the correction for winners' curse has limited impact (dark blue boxes), and that the results are very similar if we double the number of kinship SNPs (light blue boxes). Further, we find that the default version (green boxes), which estimates kinships based on a GRM constructed using only 1024 SNPs, performs as well as the alternative version that uses kinship estimates from a GRM based on 690 k or 300 k SNPs (purple boxes). Note that this suggests that the inflation observed for original fastGWA when analyzing binary traits for the family dataset is because original fastGWA updates fixed effects (whereas approximate fastGWA fixes their values under the null).

For the bottom row, we apply both original and approximate fastGWA to the 40 quantitative UK Biobank phenotypes, and to the 11 binary UK Biobank phenotypes with at least one genome-wide significant locus (the nine excluded binary phenotypes tend to be those with lowest prevalence and/or heritability). The black points show that the two tools have very similar performance, both when measured by the number of independent, genome-wide significant loci (SNPs with $P < 5 \times 10{-}8$, filtered so that no pair within 1 Mb has squared correlation above 0.1), and when measured by the mean $\chi^2(1)$ test statistics of SNPs significant from linear regression (for comparison, the red points show the performance of LDAK-KVIK).

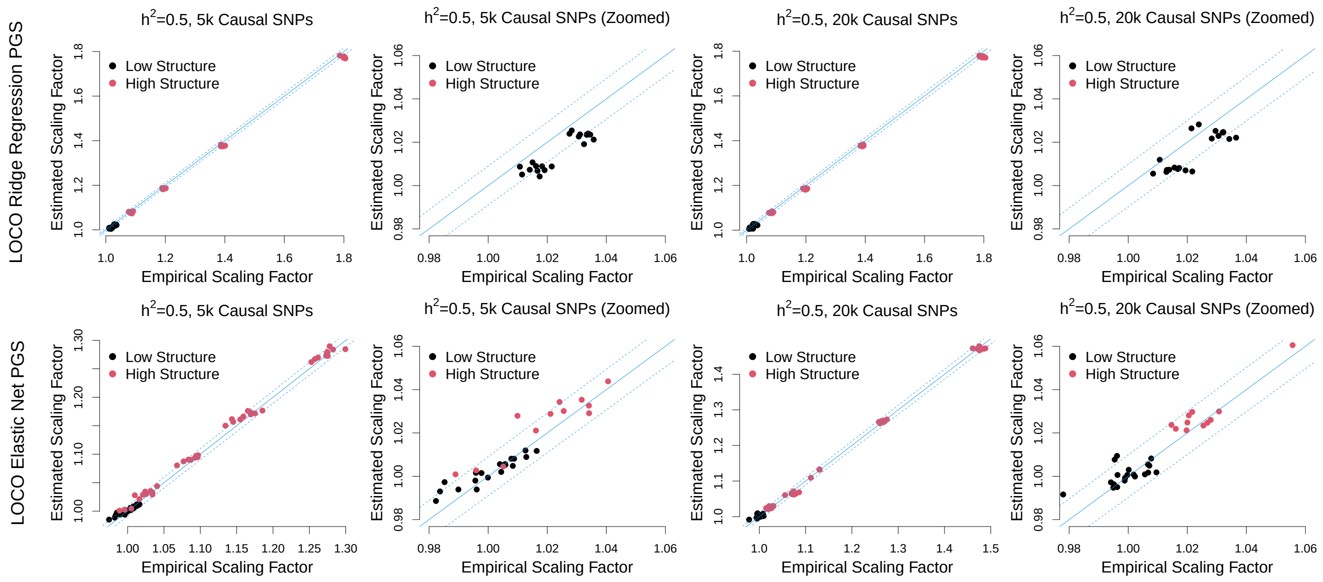

**Supplementary Figure 28: LDAK-KVIK accurately estimates $\lambda$**

We construct datasets with varying levels of relatedness; each dataset has 100 000 SNPs in linkage equilibrium, and 63 000 individuals partitioned into pairs of individuals with relatedness varying from 0 to 1 (e.g., when the relatedness is 0.5, the dataset resembles one containing 31 500 pairs of full-siblings). We then simulate phenotypes with heritability 0.5 and either 5 k or 20 k causal variants, whose effect sizes are generated assuming $\alpha = -1$. For the top row, we run LDAK-KVIK using LOCO ridge regression PGS in Step 1, and compare empirical estimates of the test statistic scaling factor with estimates of $\lambda'$ from the Grammar-Gamma formula [19] (the empirical estimates are obtained by computing the reciprocal of the mean test statistic across null SNPs). For the bottom row, we run LDAK-KVIK using LOCO elastic net PGS in Step 1, and compare empirical estimates of the test statistic scaling factor with estimates of $\lambda$ from LDAK-KVIK. Note that Columns 2 & 4 are zoomed-in versions of Columns 1 & 3, respectively, while for the bottom row, we force LDAK-KVIK to always estimate $\lambda$ (whereas it usually sets $\lambda = 1$ for datasets it determines have low structure). The solid diagonal lines mark $y = x$, while the dashed diagonal lines provide 95% confidence intervals for the empirical estimates of $\lambda$ (the uncertainty reflects that the empirical estimates are computed using a finite number of null SNPs).

The top row confirms that the Grammar-Gamma formula accurately estimates $\lambda'$, while the bottom row shows that our method for estimating $\lambda$ is generally accurate, albeit less precise than the Grammar-Gamma formula. The latter reflects that our method computes estimates of the form $\lambda = \lambda' r$, where $r$ is the ratio of test statistics across weakly-associated SNPs (i.e., the estimates include both noise from applying the Grammar-Gamma formula, and noise from estimating the ratio of test statistics). Note that this figure indicates that it would be valid for LDAK-KVIK to always estimate $\lambda$, regardless of how structured the dataset being analyzed. However, it also shows that for datasets which LDAK-KVIK determines to have low structure (the black points in the above figure), the empirical estimate of $\lambda$ is not significantly different to one, and thus for these datasets it suffices (and is more efficient) to simply set $\lambda = 1$.

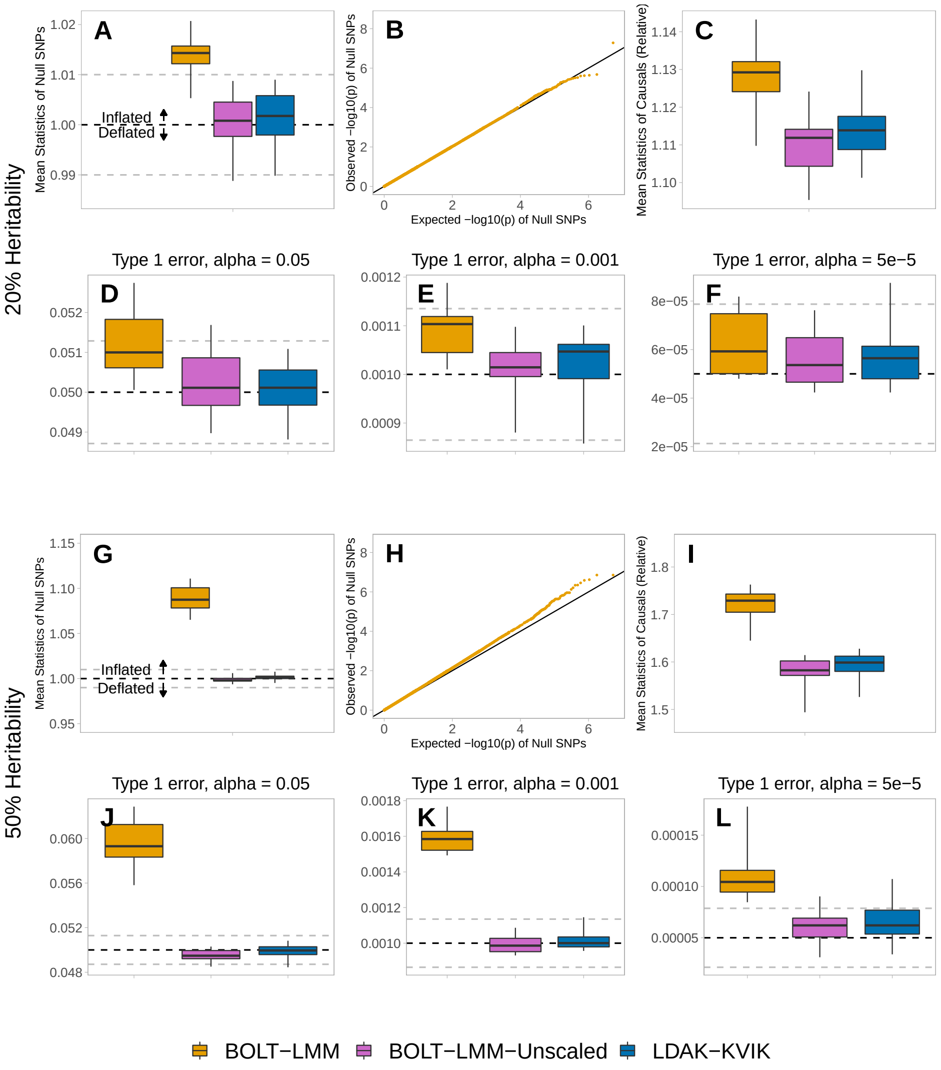

**Supplementary Figure 29: BOLT-LMM can produce inflated test statistics.** We generate quantitative phenotypes for the white dataset (368 k individuals) with heritability 0.2 or 0.5 and 5 k causal SNPs. When generating causal SNP effect sizes, we assume $\alpha = -0.25$ (i.e., per-SNP heritability increases with MAF). We analyze each phenotype using BOLT-LMM, BOLT-LMM-Unscaled (a modified version of BOLT-LMM, where we force $\lambda = 1$) and LDAK-KVIK. For each tool, we measure the type 1 error based on the mean $\chi^2(1)$ test statistic of null SNPs (Panels A & G), and based on the proportions of null SNPs with $p$-values below 0.05, 0.001 and $5 \times 10^5$ (Panels D & J, Panels E & K and Panels F & L, respectively), while we measure the power based on the mean $\chi^2(1)$ test statistic of causal SNPs, relative to the results from classical linear regression (Panels C & I). In each box, the three horizontal lines mark the median and inter-quartile range across ten replicates, while for the type 1 error panels, the solid grey lines mark the expected value of each measure under the null hypothesis, and the dashed grey lines provide a 95% confidence interval (derived by analyzing 1000 permuted phenotypes, as explained in Supplementary Figure 3). Finally, Panels B & H provide QQ-plots for BOLT-LMM, constructed using only null SNPs and combined across all ten replicates.

We see that BOLT-LMM tends to produce inflated test statistics, and that this inflation can be substantial (e.g., Panel G shows that across the phenotypes with heritability 0.5, the average inflation is 9%). The inflation reflects that BOLT-LMM tends to overestimate $\lambda$ (evidenced by the fact that the inflation disappears when we force $\lambda = 1$), and is mainly a result of BOLT-LMM (implicitly) assuming $\alpha = -1$ when estimating $\lambda$.

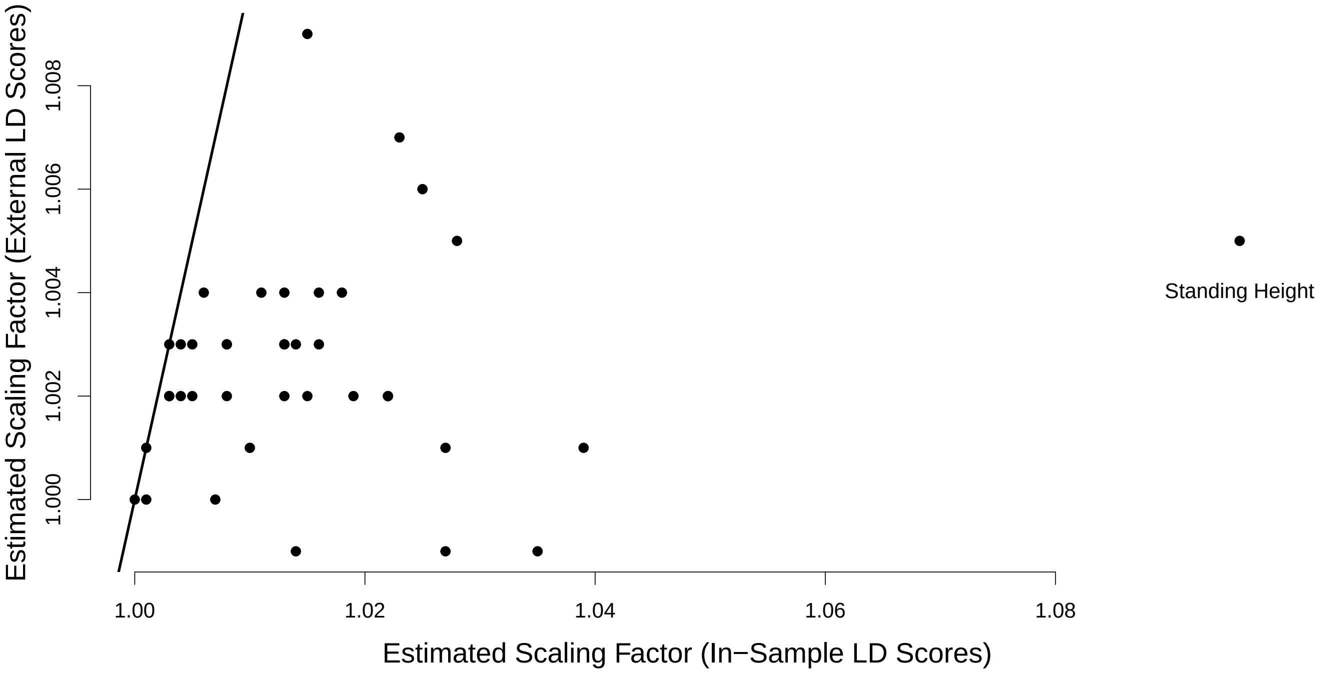

**Supplementary Figure 30: Sensitivity of BOLT-LMM to choice of LD Scores.**
BOLT-LMM estimates the test statistic scaling factor $\lambda$ using LDSC (specifically, it sets $\lambda$ so that the scaled test statistics have the same estimated intercept as those obtained using LOCO ridge regression PGS). This requires the user to provide LD Scores, which ideally should be proportional to the expected heritability tagged by each SNP. However, to compute the LD Scores, it is necessary to make assumptions regarding which SNPs are expected to contribute heritability (strictly, it is necessary to specify a heritability model, which describes the relative expected heritability contributed by each SNP). The most commonly-used LD Scores are those provided by the authors of LDSC[36], which were computed using 1000 Genome Project[35] data for 379 individuals and approximately 10M common SNPs. Using these LD Scores corresponds to assuming that the 10 M SNPs in the 1000 Genome Project data are each expected to contribute one unit of heritablity (while all other SNPs are assumed to not contribute heritability). By contrast, we can instead use in-sample LD Scores, which corresponds to assuming each SNP in the dataset is expected to contribute one unit of heritability.

This figure shows the impact on estimates of $\lambda$ of switching from using external LD Scores (those computed using the 1000 Genome Project data) to in-sample LD Scores (i.e., using the 690 k Step 1 SNPs) when analyzing the 40 quantitative UK Biobank phenotypes. In general, the estimates of $\lambda$ are larger when using in-sample LD Scores, with the most extreme case occurring for height, where the in-sample LD Scores estimate is 1.10, while the external LD scores estimate is 1.01 (this reflects that height is the most heritable trait, and likely one of the most polygenic).

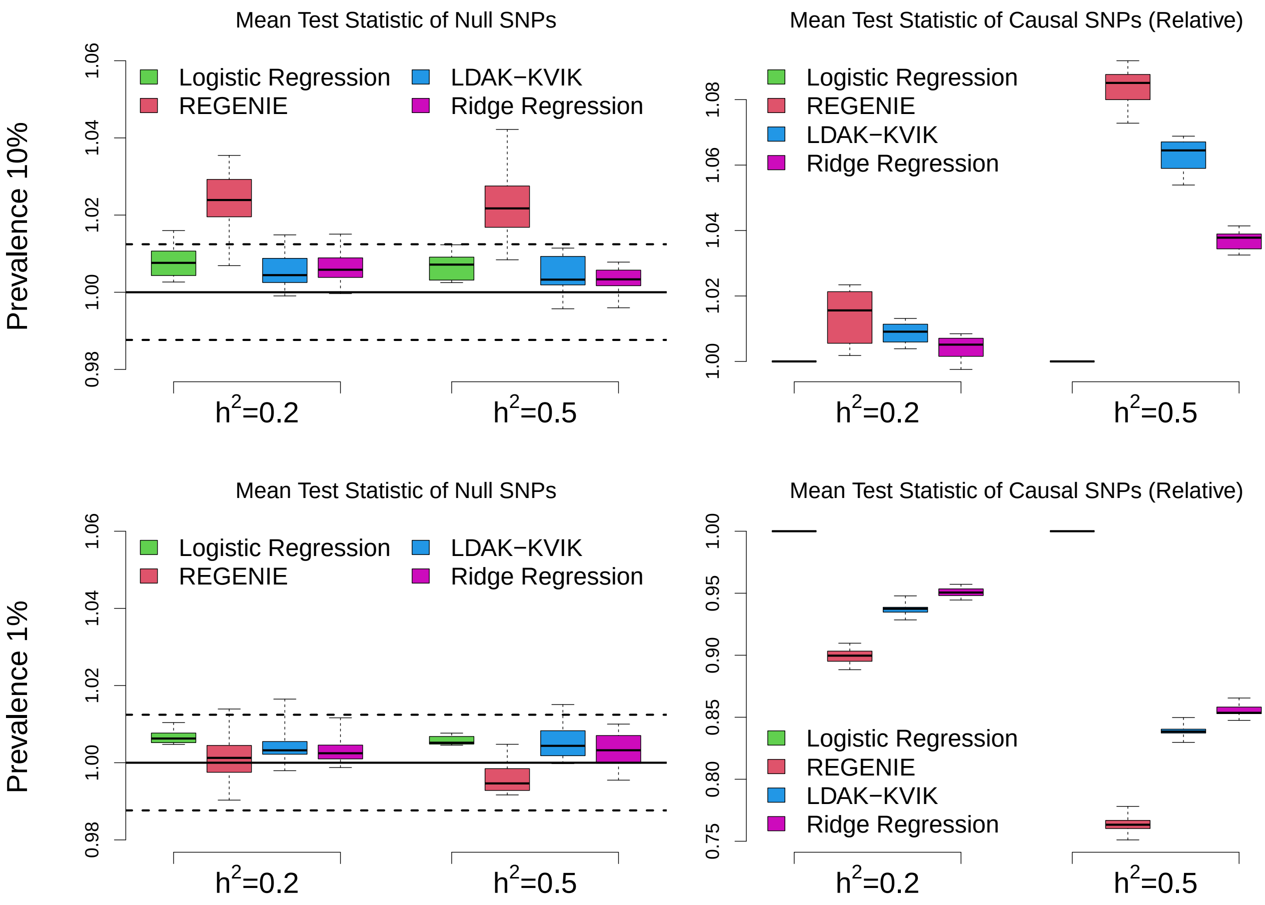

**Supplementary Figure 31: Analyzing ascertained data.**
We simulate ascertained datasets as follows. We first construct a dataset containing genotypes for 5.1 M individuals and 10 k SNPs, assuming that both individuals and SNPs are independent (i.e., no cryptic relatedness nor linkage disequilibrium). We then use this dataset to simulate binary phenotypes with heritability 0.2 or 0.5, 10 k causal SNPs, prevalence 10% or 1%, and effect sizes generated assuming $\alpha = -0.25$. For each phenotype, we sample 50 k cases and 50 k controls, and add their genotypes to 50 k randomly-sampled genotypes. The result is multiple datasets, each containing 100 k individuals and 60 k SNPs, where half the individuals are cases and half controls. We analyze these using classical logistic regression, REGENIE, the default version of LDAK-KVIK and a version of LDAK-KVIK that uses ridge regression PGS, then report the mean $\chi^2(1)$ test statistics across null and causal SNPs (left and right columns, respectively).

We find that logistic regression and LDAK-KVIK have well-controlled type 1 error, but that REGENIE produces inflated test statistics when applied to common binary phenotypes (mirroring the results for non-ascertained binary phenotypes in Supplementary Figure 7). For this reason, we include the ridge regression version of LDAK-KVIK, as this gives an idea how the results from REGENIE would look if the latter was well-calibrated. We find that there is the MMAA tools are most powerful for common binary phenotypes, but that classical logistic regression ist most powerful for rare binary phenotypes. We believe this may be because the ascertainment causes correlations between the causal SNPs (i.e., long-range linkage disequilibrium), and therefore the LOCO PGS used by the MMAA tools remove causal variation from the target chromosome. Overall, these analyses caution against using MMAA tools when analyzing acertained rare binary phenotypes.

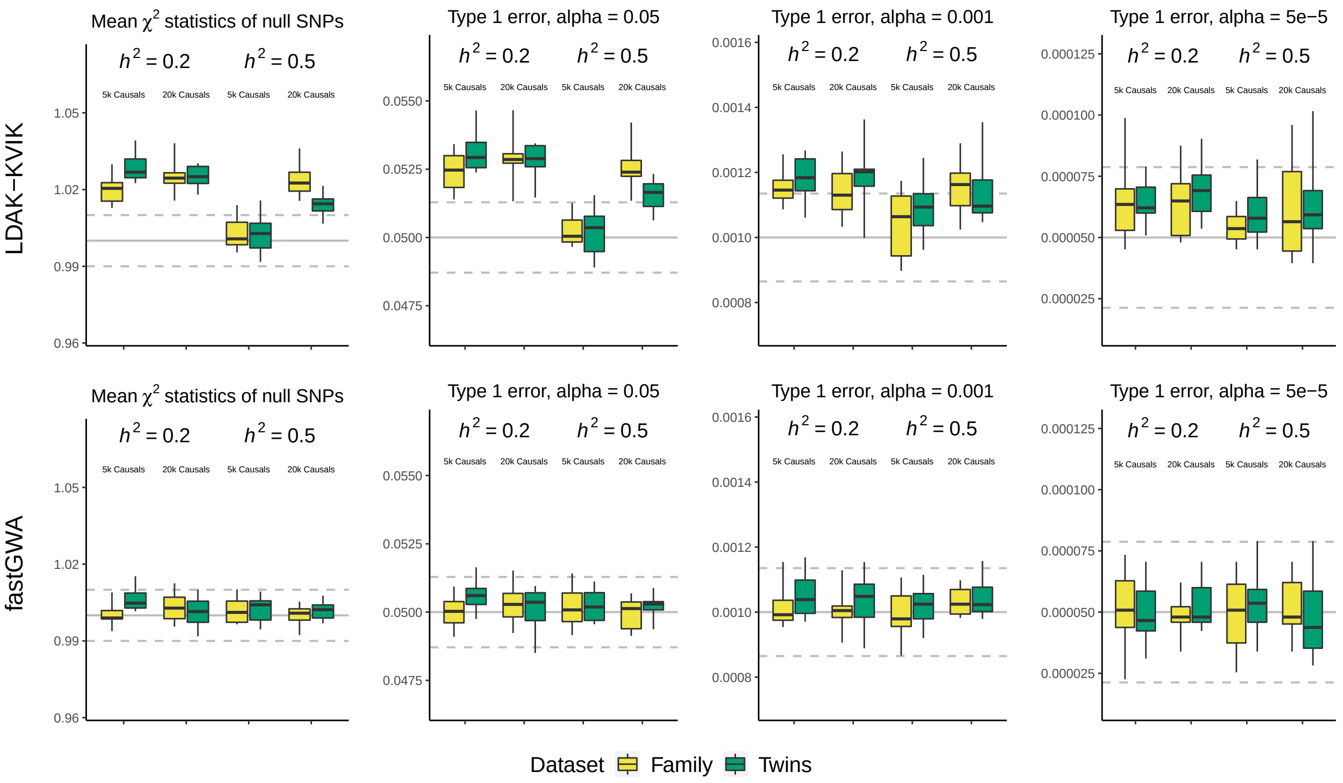

**Supplementary Figure 32: Phenotypes with a strong environmental component.** We simulate quantitative phenotypes for the family and twins dataset where common environment explains 10% of phenotypic variance. For each dataset, we consider four different scenarios, obtained by varying the heritability (0.2 or 0.5) and the number of causal SNPs (5 k or 20 k). We perform single-SNP analysis using fastGWA and LDAK-KVIK, then measure the type 1 error based on the mean $\chi^2(1)$ test statistic of null SNPs (Column 1), and based on the proportions of null SNPs with $p$-values below 0.05, 0.001 and $5 \times 10^5$ (Columns 2, 3 & 4, respectively). In each box, the three horizontal lines mark the median and inter-quartile range across ten replicates. The solid grey lines mark the expected value of each measure under the null hypothesis, while the dashed grey lines provide a 95% confidence interval (derived by analyzing 1000 permuted phenotypes, as explained in Supplementary Figure 3).

Consistent with the results of Jiang *et al.*[7], we find that LDAK-KVIK tends to produce inflated test statistics, whereas fastGWA tends to produce well-calibrated test statistics. We believe the reason for this is as follows. For phenotypes with a strong environmental component, the observed phenotypic variance matrix takes the form $V_O = \hat{h}^2_G G + \hat{h}^2_P P$, where the $(n \times n)$ matrices $G$ and $P$ record, respectively, the genetic similarity and the degree of shared environment between pairs of individuals. By default, LDAK-KVIK uses a SNP-based PGS, and therefore assumes the phenotypic variance matrix takes the form $V = K\hat{h}^2 + I(1 - \hat{h}^2)$, where $K$ is a (non-sparse) GRM. By contrast, fastGWA uses a pedigree-based PGS, and so its assumed variance matrix takes the form $\hat{V}_S = K_S\hat{h}^2 + I(1 - \hat{h}^2)$, where $K_S$ is a sparse GRM. We believe that the inflation observed for LDAK-KVIK reflects that $V$ is a poor approximation of $V_O$ (specifically, we think it tends to underestimate the phenotypic similarity for closely-related pairs of individuals). By contrast, the fact that fastGWA produces well-calibrated test statistics reflects that $V_S$ is a better approximation for $V_O$ (suggesting that for phenotypes with a strong environmental component, it is more important to accurately model the phenotypic similarity of closely-related pairs of individuals, even if this means poorly modeling the similarity of distantly-related pairs).

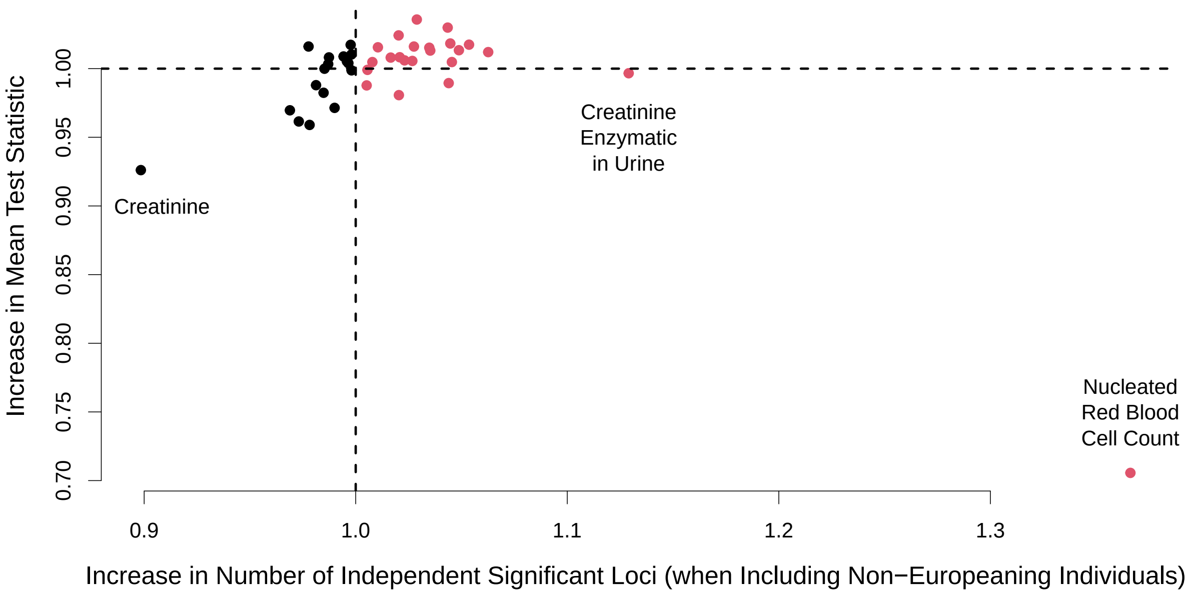

**Supplementary Figure 33: Including non-European individuals in the analysis.**

The results of the analyses of simulated phenotypes reported in Supplementary Figures 12, 13 & 14 indicate that it is valid to apply LDAK-KVIK to datasets containing a mixture of ancestries. This motivated us to repeat the analysis of the 40 quantitative UK Biobank phenotypes without filtering based on ancestry. The above figure reports the increase in the number of independent, genome-wide significant loci (SNPs with $P < 5 \times 10{-}8$, filtered so that no pair within 1 Mb has squared correlation above 0.1), and the increase in the mean test statistic of significant SNPs, when we switch from analyzing only European individuals (average sample size 438 k), to analyzing all individuals (average sample size 461 k). Note that the numbers of individuals are substantially higher than for the previous analysis (average sample size 349 k), because then we restricted to white British individuals (a subset of the Europeans) and held back 10% of individuals so that we could test the accuracy of the Step 1 PGS.

In general, we find that the two sets of analyses give very similar results; for example, for 36 of the 40 phenotypes, the number of independent, significant loci changes by less than 5%, while the number increases for 23 phenotypes and reduces for 17 phenotypes (marked by red and black points, respectively).

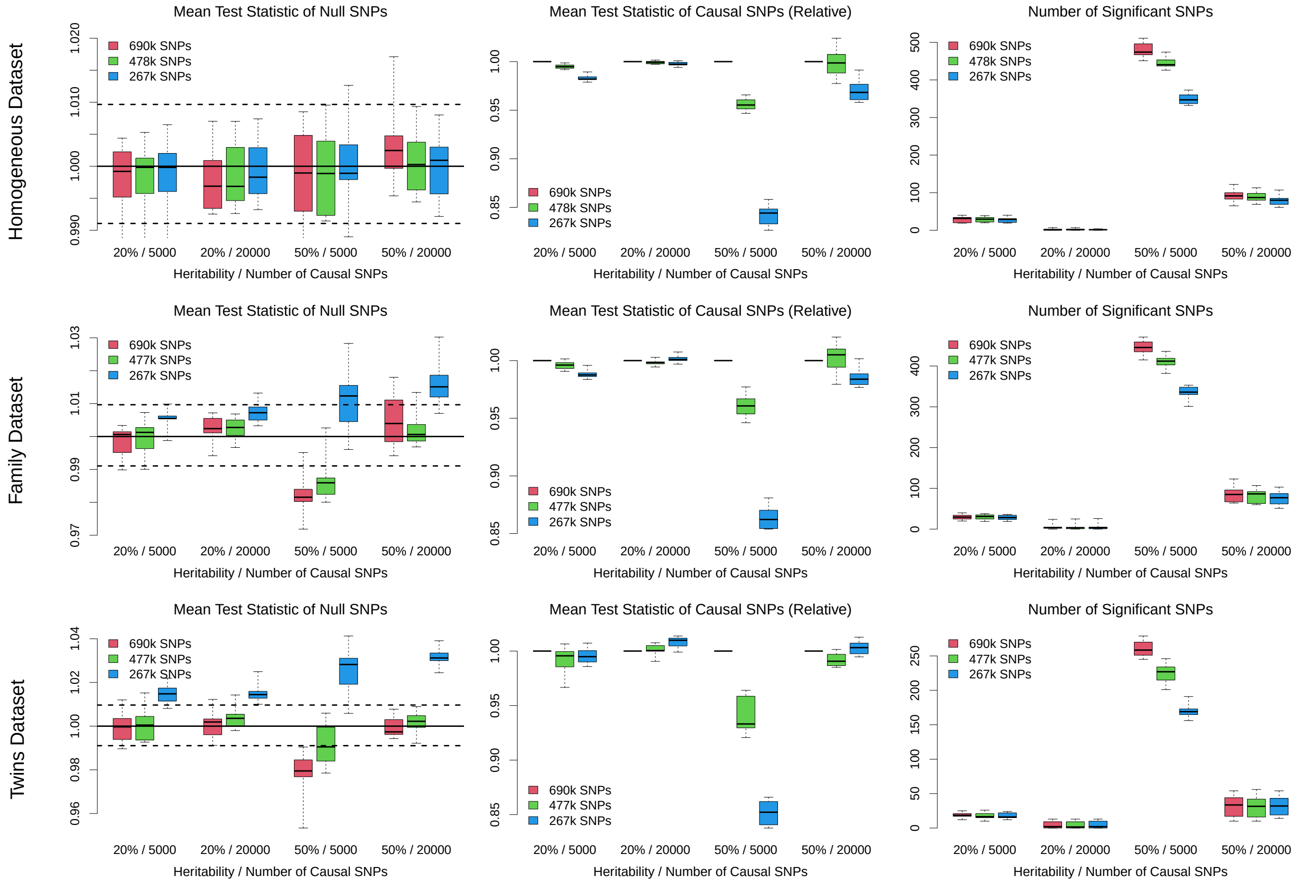

**Supplementary Figure 34: Reducing the number of Step 1 SNPs.**

Most of our applications of LDAK-KVIK used 690 k SNPs when constructing the Step 1 PGS. However, we recognize that LDAK-KVIK would run substantially faster if we reduced this number. Here, we reanalyze the simulated quantitative phenotypes for the homogeneous, family and twins datasets (which have heritability either 0.2 or 0.5, have either 5 k or 20 k causal SNPs, and were generated assuming $\alpha = -1$). We examine the impact on the type 1 error and power of LDAK-KVIK (measured based on the mean test statistics of null and causal SNPs, or the number of causal SNPs with $P < 5 \times 10{-}8$), if we reduce the number of Step 1 SNPs by approximately one-third or two-thirds (which we do by pruning the SNPs in each dataset so that no pair remains within 1 Mb with squared-correlation above 0.5 or 0.2, respectively).

In general, we find that reducing the number of Step 1 SNPs has limited impact on the type 1 error of LDAK-KVIK (the exception is when analyzing phenotypes with heritability 0.5 for the twins dataset, where LDAK-KVIK shows evidence of inflation). By contrast, we find that reducing the number of Step 1 SNPs tends to reduce the power of LDAK-KVIK, reflecting that it results in less accurate Step 1 PGS. Based on this analysis, we recommend that LDAK-KVIK is run with between 500 k and 700 k Step 1 SNPs. This can normally be achieved by restricting to high-quality directly-genotyped SNPs. However, if this is not possible, we instead suggest thinning SNPs so that no pair remains within 100 kb with squared-correlation above 0.5.

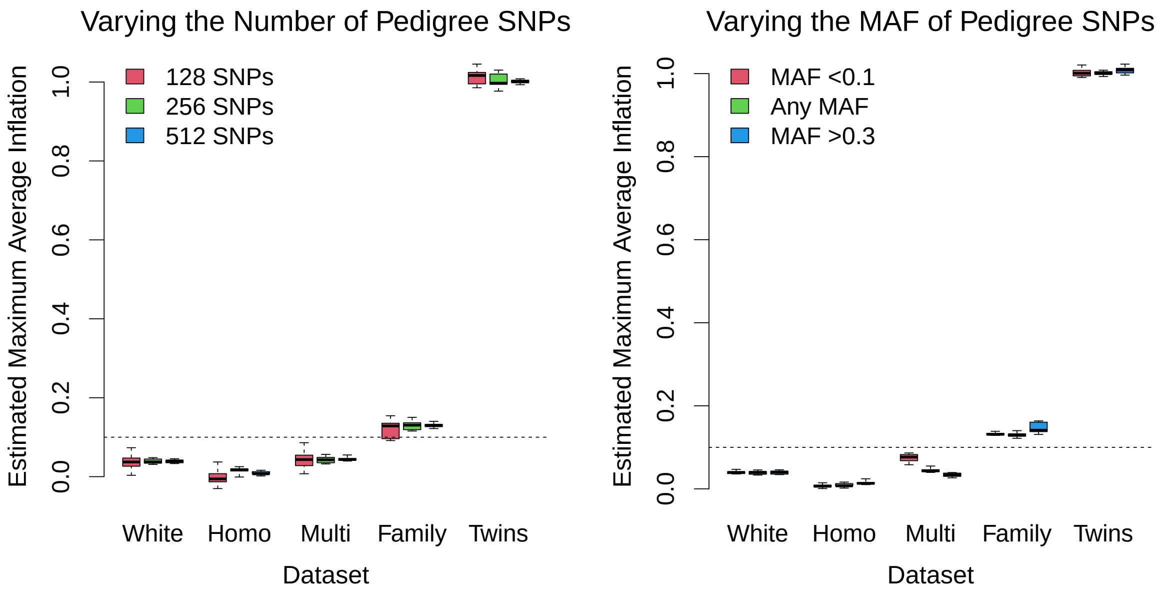

**Supplementary Figure 35: Robustness of our test for structure**

By default, our test for structure picks 512 SNPs randomly from across the genome, then uses these to compute $n\hat{\rho}^2$, an estimate of the maximum average inflation of test statistics; it then determines a dataset has high structure if $n\hat{\rho}^2 > 0.1$ and the corresponding $p$-value is below 0.001. This figure reports estimates of the maximum average inflation for the white, homogeneous, family, twins and multi-ancestry datasets. In each box, the three horizontal lines mark the median and inter-quartile range across ten replicates, while the horizontal line at $y = 0.1$ shows the threshold for determining whether a dataset has high structure. The first panel shows that our test for structure is robust to changing the number of SNPs (i.e., reducing from 512 to either 128 or 256), while the second panel shows it is robust to the MAF of the SNPs used (i.e., if we restrict to SNPs with either MAF<0.1 or MAF>0.3).

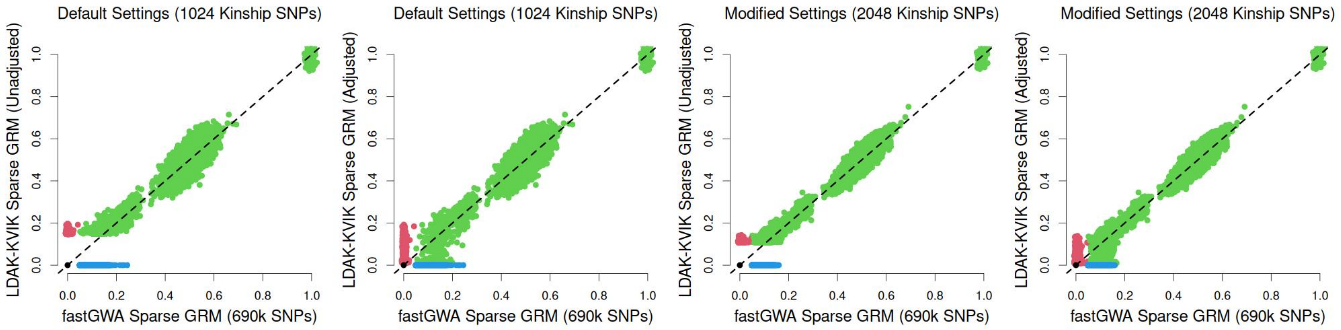

**Supplementary Figure 36: Adjusting estimates of relatedness for winners' curse.**
fastGWA constructs a sparse GRM $K_S$ by computing the genome-wide GRM $K = XX^T/m$, then setting to zero elements of $K$ below 0.05. Our approximate version of fastGWA instead constructs the sparse GRM $K'_S$ by computing $K' = X_S X_S^T/1024$, where $X_S$ contains the genotypes of 1024 SNPs picked semi-randomly from across the genome (specifically, we first randomly pick 5,120 SNPs, then retain the 1024 SNPs with highest variance), then setting to zero off-diagonal elements that are not significantly above the mean ($P > 0.1/n$). To determine significance, we assume the null distribution $N(a, b)$, where $a$ and $b$ are the mean and variance of the off-diagonal values of $K'$, respectively. Note that the standardization of genotypes ensures that $K'$ has sum zero and trace $n$, and therefore $a = -1/(n-1)$, while it can be shown[4] that $b \approx 1/1024$ (we obtain $b$ exactly by computing $X_S^T X_S$ and the diagonal values of $K'$).

Let $T$ denote the threshold for truncating $K'$ (e.g., if $n = 63\,k$, then $T = a + 4.7\sqrt{b} \approx 0.15$, while if $n = 368\,k$, then $T = a + 5.0\sqrt{b} \approx 0.16$), and suppose individuals $i$ and $i'$ are significantly related (i.e., $K'_{i,i'} > T$). In general, $K'_{i,i'} > T$ will be inflated due to winners' curse, which we correct for by assuming $K'_{i,i'}$ is sampled from the left-truncated normal distribution $N_-(c_{i,i'}, b)$, then replacing $K'_{i,i'}$ with $\hat{c}_{i,i'}$, the value that maximizes the likelihood of observing $K_{i,i'}$.

The above figure compares off-diagonal elements of $K_S$ and $K'_S$ (the sparse GRMs used by fastGWA and our approximate version of fastGWA, respectively); point colors indicate which elements are truncated by each method, while the diagonal line marks $y = x$. For the first two panels, $K'$ is constructed using 1024 SNPs (the default setting). The first panel shows that if we do not correct for winners' curse, the non-truncated elements of $K'_S$ tend to be larger than the corresponding elements of $K_S$, with the difference largest for values just above the truncation threshold. By contrast, the second panel shows that after our correction for winners' curse, the inflation is less obvious. The last two panels match the first two, except now $K'$ is constructed using 2048 SNPs. We see that the truncation threshold reduces (from about 0.15 to 0.10), and there is closer concordance between values of $K_S$ and $K'_S$.

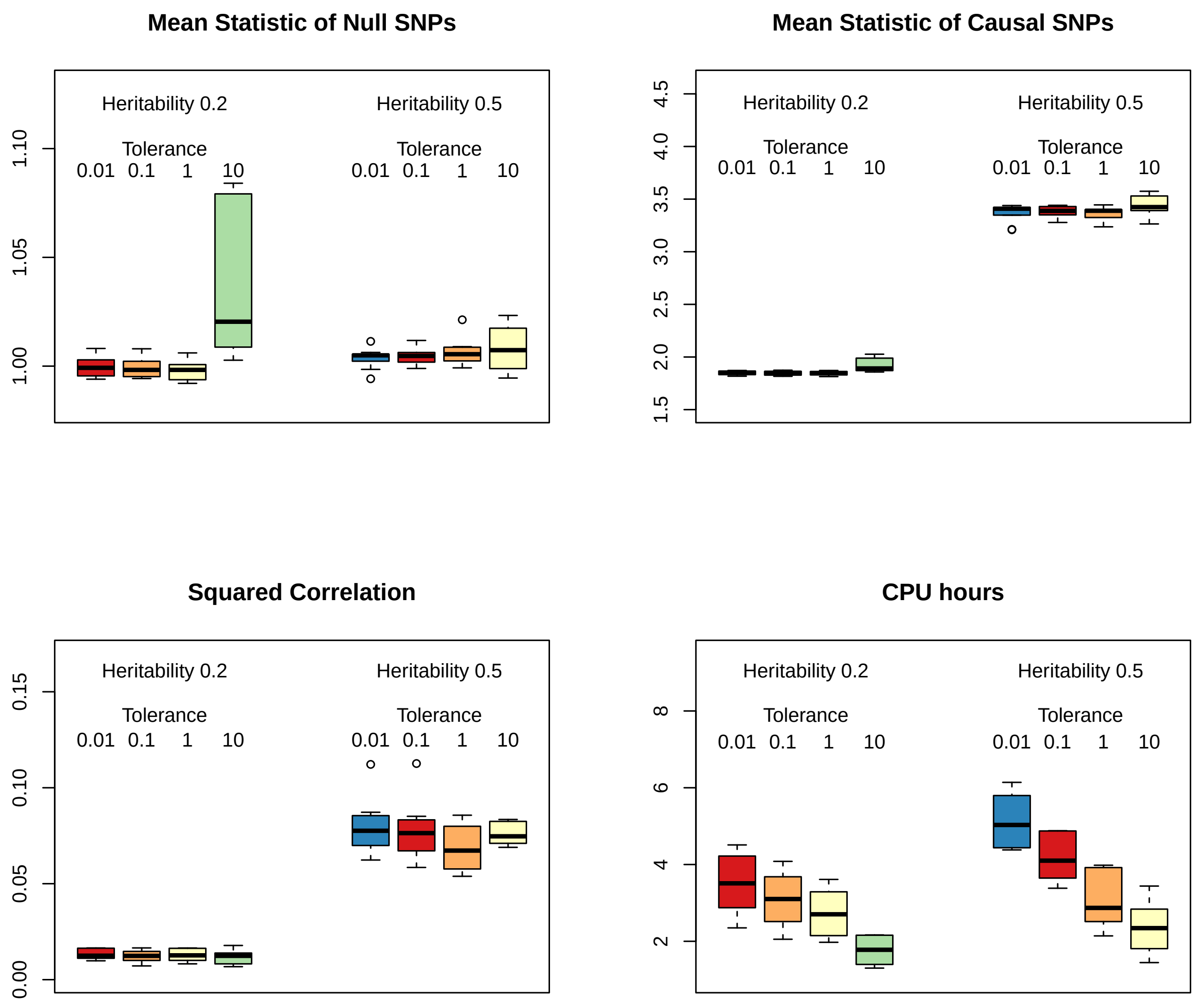

**Supplementary Figure 37: Sensitivity of our variational Bayes solver to the convergence criterion.** By default, our variational Bayes solver ignores chunks that in the previous scan caused the approximate log likelihood to change by less than $n \times 10^{-6}$. Therefore, when analyzing our simulated phenotypes for the homogeneous dataset (63 k individuals), the default tolerance threshold is $63\,000 \times 10^{-6} = 0.063$. Here we reanalyze the quantitative phenotypes varying the tolerance threshold from 0.01 to 10. For each analysis, we measure the mean $\chi^2(1)$ test statistics of null and causal SNPs (top panels), the accuracy of the Step 1 PGS (bottom left panel) and the runtime (bottom right panel). In general, we find that varying the tolerance has limited impact on the accuracy of LDAK-KVIK, albeit lower tolerances lead to longer runtimes. However, the exception is when we set the tolerance to 10 (i.e., approximately 160 times higher than the default), at which point we observe inflated test statistics.

Based on this analysis, we conclude that our default tolerance choice is reasonable.

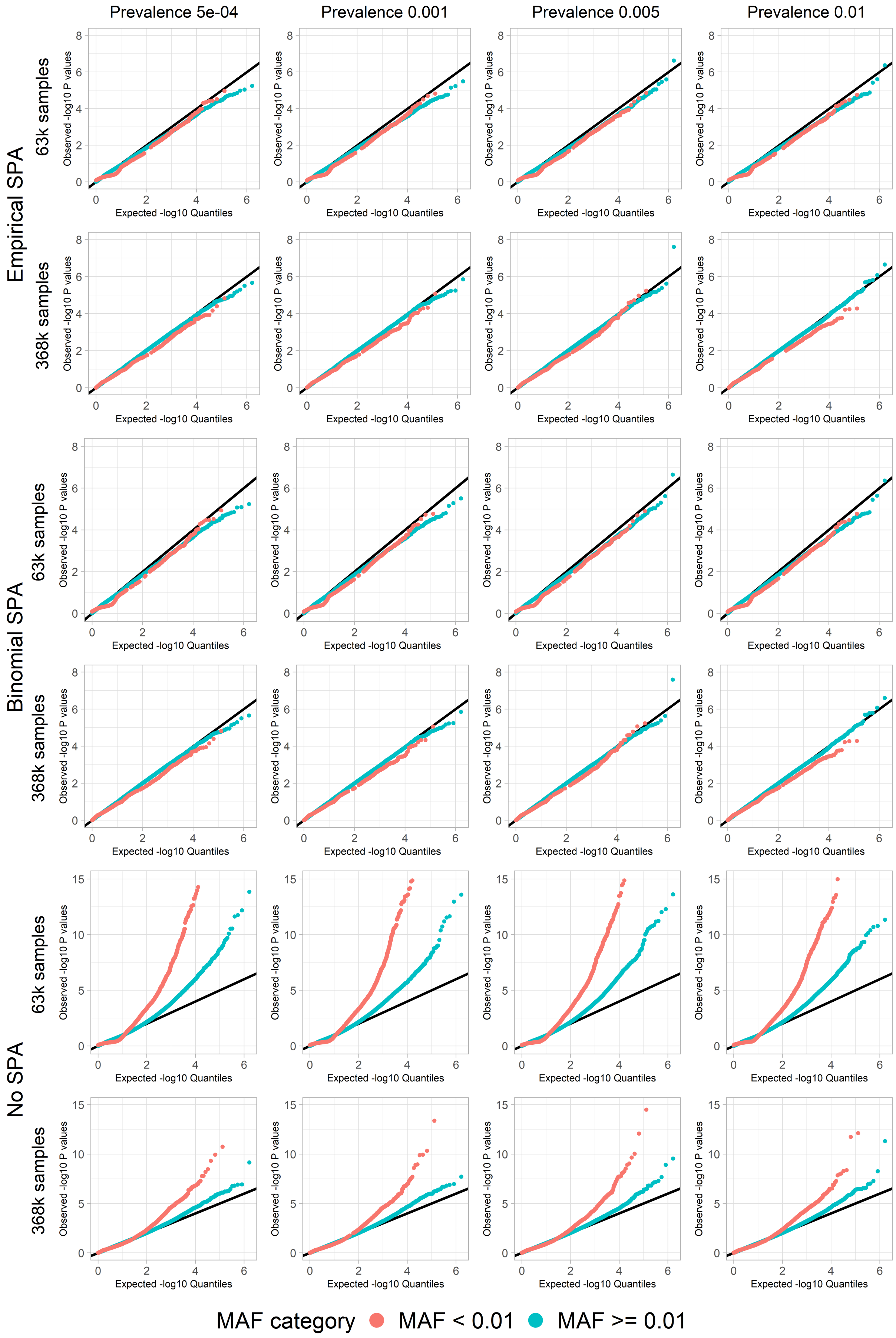

**Supplementary Figure 38: Our SPA solver controls type 1 error.**

[Please note that this figure is described on the next page.]

We simulate four binary phenotypes for both the homogeneous and white datasets (63 k and 368 k individuals, respectively). Each phenotype has heritability 0.2 and 20 k causal SNPs, with effect sizes generated assuming $\alpha = -1$, and has prevalence 1%, 0.5%, 0.1% or 0.05%. We compute $p$-values for the null SNPs using classical logistic regression, then compare ordered $-log_{10}(P)$ with the expected values under the null hypothesis; for the top two rows, we use our novel empirical SPA solver; for the middle two rows, we use a naïve Bernoulli SPA solver (we implemented the non-fast version of SPA from the fastSPA publication[31]); while for the bottom rows we do not use SPA. Point colors mark which SNPs have MAF above and below 0.01.

These figures indicate that our novel empirical SPA solver controls type 1 error, even for rare SNPs ($0.001 < \text{MAF} < 0.01$) and for very unbalanced binary datasets (for the phenotypes with prevalence 0.05%, the homogeneous and white datasets are expected to have only 32 and 184 cases, respectively). In particular, we note that our solver performs as well as the naïve Bernoulli SPA solver, despite our solver not being told the distribution of the phenotype. By contrast, we observe highly-inflated $p$-values if we do not use SPA, with the inflation highest for rare SNPs.

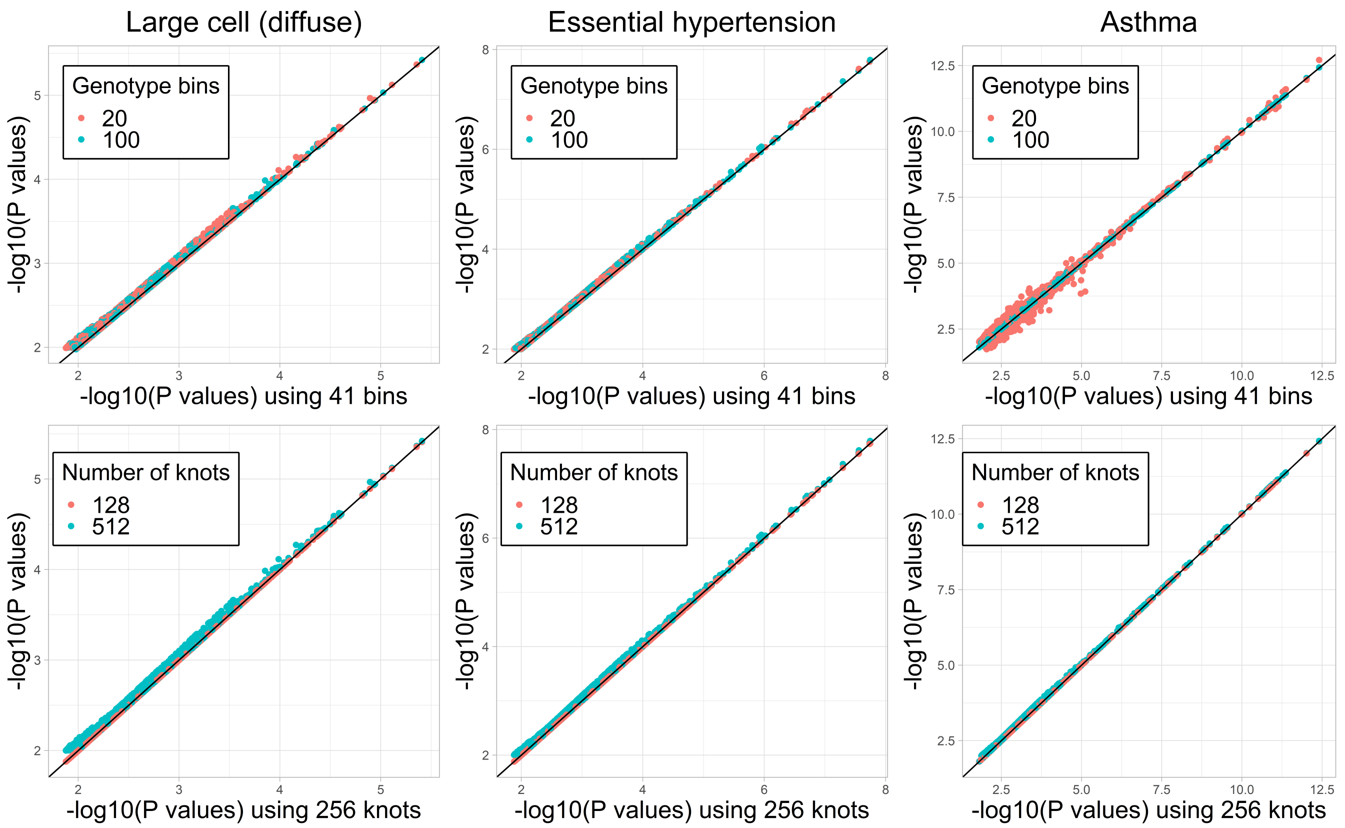

**Supplementary Figure 39: Sensitivity of our SPA solver to parameter choices.**
Our SPA solver starts by computing realizations of the cumulant generating function and its derivatives for a grid of values, whose size is determined by the numbers of bins and knots. By default, we use 41 bins and 256 knots; this figure shows the impact of changing these numbers to 20 & 100, and to 128 & 512, respectively. Specifically, we analyze three binary UK Biobank phenotypes, using 63 k individuals from the homogeneous dataset, and compare the $-log_{10}$ $p$-values of SNPs for which the SPA was used (those with uncorrected $p$-value below 0.05) when changing the numbers of bins (top panels) or knots (bottom panels). The diagonal line in each panel marks $y = x$. In general, we find that increasing the number of bins or knots has no obvious impact on the resulting $p$-values, whereas reducing either number has a small impact.

Based on this analysis, we conclude that the default numbers of bins and knots are reasonable.

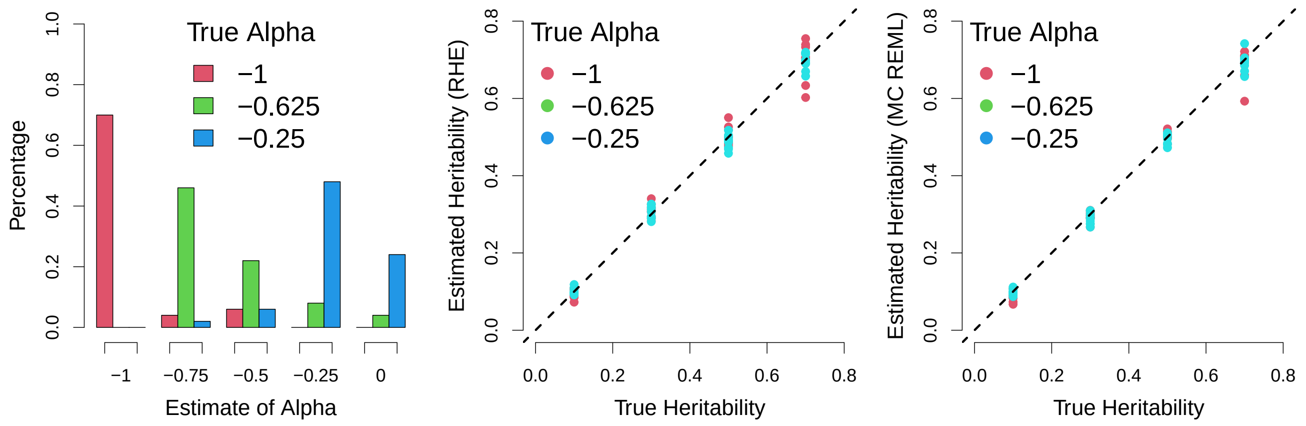

**Supplementary Figure 40: LDAK-KVIK accurately estimates $\alpha$ and $h^2$.** We simulate phenotypes for the homogeneous dataset (63 k individuals) with heritability 0.1, 0.3, 0.5 or 0.7, 10 k causal SNPs and effect sizes generated assuming $\alpha = -1$, $\alpha = -0.625$ or $\alpha = -0.25$. The left and middle panels report, respectively, estimates of $\alpha$ and $h^2$ from partitioned randomized Haseman-Elston Regression, while the right panel reports estimates of $h^2$ from Monte Carlo REML (as a reminder, LDAK-KVIK first estimates $h^2$ using partitioned randomized Haseman-Elston Regression, then updates the estimate using Monte Carlo REML).

Based on this analysis, we conclude that randomized Haseman-Elston Regression and Monte Carlo REML are effective at estimating $\alpha$ and $h^2$.

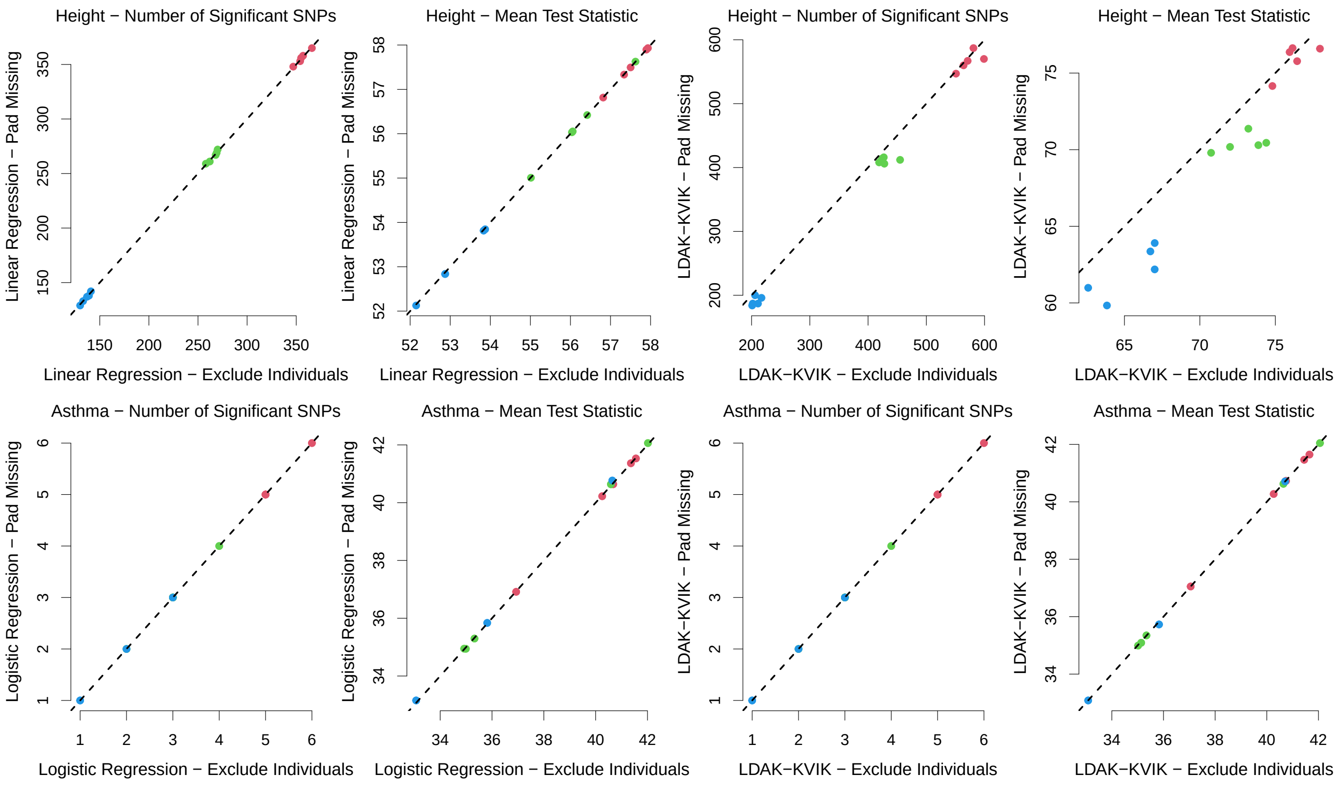

**Supplementary Figure 41: Padding missing phentoypes**

We first identify 320 k individuals in the white dataset with non-missing phenotypes for height. We then create 30 new versions of the height phenotype with 10%, 25% or 50% of values set to missing. We analyze each phenotype using classical linear regression, first excluding individuals with missing phenotypes, then "padding" missing phenotypes (i.e. analyzing all 320 k individuals, with missing phenotypes replaced by the mean). We then re-analyze each phenotype using LDAK-KVIK (first excluding individuals with missing phenotypes, then padding missing phenotypes). We then repeat this process with the phenotype asthma (except for the first sets of analyses, we use classical logistic regression instead of classical linear regression). This figure reports the number of independent, genome-wide significant loci for each phenotype (SNPs with $P < 5 \times 10{-}8$, filtered so that no pair within 1 Mb has squared correlation above 0.1), and the mean test statistic of significant SNPs, when analyzing height (top row) or asthma (bottom row); the $x$-axes correspond to analyses where individuals with missing phenotypes were excluded, while the $y$-axes correspond to analyses that padded missing phenotypes.

In general, we find that the results from analyses that used padding of missing phenotypes are very similar to those from analyses that excluded the corresponding individuals, indicating that it is valid to use padding of missing phenotypes when analyzing multiple phenotypes at the same time. In particular, we see that padding has an almost negligible impact on classical regression. When using LDAK-KVIK, we observe that padding can slightly reduce power, which reflects that the inclusion of individuals with missing phenotypes results in the Step 1 PGS having slightly lower accuracy. However, it should be noted that the reduction in power is only evident for height (i.e., a phenotype with very high heritability), and only when more than 10% of individuals are missing phenotypes (in general, we expect the proportion of individuals with missing phenotypes to be much lower).

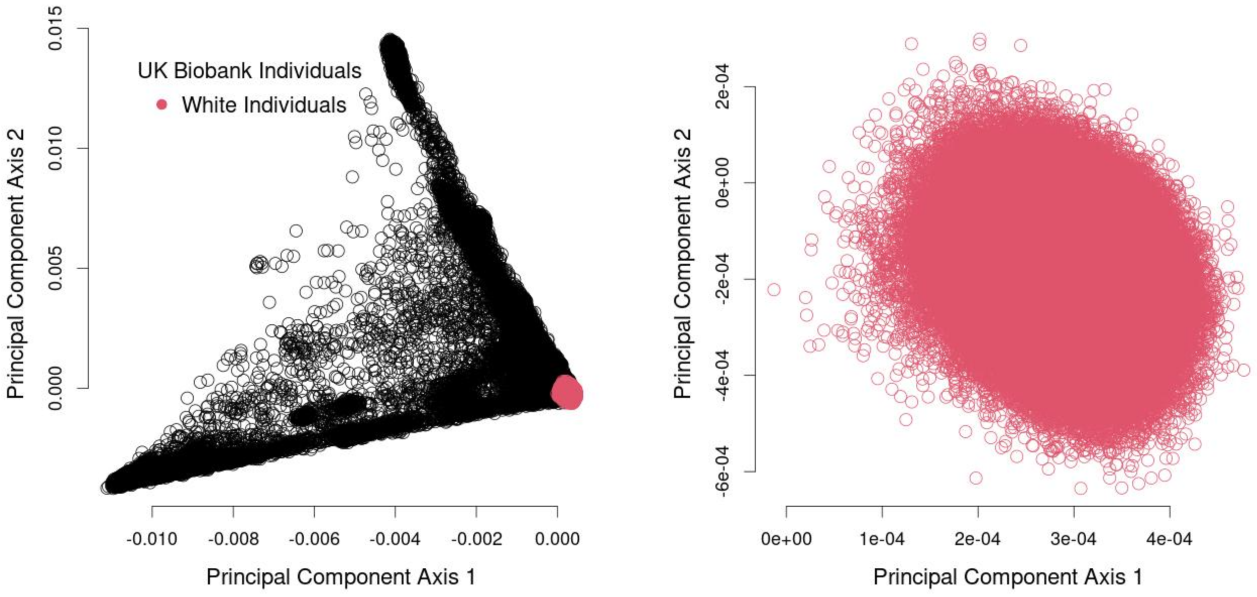

**Supplementary Figure 42: Principal component axes of the UK Biobank data.**
We perform two principal component analyses. For the left panel, we use all 487 k UK Biobank individuals, while for the right panel we use only the 409 k individuals listed in data field 22006 (those who self-identify as being white British, and whom UK Biobank determined "have very similar genetic ancestry based on a principal components analysis of the genotypes"). Note that the individuals used for the second panel are marked in red on the first panel.

| Phenotype | 690 k SNPs | | | | | | | 9.0 M SNPs |
|---|---|---|---|---|---|---|---|---|
| | CPU Hours | | | | | Numbers of | | CPU Hours |
| | **Step 1** | **Step 2** | **Step 3** | **KVIK** | **GBAT** | **Chunks** | **Updates** | **KVIK** |
| Glucose | 7.9 | 0.3 | 0.3 | 8.2 | 8.5 | 5394 | 2.8e+06 | 11.9 |
| Glycated Haemoglobin | 5.9 | 0.2 | 0.3 | 6.1 | 6.4 | 7815 | 4.5e+06 | 8.2 |
| Haemoglobin Conc. | 6.1 | 0.2 | 0.3 | 6.2 | 6.5 | 8042 | 5.3e+06 | 8.4 |
| Height | 11.4 | 0.2 | 0.3 | 11.6 | 11.9 | 13425 | 1.5e+07 | 14.3 |
| HDL Cholesterol | 6.4 | 0.2 | 0.3 | 6.6 | 6.9 | 9728 | 6.9e+06 | 8.8 |
| **Mean** | **7.5** | **0.2** | **0.3** | **7.7** | **8.0** | **8881** | **7.0e+06** | **10.3** |
| **Median** | **6.4** | **0.2** | **0.3** | **6.6** | **6.9** | **8042** | **5.3e+06** | **8.8** |
| J45 Asthma | 9.7 | 0.7 | 0.3 | 10.4 | 10.7 | 6104 | 3.1e+06 | 19.3 |
| I48 Atrial Fibrillation | 9.6 | 0.7 | 0.3 | 10.3 | 10.6 | 5394 | 3.3e+06 | 18.8 |
| I25 Ischaemic Heart Disease | 9.6 | 0.7 | 0.3 | 10.4 | 10.7 | 5394 | 3.3e+06 | 19.0 |
| K02.9 Dental Caries | 8.3 | 0.8 | 0.3 | 9.1 | 9.4 | 5394 | 2.2e+06 | 18.6 |
| I84.6 Skin Tags | 8.8 | 0.8 | 0.3 | 9.6 | 9.9 | 2697 | 1.5e+06 | 18.9 |
| **Mean** | **9.2** | **0.7** | **0.3** | **10.0** | **10.3** | **4997** | **2.7e+06** | **18.9** |
| **Median** | **9.6** | **0.7** | **0.3** | **10.3** | **10.6** | **5394** | **3.1e+06** | **18.9** |

**Supplementary Table 1: LDAK-KVIK runtimes when analyzing 368 k Individuals.**
For each of ten UK Biobank phenotypes (five quantitative and five binary), we report the CPU hours of Steps 1, 2 & 3 of LDAK-KVIK, as well as the total CPU hours for LDAK-KVIK and LDAK-KVIK-GBAT (obtained by summing the times for Steps 1 & 2, and for Steps 1, 2 & 3, respectively). We also report the number of chunks visited and number of updates of $Q_j(\gamma_j)$ by our variational Bayes solver. All analyses used the white dataset (368 k individuals and 690 k SNPs), except for the final column, which reports the total runtime of LDAK-KVIK when we increase the number of Step 2 SNPs from 690 k to 9.0 M (i.e., analyze imputed data).

| Cores | LDAK-KVIK | | | REGENIE | | |
|---|---|---|---|---|---|---|
| | **Wall Hours** | **CPU Hours** | **Speedup (%)** | **Wall Hours** | **CPU Hours** | **Speedup (%)** |
| 1 | 7.8 | 7.8 | 0 | 5.9 | 5.9 | 0 |
| 2 | 5.6 | 11.2 | 28 | 4.0 | 7.9 | 33 |
| 4 | 3.9 | 15.5 | 50 | 2.5 | 10.0 | 58 |
| 8 | 2.7 | 21.6 | 65 | 1.9 | 15.5 | 67 |

**Supplementary Table 2: The benefit of using multiple processors.**

We applied LDAK-KVIK and REGENIE to the UK Biobank phenotypes glucose, glycated haemoglobin, haemoglobin concentration, height and high-density lipoprotein (the same five quantitative phenotypes used for Supplementary Table 1). Values report the mean runtime, memory usage and speedup across the five phenotypes, when using one, two, four or eight processors (the speedup is computed relative to the analyzes using one processor). Note that we report both the wall time (the elapsed time between each job starting and finishing) and the CPU time (the wall time multipled by the number of processors used).

We see that it is possible to reduce the wall time of LDAK-KVIK by using multiple processors, and that the reductions are similar to those achieved by REGENIE. However, it should be noted that the speedups are not linear (e.g., we require four processors to reduce the runtime by half), and so using multiple cores tends to increase the CPU time. This is important because many high-performance clusters charge based on CPU time, instead of wall time, and is why we generally prefer to run LDAK-KVIK using a single processor.

| Number of | LDAK-KVIK | | | REGENIE | | |
|---|---|---|---|---|---|---|
| Phenotypes | CPU Hours | Memory (Gb) | Speedup (%) | CPU Hours | Memory (Gb) | Speedup (%) |
| 1 | 7.8 | 4.2 | 0 | 10.0 | 16 | 0 |
| 2 | 10.3 | 4.5 | 34 | 9.8 | 25 | 51 |
| 3 | 11.2 | 4.8 | 52 | 10.9 | 35 | 64 |
| 4 | 14.2 | 5.1 | 54 | 10.3 | 44 | 74 |
| 5 | 16.5 | 5.4 | 58 | 10.9 | 54 | 78 |

**Supplementary Table 3: The benefit of analyzing multiple phenotypes.**
We applied LDAK-KVIK and REGENIE to the UK Biobank phenotype glucose (the first quantitative phenotype from Supplementary Table 1, which has sample size 321 k). We first analyzed the phenotype individually, then analyzed between two and five copies of the phenotype simultaneously (we chose to duplicate the phenotype, instead of selecting unique phenotypes, to ensure that the computational requirements reflected only the increase in the number of phenotypes, and not differences in genetic architecture across phenotypes). Values report the runtime, memory usage and speedup (the latter is calculated with respect to the single-phenotype analyses). Please note that for these analyses, we used four CPUs, and so the CPU times for a single phenotype are higher than those reported in the main text.

We see that it is possible to substantially reduce the total runtime of LDAK-KVIK by analyzing multiple phenotypes. For example, it took approximately 17 CPU hours to analyze five copies of the phenotype simultaneously, whereas it would take about 39 CPU hours to analyze the five copies individually ($5 \times 7.9$). Moreover, the impact on memory demands is slight (switching from one to five copies increased the memory from 4Gb to 5Gb). Saying this, we generally recommend applying LDAK-KVIK to phenotypes individually. This is because when using a high-performance cluster, we find it is usually more efficient, in terms of cost, time to complete all analyses and ease of debugging, to run many low-resource jobs instead of fewer high-resource jobs. Furthermore, in these analyses, we found that most of the reduction in CPU hours due to analyzing multiple phenotypes was offset by the fact we used four CPUs (which as shown in Supplementary Table 2, tends to result in a halving of wall time, but a doubling in CPU time).

| MMAA Tool | Regression Model | Correction | CPU Hours | Difference |
|---|---|---|---|---|
| LDAK-KVIK | Linear | None | 0.8 | NA |
| | Logistic | None | 0.8 | 0 |
| | Logistic | Our novel SPA solver | 1.3 | 0.5 |
| | Logistic | A naïve Bernoulli SPA solver | 1.8 | 1.0 |
| REGENIE | Linear | None | 2.3 | NA |
| | Logistic | None | 2.0 | 0 |
| | Logistic | fastSPA | 5.0 | 3.0 |
| | Logistic | Approximate Firth Regression | 5.4 | 3.4 |

**Supplementary Table 4: Performance of our novel SPA solver.**

We analyzed a binary phenotype for the white dataset (368 k individuals and 690 k SNPs). We first used LDAK-KVIK to perform linear regression (without a SPA solver), followed by three versions of logistic regression: without a SPA solver, using our novel SPA solver, and using a naïve Bernoulli SPA solver (we implemented the non-fast version of SPA from the fastSPA publication[31]). We next used REGENIE to perform linear regression (without a SPA solver), followed by three versions of logistic regression: without a SPA solver, using their implementation of fastSPA, and using an approximate version of Firth Regression (the recommendation of the authors of REGENIE[6]). As well as reporting total runtime, for the analyses that use logistic regression, we also report the difference in runtime relative to non-corrected logistic regression (which is an estimate of the extra time required to compute corrected $p$-values).

We found it challenging to compare our novel SPA solver with existing solvers. This is because the runtime of each analysis depends not only on the time to perform the SPA, but also on factors such as the time to perform regular logistic regression, and the time to read in and manipulate the data. For example, while the above values suggest that our SPA solver is much faster than the implementation of fastSPA within REGENIE (1.3 CPU hours versus 5.0 CPU hours), we note that LDAK-KVIK performs non-corrected logistic regression over twice as fast as REGENIE (0.8 CPU hours versus 2.0 hours).

Nonetheless, we believe that these results show that our novel SPA solver has above-average performance. When considering absolute runtimes, we see that LDAK-KVIK can perform SPA-corrected logistic regression for a large dataset within a few CPU hours. Meanwhile, when considering relative runtimes, our solver is about twice as fast as a naïve SPA solver, and faster than the approximate Firth Regression solver used by REGENIE. Furthermore, we note that aside from its reduced runtime, our solver has the advantages that it is empirical, meaning that it can be applied to any type of phenotype, and that it does not require sparse genotypes, meaning that, for example, it can be applied to dosage data.

| | Dataset | | | | | |
|---|---|---|---|---|---|---|
| **Reported Ethnicity** | **White** | **Homogeneous** | **Family** | **Twins** | **British-Irish** | **Multi-Ancestry** |
| White | 0 | 0 | 0 | 0 | 0 | 543 |
| British | 367981 | 63000 | 63000 | 31500 (duplicated) | 51263 | 6681 |
| Irish | 0 | 0 | 0 | 0 | 11737 | 12706 |
| Any other White background | 0 | 0 | 0 | 0 | 0 | 15743 |
| Mixed | 0 | 0 | 0 | 0 | 0 | 46 |
| White and Black Caribbean | 0 | 0 | 0 | 0 | 0 | 596 |
| White and Black African | 0 | 0 | 0 | 0 | 0 | 402 |
| White and Asian | 0 | 0 | 0 | 0 | 0 | 801 |
| Any other Mixed background | 0 | 0 | 0 | 0 | 0 | 993 |
| Asian or Asian British | 0 | 0 | 0 | 0 | 0 | 42 |
| Indian | 0 | 0 | 0 | 0 | 0 | 5659 |
| Pakistani | 0 | 0 | 0 | 0 | 0 | 1745 |
| Bangladeshi | 0 | 0 | 0 | 0 | 0 | 221 |
| Any other Asian background | 0 | 0 | 0 | 0 | 0 | 1746 |
| Black or Black British | 0 | 0 | 0 | 0 | 0 | 26 |
| Caribbean | 0 | 0 | 0 | 0 | 0 | 4295 |
| African | 0 | 0 | 0 | 0 | 0 | 3202 |
| Any other Black background | 0 | 0 | 0 | 0 | 0 | 118 |
| Chinese | 0 | 0 | 0 | 0 | 0 | 1502 |
| Other ethnic group | 0 | 0 | 0 | 0 | 0 | 4351 |
| Prefer not to answer | 0 | 0 | 0 | 0 | 0 | 1582 |
| **Total** | **367981** | **63000** | **63000** | **63000** | **63000** | **63000** |

**Supplementary Table 5: Ethnic compositions of the UK Biobank datasets.**

This table provides a breakdown of our six UK Biobank datasets, based on the ethnic labels provided in data field 21000.

| Phenotype | Data Field | Sample Size | Mean | SD | Test Samples |
|---|---|---|---|---|---|
| Alanine Aminotransferase | 30620 | 350684 | 23.53 | 14.10 | 39032 |
| Albumin | 30600 | 321188 | 45.23 | 2.61 | 35786 |
| Alkaline Phosphatase | 30610 | 350811 | 83.76 | 26.52 | 39057 |
| Basophill Count | 30160 | 356431 | 0.03 | 0.05 | 39591 |
| Body Mass Index | 21001 | 366801 | 27.41 | 4.75 | 40772 |
| Calcium | 30680 | 321072 | 2.38 | 0.09 | 35770 |
| Cholesterol | 30690 | 350793 | 5.71 | 1.15 | 39054 |
| C-reactive Protein | 30710 | 350036 | 2.60 | 4.40 | 38972 |
| Creatinine | 30700 | 350616 | 72.34 | 17.93 | 39036 |
| Creatinine Enzymatic in Urine | 30510 | 357452 | 8800.26 | 5747.65 | 39738 |
| Cystatin C | 30720 | 350758 | 0.91 | 0.17 | 39058 |
| Eosinophill Percentage | 30210 | 356436 | 2.56 | 1.85 | 39591 |
| Gamma Glutamyltransferase | 30730 | 350600 | 37.39 | 41.81 | 39049 |
| Glucose | 30740 | 320821 | 5.12 | 1.21 | 35750 |
| Glycated Haemoglobin HbA1c | 30750 | 350802 | 35.97 | 6.52 | 38941 |
| Haemoglobin Concentration | 30020 | 357065 | 14.20 | 1.23 | 39654 |
| HDL Cholesterol | 30760 | 321043 | 1.45 | 0.38 | 35772 |
| Height | 50 | 367194 | 168.72 | 9.25 | 40808 |
| High Light Scatter Reticulocyte Percentage | 30290 | 351312 | 0.40 | 0.33 | 39061 |
| IGF-1 | 30770 | 348904 | 21.38 | 5.66 | 38833 |
| LDL Cholesterol | 30780 | 350138 | 3.57 | 0.87 | 38992 |
| Lymphocyte Percentage | 30180 | 356436 | 28.63 | 7.36 | 39591 |
| Mean Corpuscular Haemoglobin | 30050 | 357062 | 31.55 | 1.82 | 39654 |
| Mean Corpuscular Haemoglobin Concentration | 30060 | 357059 | 34.54 | 1.06 | 39653 |
| Mean Platelet Thrombocyte Volume | 30100 | 357056 | 9.32 | 1.08 | 39655 |
| Mean Reticulocyte Volume | 30260 | 351312 | 105.81 | 7.77 | 39061 |
| Monocyte Count | 30130 | 356431 | 0.48 | 0.22 | 39591 |
| Nucleated Red Blood Cell Count | 30170 | 356423 | 0.00 | 0.03 | 39590 |
| Phosphate | 30810 | 320580 | 1.16 | 0.16 | 35719 |
| Platelet Count | 30080 | 357061 | 253.41 | 59.89 | 39655 |
| Platelet Crit | 30090 | 357057 | 0.23 | 0.05 | 39655 |
| Platelet Distribution Width | 30110 | 357056 | 16.49 | 0.52 | 39655 |
| Red Blood Cell Erythrocyte Count | 30010 | 357064 | 4.51 | 0.41 | 39655 |
| SHBG | 30830 | 318117 | 51.86 | 27.61 | 35437 |
| Sodium in Urine | 30530 | 356709 | 76.22 | 43.61 | 39640 |
| Total Bilirubin | 30840 | 349347 | 9.13 | 4.42 | 38879 |
| Triglycerides | 30870 | 350514 | 1.76 | 1.02 | 39025 |
| Urate | 30880 | 350359 | 309.28 | 80.28 | 39015 |
| Urea | 30670 | 350556 | 5.44 | 1.39 | 39025 |
| White Blood Cell Leukocyte Count | 30000 | 357060 | 6.89 | 2.07 | 39655 |

**Supplementary Table 6: Baseline characteristics of the 40 quantitative UK Biobank phenotypes.** The sample sizes, means and standard deviations correspond to phenotyped individuals within the white dataset (367 981 white British individuals), which we used for most of our analyses of the UK Biobank phenotypes. The test samples are the number of phenotyped individuals within the independent test dataset (40 887 white British individuals), which we used for measuring the accuracy of Step 1 PGS.

| **Phenotype** | **ICD-10 Code** | **Prevalence** |
|---|---|---|
| Essential Primary Hypertension | I10 | 0.2871 |
| Disorders of Lipoprotein Metabolism and Other Lipidaemias | E78 | 0.1422 |
| Diverticular Disease of Intestine | K57 | 0.1227 |
| Chronic Ischaemic Heart Disease | I25 | 0.0941 |
| Asthma | J45 | 0.0899 |
| Gonarthrosis Arthrosis of Knee | M17 | 0.0773 |
| Non-insulin-dependent Diabetes Mellitus | E11 | 0.0715 |
| Atrial Fibrillation and Flutter | I48 | 0.0699 |
| Obesity | E66 | 0.0640 |
| Other Hypothyroidism | E03 | 0.0557 |
| Gastric ulcer | K25 | 0.0163 |
| Dental Caries Unspecified | K02.9 | 0.0102 |
| Residual Haemorrhoidal Skin Tags | I84.6 | 0.0098 |
| Anal Fistula | K60.3 | 0.0036 |
| Large Cell Diffuse | C83.3 | 0.0025 |
| Unspecified Abdominal Hernia | K46 | 0.0018 |
| Perichondritis of External Ear | H61.0 | 0.0012 |
| Septicaemia due to Other Specified Staphylococcus | A41.1 | 0.0008 |
| Other Sleep Disorders | G47.8 | 0.0004 |
| Malignant Neoplasm of Oropharynx | C10 | 0.0002 |

**Supplementary Table 7: Baseline characteristics of the 20 binary UK Biobank phenotypes.** We obtained ICD-10 codes from data field 41270. Note that there are no missing values, because everyone not recorded as being a case for a particular phenotype (i.e., not having the corresponding ICD-10 code) is automatically assumed to be a control.

#### References

1. Loh, P.-R. *et al.* Efficient Bayesian mixed-model analysis increases association power in large cohorts. *Nature genetics* **47,** 284–290 (2015).
2. Speed, D., Holmes, J. & Balding, D. J. Evaluating and improving heritability models using summary statistics. *Nature Genetics* **52,** 458–462 (2020).
3. Yang, J. *et al.* Common SNPs explain a large proportion of the heritability for human height. *Nat. Genet.* **42,** 565–569 (2010).
4. Speed, D., Hemani, G., Johnson, M. & Balding, D. Improved heritability estimation from genome-wide SNP data. *Am. J. Hum. Genet.* **91,** 1011–1021 (2012).
5. Speed, D. & Evans, D. Estimating disease heritability from complex pedigrees allowing for ascertainment and covariates. *The American Journal of Human Genetics* **111,** 680–690. ISSN: 0002-9297 (2024).
6. Mbatchou, J. *et al.* Computationally efficient whole-genome regression for quantitative and binary traits. *Nature genetics* **53,** 1097–1103 (2021).
7. Jiang, L. *et al.* A resource-efficient tool for mixed model association analysis of large-scale data. *Nature genetics* **51,** 1749–1755 (2019).
8. Yang, J., Zaitlen, N. A., Goddard, M. E., Visscher, P. M. & Price, A. L. Advantages and pitfalls in the application of mixed-model association methods. *Nature genetics* **46,** 100–106 (2014).
9. Zhou, W. *et al.* Efficiently controlling for case-control imbalance and sample relatedness in large-scale genetic association studies. *Nature genetics* **50,** 1335–1341 (2018).
10. Speed, D. *et al.* Reevaluation of SNP heritability in complex human traits. *Nat. Genet.* **49,** 986–992 (2017).
11. Zeng, J. *et al.* Widespread signatures of natural selection across human complex traits and functional genomic categories. *Nature Communications* **12,** 1164 (2021).
12. Schoech, A. P. *et al.* Quantification of frequency-dependent genetic architectures in 25 UK Biobank traits reveals action of negative selection. *Nature Communications* **10,** 790 (Feb. 2019).
13. Gazal, S. *et al.* Linkage disequilibrium-dependent architecture of human complex traits shows action of negative selection. *Nat. Genet.* **49** (2017).
14. Zeng, J. *et al.* Signatures of negative selection in the genetic architecture of human complex traits. *Nat. Genet.* **50,** 746–753 (2018).
15. Pazokitoroudi, A. *et al.* Efficient variance components analysis across millions of genomes. *Nature communications* **11,** 4020 (2020).
16. Kang, H. M. *et al.* Efficient control of population structure in model organism association mapping. *Genetics* **178,** 1709–1723 (2008).
17. Zhou, X. & Stephens, M. Genome-wide efficient mixed-model analysis for association studies. *Nature genetics* **44,** 821–824 (2012).
18. Lippert, C. *et al.* FaST linear mixed models for genome-wide association studies. *Nature methods* **8,** 833–835 (2011).

19. Svishcheva, G. R., Axenovich, T. I., Belonogova, N. M., Van Duijn, C. M. & Aulchenko, Y. S. Rapid variance components–based method for whole-genome association analysis. *Nature genetics* **44,** 1166–1170 (2012).

20. Bulik-Sullivan, B. K. *et al.* LD Score Regression distinguishes confounding from polygenicity in genome-wide association studies. *Nature genetics* **47,** 291–295 (2015).

21. Jiang, L., Zheng, Z., Fang, H. & Yang, J. A generalized linear mixed model association tool for biobank-scale data. *Nature Genetics* **53,** 1616–1621 (2021).

22. Berrandou, T., Balding, D. & Speed, D. LDAK-GBAT: Fast and powerful gene-based association testing using summary statistics. *Am. J. Hum. Genet.* **110,** 23–29 (2023).

23. Sudlow, C. *et al.* UK Biobank: an open access resource for identifying the causes of a wide range of complex diseases of middle and old age. *PLoS medicine* **12,** e1001779 (2015).

24. Bycroft, C. *et al.* The UK Biobank resource with deep phenotyping and genomic data. *Nature* **562,** 203–209 (2018).

25. Hunter-Zinck, H. *et al.* Genotyping Array Design and Data Quality Control in the Million Veteran Program. *American Journal of Human Genetics* **106,** 535–548 (2020).

26. Psychiatric GWAS Consortium Coordinating Committee. Genomewide Association Studies: History, Rationale, and Prospects for Psychiatric Disorders. *Am. J. Psychiatry* **166,** 540–556 (2009).

27. De Leeuw, C. A., Mooij, J. M., Heskes, T. & Posthuma, D. MAGMA: generalized gene-set analysis of GWAS data. *PLoS computational biology* **11,** e1004219 (2015).

28. Liu, Y. *et al.* ACAT: A Fast and Powerful p-value Combination Method for Rare-Variant Analysis in Sequencing Studies. *The American Journal of Human Genetics* **104,** 410–421 (2019).

29. Kent, W. *et al.* The human genome browser at UCSC. *Genome Res.* **12,** 996–1006 (2002).

30. Bethesda (MD): National Library of Medicine (US), N. C. f. B. I. *The NCBI handbook [Internet]* 2002.

31. Dey, R., Schmidt, E., Abecasis, G. & Lee, S. A Fast and Accurate Algorithm to Test for Binary Phenotypes and Its Application to PheWAS. *Am. J. Hum. Genet.* **101,** 37–49 (2017).

32. Hastie, T., Tibshirani, R. & Friedman, J. *The Elements of Statistical Learning* (Springer, 2001).

33. Privé, F., Aschard, H., Ziyatdinov, A. & Blum, M. Efficient analysis of large-scale genome-wide data with two R packages: bigstatsr and bigsnpr. *Bioinformatics* **34,** 2781–2787 (2018).

34. Manichaikul, A. *et al.* Robust relationship inference in genome-wide association studies. *Bioinformatics* **26,** 2867–2873 (2010).

35. The 1000 Genomes Project Consortium. A map of human genome variation from population-scale sequencing. *Nature* **467,** 1061–1073 (2010).

36. Bulik-Sullivan, B. *et al.* LD Score Regression Distinguishes Confounding from Polygenicity in Genome-Wide Association Studies. *Nat. Genet.* **47,** 291–295 (2015).

37. Firth, D. Bias reduction of maximum likelihood estimates. *Biometrika* **80,** 27–38 (1993).

38. Speed, D. & Balding, D. J. SumHer better estimates the SNP heritability of complex traits from summary statistics. *Nature genetics* **51,** 277–284 (2019).
